# Immune activation and immune-associated neurotoxicity in Long-COVID: A systematic review and meta-analysis of 82 studies comprising 58 cytokines/chemokines/growth factors

**DOI:** 10.1101/2024.02.08.24302516

**Authors:** Abbas F. Almulla, Yanin Thipakorn, Bo Zhou, Aristo Vojdani, Michael Maes

## Abstract

**Background:** Multiple studies have shown that Long COVID (LC) disease is associated with heightened immune activation, as evidenced by elevated levels of inflammatory mediators. However, there is no comprehensive meta-analysis focusing on activation of the immune inflammatory response system (IRS) and the compensatory immunoregulatory system (CIRS) along with other immune phenotypes in LC patients.

**Objectives:** This meta-analysis is designed to explore the IRS and CIRS profiles in LC patients, the individual cytokines, chemokines, growth factors, along with C-reactive protein (CRP) and immune-associated neurotoxicity.

**Methods:** To gather relevant studies for our research, we conducted a thorough search using databases such as PubMed, Google Scholar, and SciFinder, covering all available literature up to December 20th, 2023.

**Results:** The current meta-analysis encompassed 82 studies that examined multiple immune profiles, C-reactive protein, and 58 cytokines/chemokines/growth factors in 3836 LC patients versus 4537 normal controls (NC). LC patients showed significant increases in IRS/CIRS ratio (standardized mean difference (SMD:0.156, confidence interval (CI): 0.051;0.261), IRS (SMD: 0.345, CI: 0.222;0.468), M1 macrophage (SMD: 0.421, CI: 0.290;0.551), T helper (Th)1 (SMD: 0.353, CI: 0.189;0.517), Th17 (SMD: 0.492, CI: 0.332;0.651) and immune-associated neurotoxicity (SMD: 0.327 CI: 0.205;0.448). In addition, CRP and 19 different cytokines displayed significantly elevated levels in LC patients compared to NC.

**Conclusion:** LC disease is characterized by IRS activation and increased immune-associated neurotoxicity.

## Introduction

Current research continues to reveal an increasing prevalence of various symptoms among coronavirus survivors (O’Mahoney 2023). These symptoms extend beyond chronic fatigue, affective manifestations, cognitive dysfunction, and insomnia (Badenoch, Rengasamy et al. 2022, Premraj, Kannapadi et al. 2022). This emerging condition, often referred to as ‘long COVID’ (LC), poses a complex challenge for healthcare systems worldwide (Prashar 2023). The persistent nature of these symptoms significantly influences survivors’ quality of life and raises important questions about the long-term effects of COVID-19 (Maes, Al-Rubaye et al. 2022). Currently, research efforts are focused to delineate the mechanisms underlying these long-lasting manifestations and to develop effective treatments for LC.

Overall, it appears that the activation of immune-inflammatory pathways is one of the primary underlying factors in LC disease. Indeed, accumulating evidence suggests that LC is accompanied by a chronic immune-inflammatory response, as indicated by elevated levels of interferon (IFN)-γ, IFN-β, IFN-λ1, CXCL10, and CXCL9 (Peluso, Lu et al. 2021, Phetsouphanh, Darley et al. 2022). Espín et al.’s scoping review revealed that cytokine groups comprising IFN-β, pentraxin 3 (PTX3); IFN-β, PTX3, IFN-γ; IFN-β, PTX3, IFN-λ2/3, IL-6; and a panel of 29 cytokines demonstrated relatively high accuracy in differentiating LC patients from asymptomatic controls (Espín, Yang et al. 2023).

Immune cell phenotyping has revealed an increased frequency of CD8+ T-cell, Immunoglobulin and Mucin-domain containing-3 (TIM-3)+ and CD8+, Programmed Cell Death Protein 1 (PD-1)+ TIM-3+ T cells, suggesting a chronic activation of CD8+ T cells in LC patients (Phetsouphanh, Darley et al. 2022). In LC patients, there was a notable increase in the frequencies of CD14+CD16+ inflammatory monocytes and plasmacytoid dendritic cells marked, suggesting sustained T cell activation, likely due to persistent inflammation or antigen presentation (Phetsouphanh, Darley et al. 2022). LC patients also exhibited elevated levels of highly cytotoxic cell populations, memory natural killer (NK) cells, and persistent antigen-specific responses in CD4+ and T follicular helper cells, indicating a robust antiviral immune response (Galán, Vigón et al. 2022).

A recent meta-analysis has revealed that patients with LC have markedly higher levels of inflammatory and vascular mediators, including C-reactive protein (CRP), D-dimer, lactate dehydrogenase (LDH), and leukocytes (Yong, Halim et al. 2023). In addition, there was an observed elevation in lymphocytes and IL-6 in these patients (Yong, Halim et al. 2023). Yin et al. (2023) also reported a significant increase in IL-6 levels in LC patients compared to healthy controls (Yin, Agbana et al. 2023).

Importantly, numerous prior studies have confirmed that the phenome of LC comprises interrelated increases in chronic fatigue syndrome (CFS), depression, and anxiety symptoms, which were delineated as core symptoms of LC disease (Al-Hadrawi, Al-Rubaye et al. 2022, Al-Hakeim, Al-Rubaye et al. 2022, Al-Hakeim, Al-Rubaye et al. 2023, Al-Hakeim, Khairi Abed et al. 2023, Almulla, Al-Hakeim et al. 2023). Moreover, this LC symptom complex was strongly predicted by the severity of acute COVID-19 infection, inflammatory mediators (CRP and the NLRP-3 inflammasome), and oxidative stress biomarkers (Al-Hadrawi, Al-Rubaye et al. 2022, Al-Hakeim, Al-Rubaye et al. 2022, Al-Hakeim, Al-Rubaye et al. 2023). This is highly relevant because CFS, depression and anxiety are characterized by aberrations in the immune-inflammatory response system (IRS), the compensatory immunoregulatory system (CIRS), and the IRS/CIRS ratio (Maes 2009, Maes, Mihaylova et al. 2012, Maes, Twisk et al. 2012, Maes and Carvalho 2018). Furthermore, CFS and affective syndromes are associated with activation of different IRS profiles, including the classical M1 macrophage, alternative M2 macrophage, T helper (Th)-1, Th-2, and Th-17 profiles, as well as an immune-associated profiles that may cause increased neurotoxicity (Maes 2009, Maes, Mihaylova et al. 2012, Maes, Twisk et al. 2012, Maes and Carvalho 2018). Electronic Supplementary File (ESF)-1 Table 1 lists the cytokines and chemokines that were used to compute the above immune profiles and ESF-1, table 2 lists how the immune profiles were composed. The cytokines and chemokines belonging to the neurotoxicity profile all show a potential to damage the function or structure of neuronal cells or affect neuroplasticity or neurogenesis (Maes and Carvalho 2018, Maes, Rachayon et al. 2022, Almulla, Abbas Abo Algon et al. 2023, Almulla, Abdul Jaleel et al. 2023, Sreenivas, Maes et al. 2024). Consequently, increased immune-associated neurotoxicity could explain the pathophysiology of the neuro-psychiatric symptoms of LC disease.

**Table 1:**
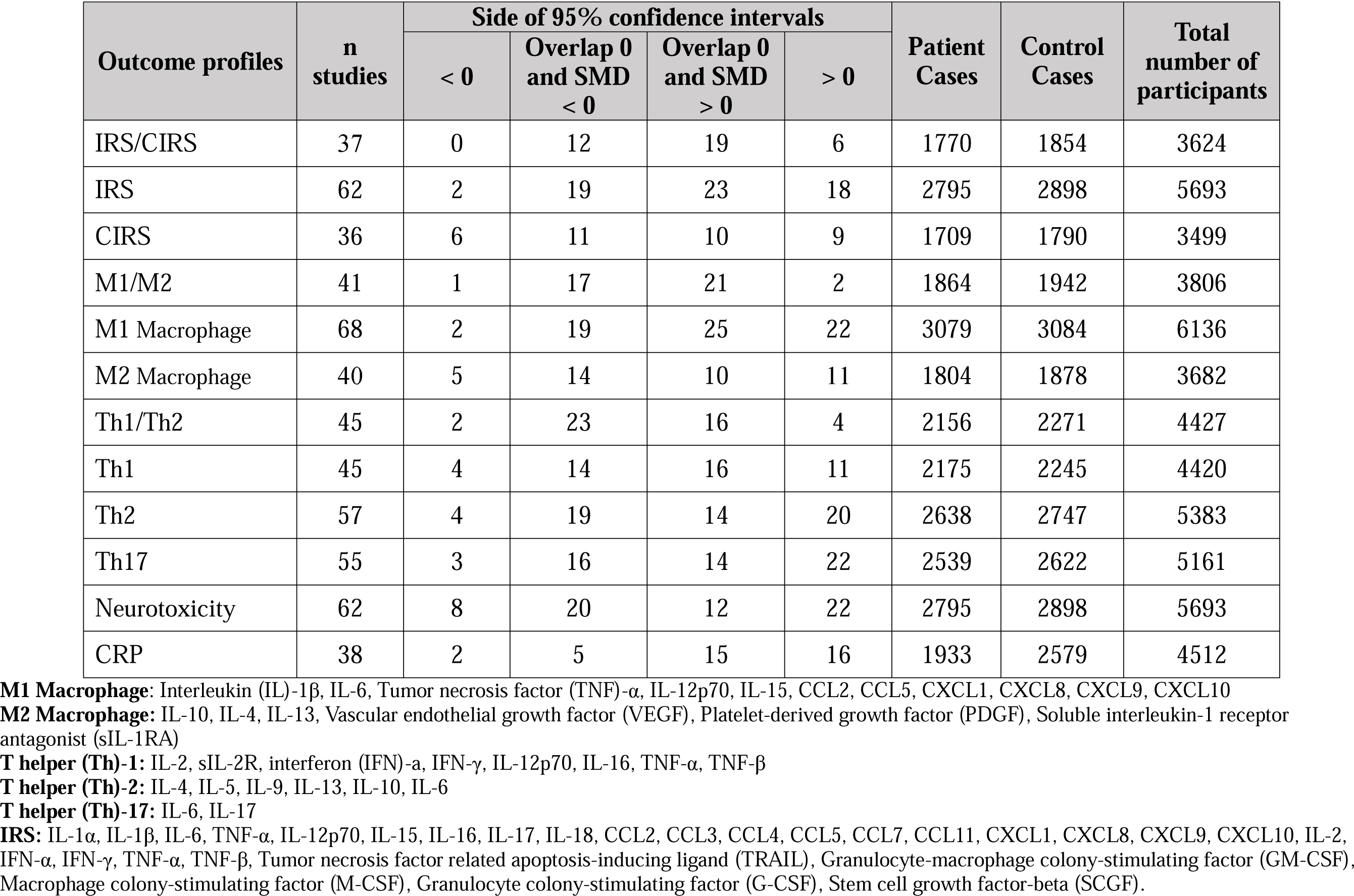
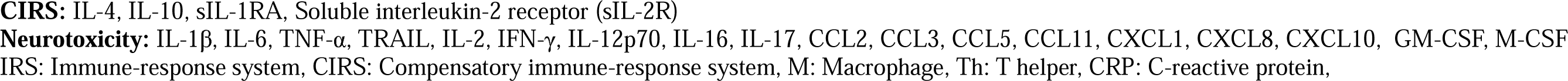
The outcomes and number of patients with Long COVID (LC) disease and healthy control along with the side of standardized mean difference (SMD) and the 95% confidence intervals with respect to zero SMD.

| Outcome profiles | n studies | Side of 95% confidence intervals |  |  |  | Patient Cases | Control Cases | Total number of participants |
| --- | --- | --- | --- | --- | --- | --- | --- | --- |
|  |  | < 0 | Overlap 0 and SMD < 0 | Overlap 0 and SMD > 0 | > 0 |  |  |  |
| IRS/CIRS | 37 | 0 | 12 | 19 | 6 | 1770 | 1854 | 3624 |
| IRS | 62 | 2 | 19 | 23 | 18 | 2795 | 2898 | 5693 |
| CIRS | 36 | 6 | 11 | 10 | 9 | 1709 | 1790 | 3499 |
| M1/M2 | 41 | 1 | 17 | 21 | 2 | 1864 | 1942 | 3806 |
| M1 Macrophage | 68 | 2 | 19 | 25 | 22 | 3079 | 3084 | 6136 |
| M2 Macrophage | 40 | 5 | 14 | 10 | 11 | 1804 | 1878 | 3682 |
| Th1/Th2 | 45 | 2 | 23 | 16 | 4 | 2156 | 2271 | 4427 |
| Th1 | 45 | 4 | 14 | 16 | 11 | 2175 | 2245 | 4420 |
| Th2 | 57 | 4 | 19 | 14 | 20 | 2638 | 2747 | 5383 |
| Th17 | 55 | 3 | 16 | 14 | 22 | 2539 | 2622 | 5161 |
| Neurotoxicity | 62 | 8 | 20 | 12 | 22 | 2795 | 2898 | 5693 |
| CRP | 38 | 2 | 5 | 15 | 16 | 1933 | 2579 | 4512 |

**M1 Macrophage:** Interleukin (IL)-1 $\beta$ , IL-6, Tumor necrosis factor (TNF)- $\alpha$ , IL-12p70, IL-15, CCL2, CCL5, CXCL1, CXCL8, CXCL9, CXCL10
**M2 Macrophage:** IL-10, IL-4, IL-13, Vascular endothelial growth factor (VEGF), Platelet-derived growth factor (PDGF), Soluble interleukin-1 receptor antagonist (sIL-1RA)
**T helper (Th)-1:** IL-2, sIL-2R, interferon (IFN)- $\alpha$ , IFN-γ , IL-12p70, IL-16, TNF-α , TNF-β
**T helper (Th)-2:** IL-4, IL-5, IL-9, IL-13, IL-10, IL-6
**T helper (Th)-17:** IL-6, IL-17
**IRS:** IL-1α , IL-1β , IL-6, TNF-α , IL-12p70, IL-15, IL-16, IL-17, IL-18, CCL2, CCL3, CCL4, CCL5, CCL7, CCL11, CXCL1, CXCL8, CXCL9, CXCL10, IL-2, IFN-α , IFN-γ , TNF-α , TNF-β , Tumor necrosis factor related apoptosis-inducing ligand (TRAIL), Granulocyte-macrophage colony-stimulating factor (GM-CSF), Macrophage colony-stimulating factor (M-CSF), Granulocyte colony-stimulating factor (G-CSF), Stem cell growth factor-beta (SCGF).
**CIRS:** IL-4, IL-10, sIL-1RA, Soluble interleukin-2 receptor (sIL-2R)
**Neurotoxicity:** IL-1β , IL-6, TNF-α , TRAIL, IL-2, IFN-γ , IL-12p70, IL-16, IL-17, CCL2, CCL3, CCL5, CCL11, CXCL1, CXCL8, CXCL10, GM-CSF, M-CSF
IRS: Immune-response system, CIRS: Compensatory immune-response system, M: Macrophage, Th: T helper, CRP: C-reactive protein,

**Table 2:** Results of meta-analysis performed on several outcomes (Inflammatory profiles and solitary cytokines) variables with.

| Outcome feature sets | n | Groups | SMD | 95% CI | z | p | Q | df | p | I<sup>2</sup> (%) | $\tau^2$ | T |
| --- | --- | --- | --- | --- | --- | --- | --- | --- | --- | --- | --- | --- |
| IRS/CIRS | 37 | Overall | 0.126 | 0.024;0.228 | 2.421 | 0.015 | 58.862 | 36 | 0.009 | 38.840 | 0.032 | 0.180 |
| IRS | 62 | Overall | 0.247 | 0.130;0.363 | 4.150 | <0.0001 | 218.455 | 61 | <0.0001 | 72.077 | 0.139 | 0.372 |
| CIRS | 36 | Overall | 0.070 | -0.110;0.250 | 0.760 | 0.447 | 181.135 | 35 | <0.0001 | 80.677 | 0.214 | 0.463 |
| M1/M2 | 41 | Overall | 0.061 | -0.061;0.183 | 0.982 | 0.326 | 98.357 | 40 | <0.0001 | 59.332 | 0.078 | 0.280 |
| M1 Macrophage | 68 | Overall | 0.327 | 0.200;0.454 | 5.041 | <0.0001 | 311.920 | 67 | <0.0001 | 78.520 | 0.200 | 0.447 |
| M2 Macrophage | 40 | Overall | 0.125 | -0.052;0.301 | 1.387 | 0.165 | 204.570 | 39 | <0.0001 | 80.936 | 0.229 | 0.478 |
| Th1/Th2 | 45 | Overall | 0.027 | -0.088;0.124 | 0.455 | 0.649 | 113.959 | 44 | <0.0001 | 61.390 | 0.080 | 0.284 |
| Th1 | 45 | Overall | 0.230 | 0.073;0.388 | 2.869 | 0.004 | 218.508 | 44 | <0.0001 | 79.863 | 0.203 | 0.451 |
| Th2 | 57 | Overall | 0.241 | 0.103;0.380 | 3.422 | 0.001 | 270.383 | 56 | <0.0001 | 79.289 | 0.201 | 0.448 |
| Th17 | 55 | Overall | 0.316 | 0.163;0.468 | 4.067 | <0.0001 | 312.028 | 54 | <0.0001 | 82.694 | 0.250 | 0.500 |
| Neurotoxicity | 62 | Overall | 0.260 | 0.143;0.377 | 4.347 | <0.0001 | 220.506 | 61 | <0.0001 | 72.336 | 0.141 | 0.375 |
| CRP | 38 | Overall | 0.507 | 0.299;0.715 | 4.779 | <0.0001 | 355.004 | 37 | <0.0001 | 89.578 | 0.365 | 0.604 |

**M1 Macrophage:** Interleukin (IL)-1 $\beta$ , IL-6, Tumor necrosis factor (TNF)- $\alpha$ , IL-12p70, IL-15, CCL2, CCL5, CXCL1, CXCL8, CXCL9, CXCL10
**M2 Macrophage:** IL-10, IL-4, IL-13, Vascular endothelial growth factor (VEGF), Platelet-derived growth factor (PDGF), Soluble interleukin-1 receptor antagonist (sIL-1RA)
**T helper (Th)-1:** IL-2, sIL-2R, interferon (IFN)- $\alpha$ , IFN-γ , IL-12p70, IL-16, TNF-α , TNF-β
**T helper (Th)-2:** IL-4, IL-5, IL-9, IL-13, IL-10, IL-6
**T helper (Th)-17:** IL-6, IL-17
**IRS:** IL-1α , IL-1β , IL-6, TNF-α , IL-12p70, IL-15, IL-16, IL-17, IL-18, CCL2, CCL3, CCL4, CCL5, CCL7, CCL11, CXCL1, CXCL8, CXCL9, CXCL10, IL-2, IFN-α , IFN-γ , TNF-α , TNF-β , Tumor necrosis factor related apoptosis-inducing ligand (TRAIL), Granulocyte-macrophage colony-stimulating factor (GM-CSF), Macrophage colony-stimulating factor (M-CSF), Granulocyte colony-stimulating factor (G-CSF), Stem cell growth factor-beta (SCGF).
**CIRS:** IL-4, IL-10, sIL-1RA, Soluble interleukin-2 receptor (sIL-2R)
**Neurotoxicity:** IL-1β , IL-6, TNF-α , TRAIL, IL-2, IFN-γ , IL-12p70, IL-16, IL-17, CCL2, CCL3, CCL5, CCL11, CXCL1, CXCL8, CXCL10, GM-CSF, M-CSF
IRS: Immune-response system, CIRS: Compensatory immune-response system, M: Macrophage, Th: T helper, CRP: C-reactive protein,

However, the two meta-analyses that examined immune functions in LC did not provide a comprehensive examination of the immune profiles, including the neurotoxicity profile (Yin, Agbana et al. 2023, Yong, Halim et al. 2023). Therefore, a more extensive examination of cytokines/chemokines/growth factor profiles in LC would be instrumental in elucidating the mechanisms of this illness. Hence, the objective of the present meta-analysis is to delineate the precise aberrations in IRS, CIRS, M1, M2 macrophages, T helper (Th)1, Th2, Th17, and neurotoxicity profiles in patients with LC. Moreover, we will also examine CRP and solitary levels of 58 different cytokines/chemokines/growth factors in LC disease. ESF-1, Table 1 lists all cytokines, chemokines and growth factors that were examined in the present study.

## Materials and method

In conducting this study, we rigorously applied several methodological frameworks, including the Preferred Reporting Items for Systematic Reviews and Meta-Analyses (PRISMA) 2020 (Page, McKenzie et al. 2021), the Cochrane Handbook for Systematic Reviews and Interventions (Cumpston, Li et al. 2019), and guidelines for Meta-Analyses of Observational Studies in Epidemiology. This analysis involved patients with LC disease and normal controls, focusing on peripheral cytokines, chemokines and growth factors, and acute phase protein, namely CRP. We then calculated composite indices reflecting different immune profiles based on individual cytokines and chemokines, including IRS, CIRS, neurotoxicity, M1 and M2 macrophages, T helper 1,2 and 17, IRS/CIRS, M1/M2 and Th1 / Th2 ratios (see ESF-1, table 2).

### Search strategy

In our quest to accumulate exhaustive data on immune profiles in LC, we sourced electronic databases such as PubMed/MEDLINE, Google Scholar, and SciFinder. This search, spanning from August until December 15th, 2023, was guided by specific keywords and Mesh terms listed in Table 3 of the ESF-1. To ensure comprehensive coverage, we also examined reference lists from relevant studies and earlier meta-analyses, thereby leaving no significant study unconsidered.

**Table 3:**
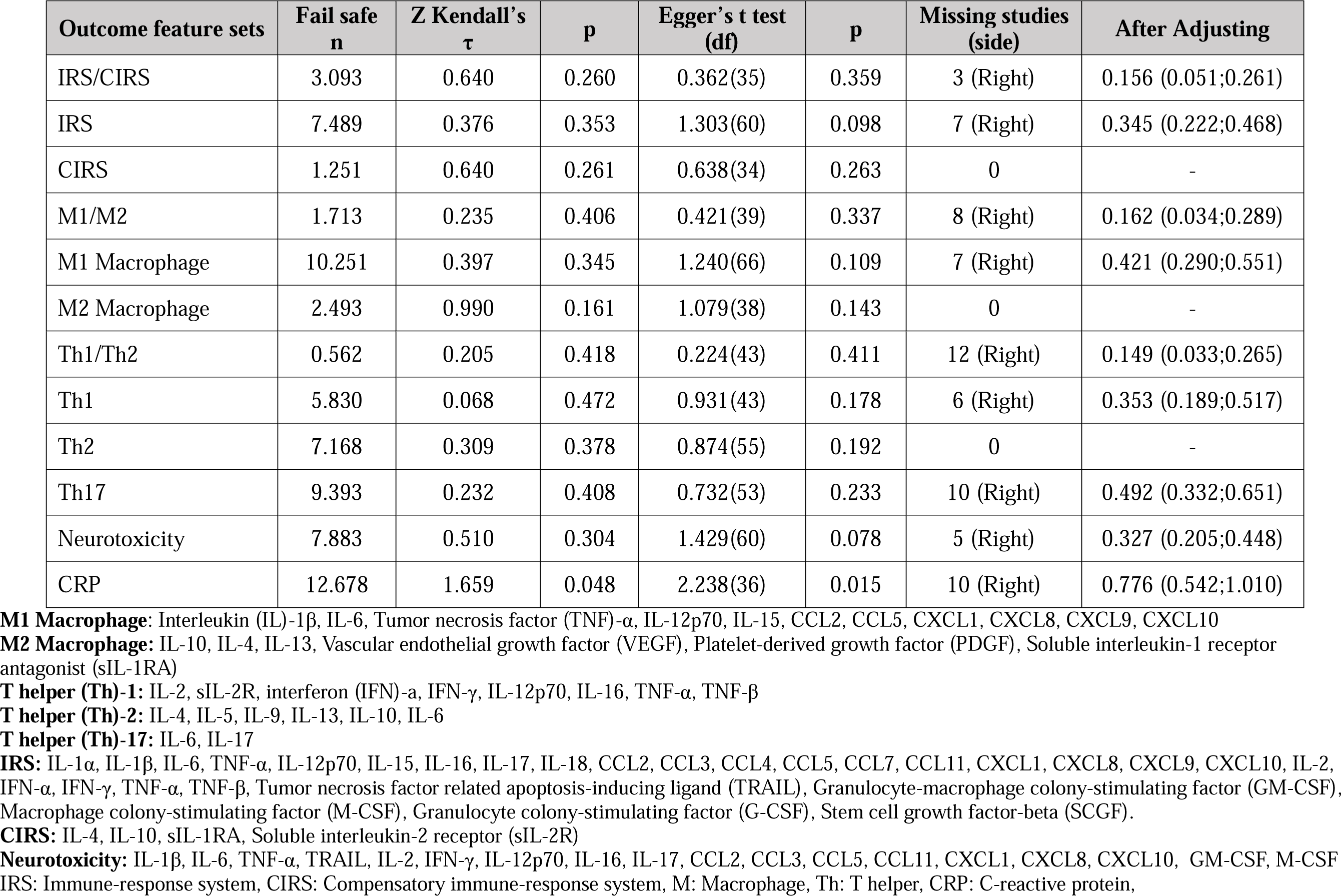
Results on publication bias.

| Outcome feature sets | Fail safe<br>n | Z Kendall's<br>$\tau$ | p | Egger's t test<br>(df) | p | Missing studies<br>(side) | After Adjusting |
| --- | --- | --- | --- | --- | --- | --- | --- |
| IRS/CIRS | 3.093 | 0.640 | 0.260 | 0.362(35) | 0.359 | 3 (Right) | 0.156 (0.051;0.261) |
| IRS | 7.489 | 0.376 | 0.353 | 1.303(60) | 0.098 | 7 (Right) | 0.345 (0.222;0.468) |
| CIRS | 1.251 | 0.640 | 0.261 | 0.638(34) | 0.263 | 0 | - |
| M1/M2 | 1.713 | 0.235 | 0.406 | 0.421(39) | 0.337 | 8 (Right) | 0.162 (0.034;0.289) |
| M1 Macrophage | 10.251 | 0.397 | 0.345 | 1.240(66) | 0.109 | 7 (Right) | 0.421 (0.290;0.551) |
| M2 Macrophage | 2.493 | 0.990 | 0.161 | 1.079(38) | 0.143 | 0 | - |
| Th1/Th2 | 0.562 | 0.205 | 0.418 | 0.224(43) | 0.411 | 12 (Right) | 0.149 (0.033;0.265) |
| Th1 | 5.830 | 0.068 | 0.472 | 0.931(43) | 0.178 | 6 (Right) | 0.353 (0.189;0.517) |
| Th2 | 7.168 | 0.309 | 0.378 | 0.874(55) | 0.192 | 0 | - |
| Th17 | 9.393 | 0.232 | 0.408 | 0.732(53) | 0.233 | 10 (Right) | 0.492 (0.332;0.651) |
| Neurotoxicity | 7.883 | 0.510 | 0.304 | 1.429(60) | 0.078 | 5 (Right) | 0.327 (0.205;0.448) |
| CRP | 12.678 | 1.659 | 0.048 | 2.238(36) | 0.015 | 10 (Right) | 0.776 (0.542;1.010) |

**M1 Macrophage:** Interleukin (IL)-1 $\beta$ , IL-6, Tumor necrosis factor (TNF)- $\alpha$ , IL-12p70, IL-15, CCL2, CCL5, CXCL1, CXCL8, CXCL9, CXCL10
**M2 Macrophage:** IL-10, IL-4, IL-13, Vascular endothelial growth factor (VEGF), Platelet-derived growth factor (PDGF), Soluble interleukin-1 receptor antagonist (sIL-1RA)
**T helper (Th)-1:** IL-2, sIL-2R, interferon (IFN)- $\alpha$ , IFN-γ , IL-12p70, IL-16, TNF-α , TNF-β
**T helper (Th)-2:** IL-4, IL-5, IL-9, IL-13, IL-10, IL-6
**T helper (Th)-17:** IL-6, IL-17
**IRS:** IL-1α , IL-1β , IL-6, TNF-α , IL-12p70, IL-15, IL-16, IL-17, IL-18, CCL2, CCL3, CCL4, CCL5, CCL7, CCL11, CXCL1, CXCL8, CXCL9, CXCL10, IL-2, IFN-α , IFN-γ , TNF-α , TNF-β , Tumor necrosis factor related apoptosis-inducing ligand (TRAIL), Granulocyte-macrophage colony-stimulating factor (GM-CSF), Macrophage colony-stimulating factor (M-CSF), Granulocyte colony-stimulating factor (G-CSF), Stem cell growth factor-beta (SCGF).
**CIRS:** IL-4, IL-10, sIL-1RA, Soluble interleukin-2 receptor (sIL-2R)
**Neurotoxicity:** IL-1β , IL-6, TNF-α , TRAIL, IL-2, IFN-γ , IL-12p70, IL-16, IL-17, CCL2, CCL3, CCL5, CCL11, CXCL1, CXCL8, CXCL10, GM-CSF, M-CSF
IRS: Immune-response system, CIRS: Compensatory immune-response system, M: Macrophage, Th: T helper, CRP: C-reactive protein,

### Eligibility criteria

This meta-analysis primarily focused on articles from peer-reviewed journals in English. However, it also considered grey literature and articles in Thai, French, Spanish, Turkish, German, Italian, and Arabic. The studies selected were observational, either case-control or cohort, involving controls and examining cytokines, chemokines and growth factors. The included LC patients were diagnosed based on the World Health Organization (WHO) criteria (World Health Organization 2021). Studies with baseline assessments of immune biomarkers in follow-up were also included. Studies that used unusual media (e.g., hair, saliva, whole blood, platelet-rich plasma), genetics, translational research, or did not provide a control group were not included. Studies not providing mean and standard deviation (SD) or standard error (SE) of biomarkers were initially excluded, but the authors were contacted for this information. In the absence of a response, we computed) orean (SD) from available data using the approach of (Wan, Wang et al. 2014) and an online Mean Variance Estimation tool (hkbu.edu.hk), or estimated the mean (SD) from graphical data using Web Plot Digitizer (https://automeris.io/WebPlotDigitizer/).

### Primary and secondary outcomes

This meta-analysis’s essential findings, presented in **Table 1**, include data on the composite scores of IRS/CIRS ratio, IRS, CIRS, M1/M1 and Th1/Th2 ratios, M1, M2, Th1, Th2, Th17, and immune-related neurotoxicity profiles. Upon determining the significance of these composite measures, we further investigated their individual indicators as secondary outcomes. These encompass solitary levels of CRP and 58 cytokines/chemokines/growth factors listed in ESF-1, Table 1.

### Screening and data extraction

The initial assessment of studies for inclusion in this meta-analysis was conducted by two researchers, AA and YT. They screened study titles and abstracts against set inclusion criteria. After this initial screening, we acquired and reviewed the full texts of studies that appeared to meet our criteria while excluding those that failed to meet our exclusion standards. AA and YT then meticulously catalogued relevant data into a custom-designed EXCEL file. This file captured details such as author names, dates of studies, biomarker names, mean and SD of biomarkers, participant numbers in patient and control groups, and sample sizes of the studies. Additionally, the file included information about the study design, sample types (e.g., serum, plasma, CSF, brain tissues, blood cells), post COVID period, number of patients admitted to the intensive care unit (ICU) during acute stage of illness, mean (SD) of participants’ ages, gender, post-COVID period, admission to the intensive care unit (ICU) during the acute stage of illness, and geographic location of the study. Any discrepancies in data entry or interpretation were resolved by consulting with the senior author, MM.

For assessing the methodological quality of the studies included in this research, we utilized the Immunological Confounder Scale (ICS), as proposed by (Andres-Rodriguez, Borras et al. 2020) and later modified by the last author (MM) specifically for studies concerning immune activation in LC. This scale includes two distinct assessment tools: the Quality Scale and the Redpoints Scale, both detailed in Table 4 of the ESF-1. These scales have been previously employed in meta-analyses focusing on lipid peroxidation and tryptophan catabolite data in those with affective disorders (Almulla, Thipakorn et al. 2022, Almulla, Thipakorn et al. 2023). The Quality Scale, with a scoring range from 0 (indicating lower quality) to 10 (representing higher quality), examines aspects such as the sample size, the control over confounding variables, and the sampling duration. The Redpoints Scale, in contrast, is tailored to identify potential biases in immune activation in LC studies by assessing how well critical confounding factors are controlled. A score of 0 on this scale signifies the highest level of control, whereas a score of 26 reflects a complete lack of control over confounding variables.

### Data analysis

In this meta-analysis, we utilized CMA V4 software and adhered to PRISMA guidelines, as detailed in Table 5 of the ESF-1. Our analysis required at least two studies per identified biomarker. For calculating IRS, we assumed dependence and incorporated various relevant inflammatory biomarkers into the CMA, including IL-1α, IL-1β, IL-6, TNF-α, IL-12p70, IL-15, IL-16, IL-17, IL-18, CCL2, CCL3, CCL4, CCL5, CCL7, CCL11, CXCL1, CXCL8, CXCL9, CXCL10, IL-2, IFN-α, IFN-γ, TNF-β, GM-CSF, and G-CSF. The CIRS calculation included IL-4, IL-10, sIL-1RA, and sIL-2R. In the case of neurotoxicity, we included biomarkers such as IL-1β, IL-6, TNF-α, IL-2, IFN-γ, IL-12p70, IL-16, IL-17, CCL2, CCL3, CCL5, CCL11, CXCL1, CXCL8, CXCL10, GM-CSF, and M-CSF. The classical M1 macrophage profile was computed with the inclusion of IL-1β, IL-6, TNF-α, IL-12p70, IL-15, CCL2, CCL5, CXCL1, CXCL8, CXCL9, CXCL10. The alternative M2 macrophage profile included IL-10, IL-4, IL-13, VEGF, PDGF, and sIL-1RA. Additionally, the current meta-analysis computed the a) Th1 profile by including IL-2, sIL-2R, IFN-α, IFN-γ, IL-12p70, IL-16, TNF-α, and TNF-β, b) Th2 by including IL-4, IL-5, IL-9, IL-13, IL-10, and IL-6, and c) Th17 by including IL-6 and IL-17. Moreover, in the current meta-analysis, we computed the IRS/CIRS ratio by selecting a positive effect size for all IRS data and a negative effect size for CIRS biomarkers in comparing patients with normal controls while assuming dependence. Likewise, M1/M2 and Th1/Th2 ratios were computed by selecting positive effect sizes for M1 and Th1 and negative effect sizes for M2 and Th2.

In our study, we applied a random-effects model using constrained maximum likelihood for aggregating effect sizes. We considered a p-value less than 0.05 (two-tailed) as statistically significant and reported effect sizes as standardized mean differences (SMD) with 95% confidence intervals (95% CI). For categorizing effect sizes, we used SMD values of 0.80, 0.5, and 0.20 to denote large, moderate, and small effects, respectively, following Cohen’s guidelines (Cohen 2013). Heterogeneity was assessed through tau-squared statistics, Q, and I^2^ values, a method consistent with previous meta-analyses (Almulla, Supasitthumrong et al. 2022, Almulla, Thipakorn et al. 2022, Almulla, Vasupanrajit et al. 2022, Vasupanrajit, Jirakran et al. 2022, Almulla, Thipakorn et al. 2023). Meta-regression was employed to identify factors contributing to heterogeneity. Subgroup analysis, considering each determinant as a separate unit, was used to explore biomarkers variations between periods following acute illness recovery and across different mediums like serum, plasma, blood cells, and brain tissues.

Sensitivity analyses were conducted using the leave-one-out approach to assess the robustness of the effect sizes. To detect publication bias, methods including the fail-safe N technique, the continuity-corrected Kendall tau, and Egger’s regression intercept were employed, with the latter two utilizing one-tailed p-values. In instances where Egger’s test suggested asymmetry, the trim-and-fill method was applied to estimate and incorporate missing studies, subsequently recalculating the adjusted effect sizes. Additionally, funnel plots, which plot study precision against standardized mean difference (SMD), were used. These plots not only show observed studies but also the imputed missing studies, aiding in the identification of small study effects.

## Results

### Search Outcomes

In our research, we employed specific keywords and mesh terms detailed in ESF-1, Table 3 to search through literature repositories such as PubMed, Google Scholar, and SciFinder. Our initial search yielded 3792 studies, as depicted in the PRISMA flow diagram (**Figure 1**), which delineates the study selection and exclusion process. Following a thorough screening to eliminate irrelevant and duplicate studies, we narrowed our focus to 221 articles. Out of these, only 87 met the strict criteria for inclusion in our systematic review. The exclusion of 130 articles was due to their non-compliance with our predefined standards. Furthermore, an additional 5 articles were excluded because of the lack of clear, extractable data. Consequently, a total of 82 studies were ultimately selected for the meta-analysis, adhering to our established inclusion and exclusion guidelines (Townsend, Dyer et al. 2020, Ahearn-Ford, Lunjani et al. 2021, Aparisi, Ybarra-Falcón et al. 2021, Colarusso, Maglio et al. 2021, Gameil, Marzouk et al. 2021, Loretelli, Abdelsalam et al. 2021, Montefusco, Ben Nasr et al. 2021, Ong, Fong et al. 2021, Patterson, Guevara-Coto et al. 2021, Peluso, Lu et al. 2021, Soni, Soni et al. 2021, Sun, Tang et al. 2021, Taha, Samaan et al. 2021, Utrero-Rico, González-Cuadrado et al. 2021, Wallis, Heiden et al. 2021, Wu, Deng et al. 2021, Acosta-Ampudia, Monsalve et al. 2022, Alfadda, Rafiullah et al. 2022, Aparisi, Ybarra-Falcón et al. 2022, Arslan, Aksakal et al. 2022, Cervia, Zurbuchen et al. 2022, Corrêa, Deus et al. 2022, Díaz-Salazar, Navas et al. 2022, Dugani, Mehta et al. 2022, Durstenfeld, Peluso et al. 2022, Elizabeth, Thomas et al. 2022, Fan, Wong et al. 2022, Fernández-de-Las-Peñas, Ryan-Murua et al. 2022, Ferrando, Dornbush et al. 2022, Flaskamp, Roubal et al. 2022, Fogarty, Ward et al. 2022, García-Abellán, Fernández et al. 2022, Johannes, Andrea et al. 2022, Kruger, Vlok et al. 2022, Littlefield, Watson et al. 2022, Magdy, Eid et al. 2022, Martone, Tosato et al. 2022, Meisinger, Goßlau et al. 2022, Meltendorf, Vogel et al. 2022, Nádasdi, Sinkovits et al. 2022, Patel, Knauer et al. 2022, Patra and Ray 2022, Peluso, Sans et al. 2022, Petrella, Nenna et al. 2022, Phetsouphanh, Darley et al. 2022, Queiroz, Neves et al. 2022, Saxena, Saxena et al. 2022, Schultheiß, Willscher et al. 2022, Sibila, Perea et al. 2022, Simone, Caitlin et al. 2022, Stavileci, Özdemir et al. 2022, Taeschler, Adamo et al. 2022, Wechsler, Butuci et al. 2022, Wiech, Chroscicki et al. 2022, Zhao, Schank et al. 2022, Al-Hakeim, Abed et al. 2023, Al-Hakeim, Al-Rubaye et al. 2023, Al-Hakeim, Al-Rubaye et al. 2023, Al-Hakeim, Al-Rubaye et al. 2023, Al-Hakeim, Khairi Abed et al. 2023, Allan-Blitz, Goodrich et al. 2023, Berentschot, Drexhage et al. 2023, Berezhnoy, Bissinger et al. 2023, Elke, Alvin et al. 2023, Garcia-Gasalla, Berman-Riu et al. 2023, Gomes, Brito et al. 2023, Julia, Stephan et al. 2023, Kelly, Sonya et al. 2023, Kuchler, Günthner et al. 2023, Melhorn, Alamoudi et al. 2023, Mitrović-Ajtić, Đikić et al. 2023, Neves, Quaresma et al. 2023, Park, Dean et al. 2023, Schultheiß, Willscher et al. 2023, Scott, Pearmain et al. 2023, Siekacz, Kumor-Kisielewska et al. 2023, Sommen, Havdal et al. 2023, Stepanova, Korol et al. 2023, Teng, Song et al. 2023, Torres-Ruiz, Lomelín-Gascón et al. 2023, Vazquez-Alejo, Tarancon-Diez et al. 2023, Xu, Zheng et al. 2023).

**Figure 1.**
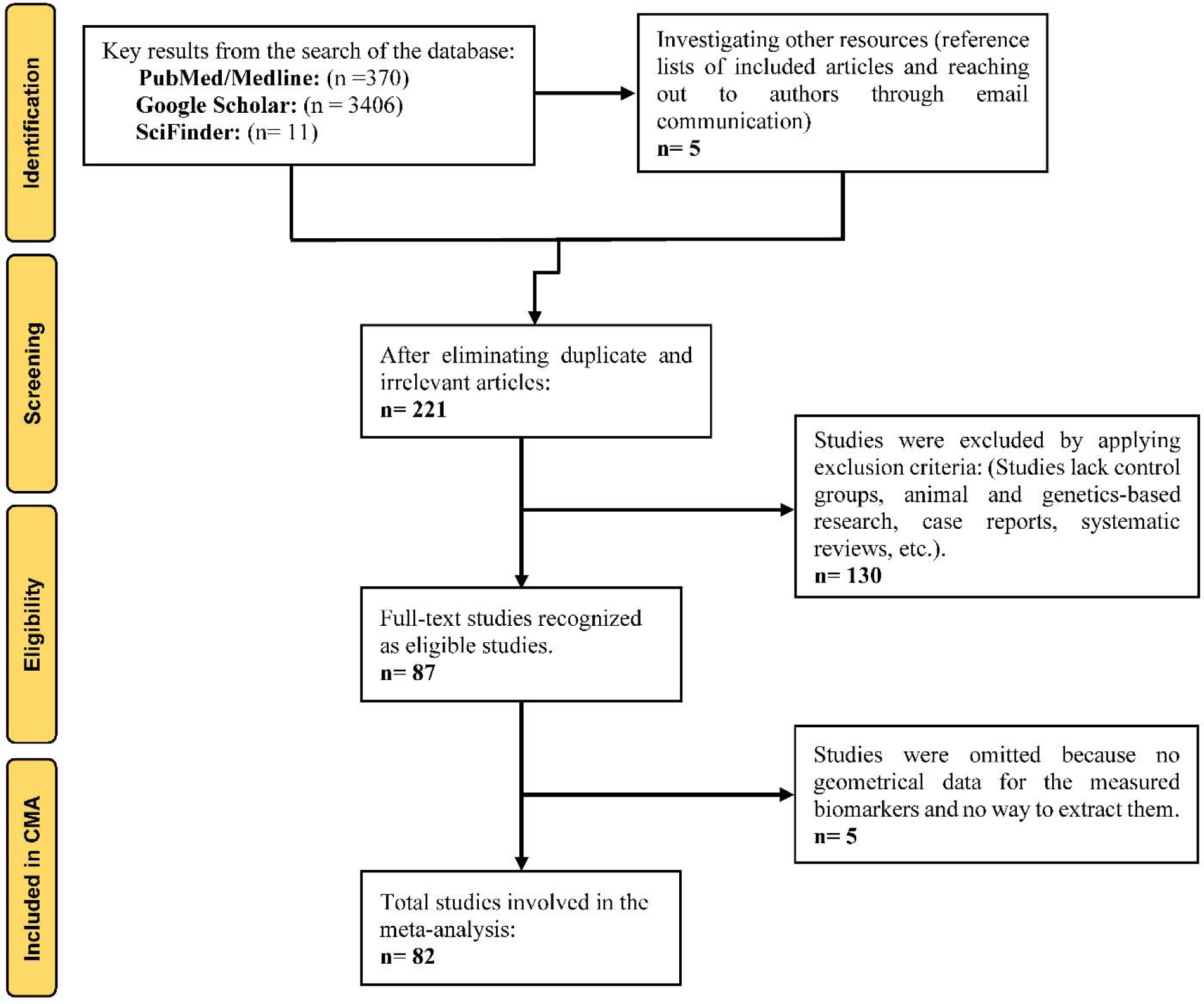
The PRISMA flow chart

Our meta-analysis synthesized data from 82 studies investigating immune biomarkers (cytokines/chemokines/growth factors and CRP) using various biological samples: 50 studies used serum, 31 utilized plasma, and 1 employed PMBC media. The total participant pool included 8373 individuals, comprising 4537 normal controls and 3836 patients with LC symptoms. The age range of participants spanned from 29 to 67 years. Diverse immunoassay techniques were employed to assess the inflammatory biomarkers, flow cytometry and multiplex immunoassay were the most common techniques as detailed in ESF-1, Table 6. The United States, Spain, and Germany were the leading contributors in this research domain, with 14, 9, and 8 studies, respectively. Other significant contributions came from Italy (6 studies), Iraq (5), Brazil (4), China, India, Ireland, and the UK (3 each). Egypt, the Netherlands, Singapore, South Africa, Switzerland, and Turkey each published 2 studies. As per ESF-1, Table 6, other countries also contributed one study each to this field. This meta-analysis evaluated the median values and minimum and maximum ranges for quality and redpoint scores. The median scores for quality and redpoint were determined to be 4,5 (ranging from 2,5 to 8,5) and 11,5 (ranging from 7,5 to 15), respectively, as presented in ESF-1, Table 6.

## Primary outcome variables

### IRS and CIRS in Long COVID patients versus normal controls

**Table 1** and **Figure 2** show the effect size of the IRS/CIRS ratio computed from 37 studies. None of these displayed CIs entirely below zero, but 6 had CIs entirely above zero. Additionally, 31 studies exhibited overlapping CIs with a mix of findings: 12 studies reported negative SMD values, whereas 19 had positive SMD values. **The forest plot (Figure 2)** indicates a significant increase in the IRS/CIRS ratio in LC patients compared to NC. Our bias analysis pointed to 3 studies missing on the right side of the forest plot, and adjusting the effect size to include these studies alters it to 0.156.

**Figure 2.**
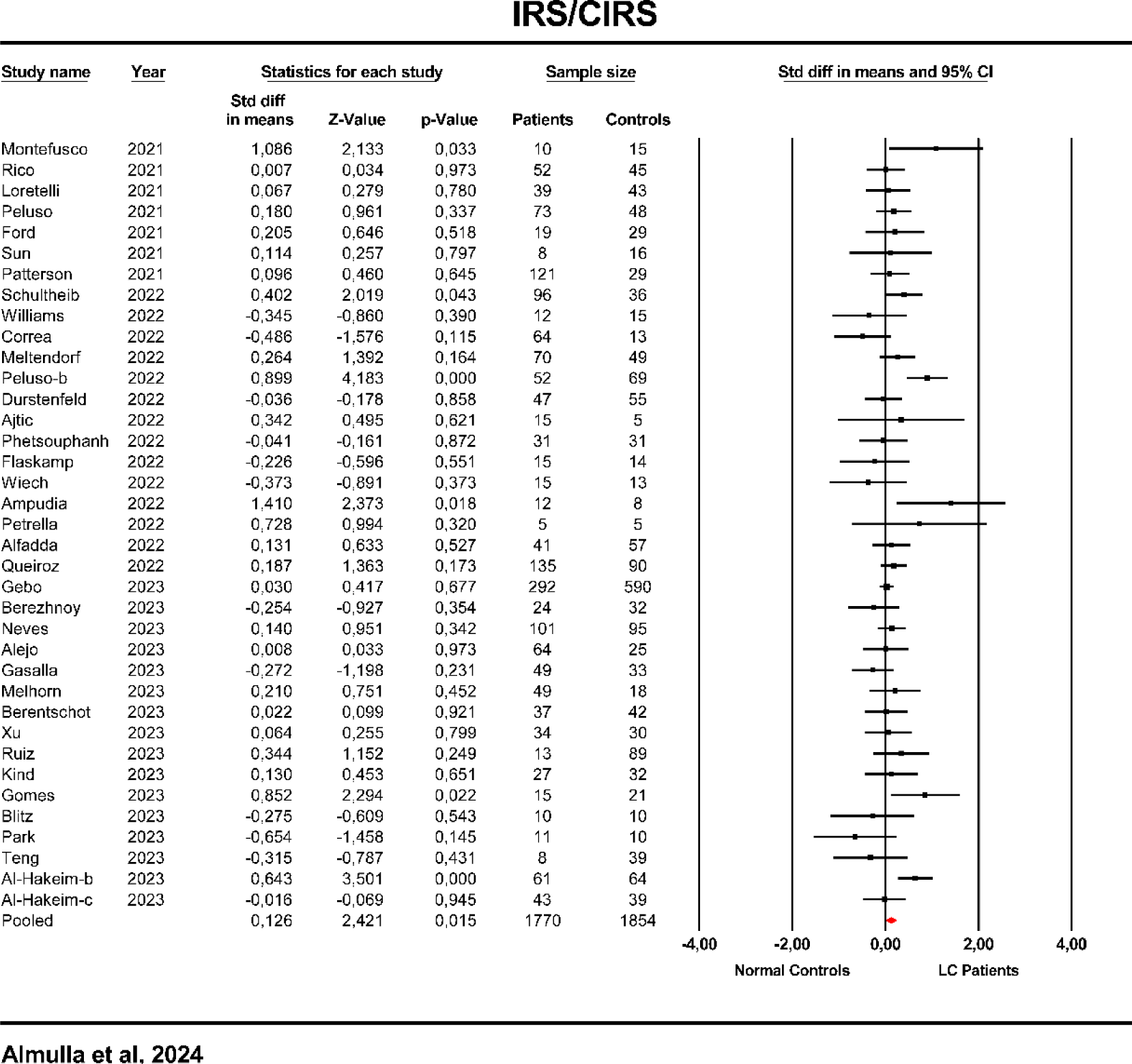
Forest plot of immune response system (IRS)/compensatory immune response system (CIRS) in patients with Long COVID (LC) and normal controls.

We included 62 studies to evaluate the IRS as displayed in **Table 1**. Of these, 2 studies had CIs completely below zero, while 18 showed CIs entirely above zero. A significant proportion, 42 studies, presented overlapping CIs with diverse outcomes: 19 studies noted negative SMD values, and 23 reported positive SMD values. **Table 2** and **Figure 3**, show that LC patients, compared to NC, demonstrated a significant increase in IRS. **Table 3** identified 7 studies missing on the right side of the forest plot. Including these missing studies increased the SMD value to 0.345.

**Figure 3.**
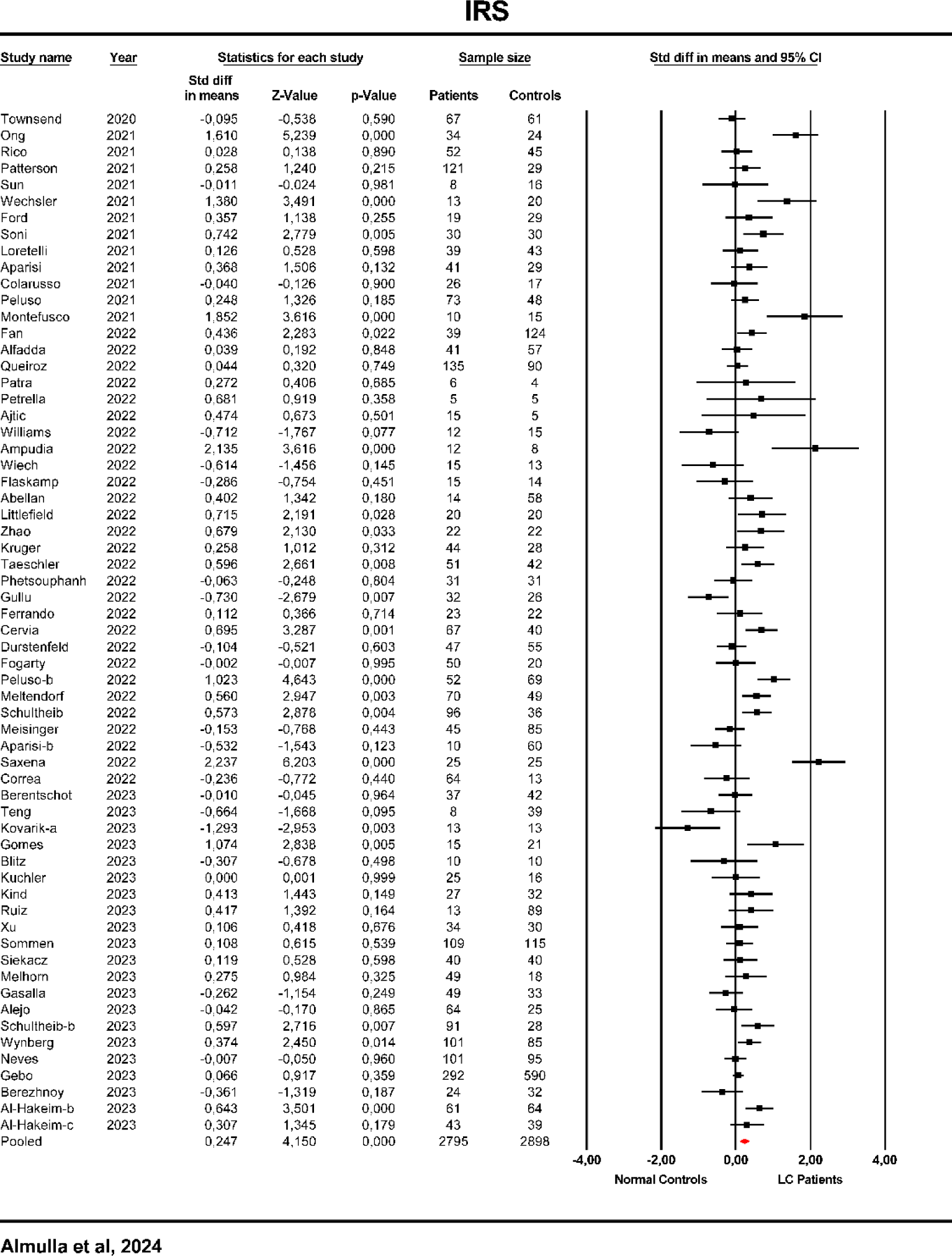
Forest plot of immune response system (IRS) in patients with Long COVID (LC) and normal controls.

In our research, the effect size for CIRS was derived from 36 studies, as detailed in **Table 1** and ESF-2, Figure 1. The analysis (**Table 2**) revealed no significant difference in CIRS levels between LC patients and NC. **Table 3** indicates no bias was found.

### Classical M1 and alternative M2 macrophages

The effect size for the M1/M2 macrophage ratio was calculated using data from 41 studies, as shown in **Table 1**. No significant M1/M2 ratio difference between LC patients and NC was found, as shown in **Table 2** and ESF-2, Figure 2. **Table 3** shows significant bias with 8 studies missing to the right of the forest plot. The inclusion of these missing studies adjusted the SMD value to 0.162, revealing a significant increase in the M1/M2 ratio.

We integrated data from 68 studies to assess M1 macrophage activation, as indicated in **Table 1**. Within these studies, 2 demonstrated CIs entirely below zero, and 22 had CIs completely above zero. A significant number 44 studies showed overlapping CIs with varied outcomes: 19 with negative SMD and 25 with positive SMD values. **Table 2** and **Figure 4** reveal that LC patients have significantly activated M1 macrophages compared to NC. Additionally, bias analysis uncovered 7 studies missing on the right side of the forest plot, and incorporating these studies increased the SMD value to 0.421. We analyzed M2 macrophage activation across 40 studies. The results, depicted in **Table 2** and ESF-2, Figure 3, indicated no significant changes in M2 macrophage status. The bias results in **Table 3**, found no evidence of bias.

**Figure 4.**
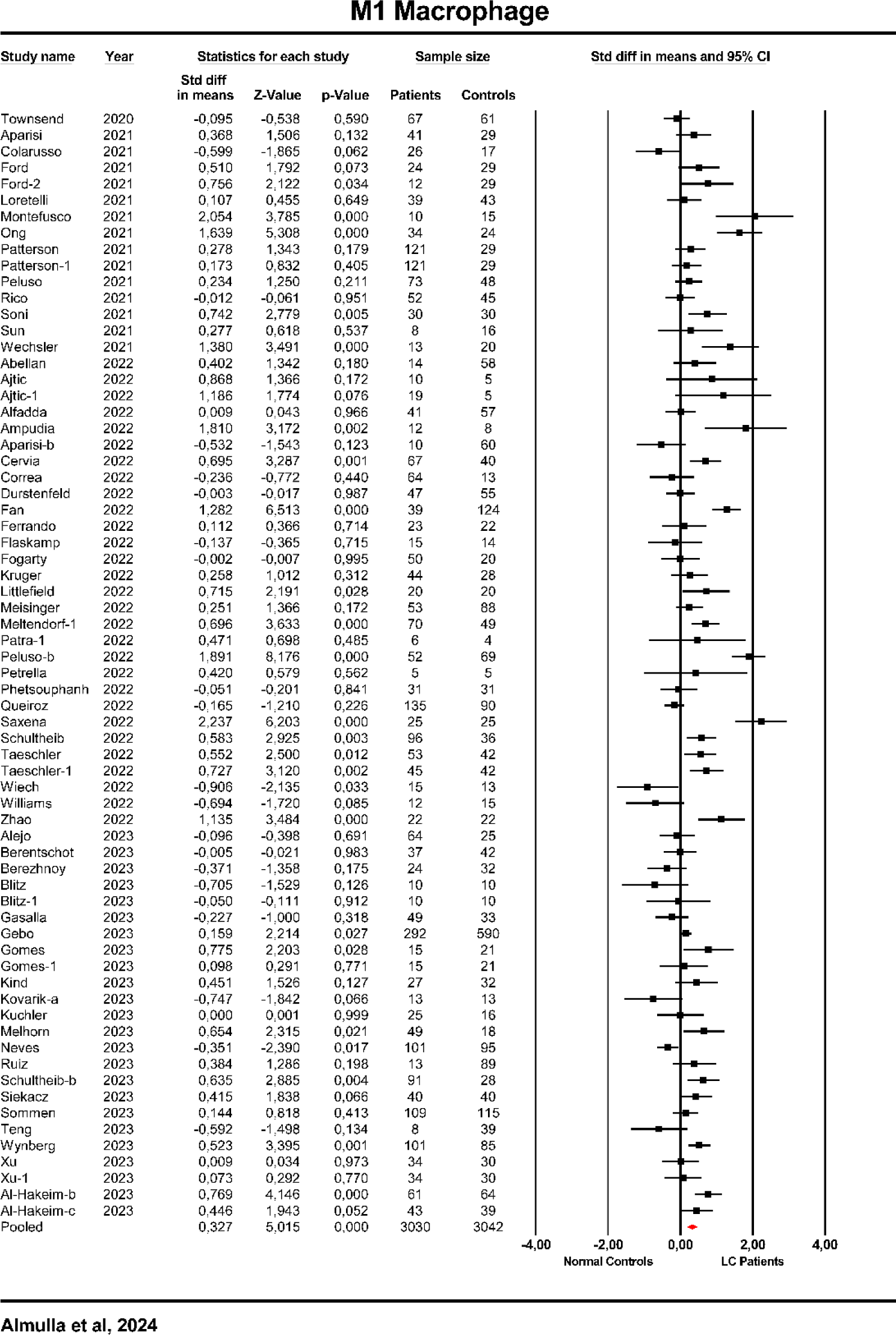
Forest plot of Macrophage M1 in patients with Long COVID (LC) and normal controls.

### Th1/ Th2 ratio, Th1, Th2 and Th17

Our study involved an analysis of 45 studies to assess the Th1/Th2 ratio, as detailed in **Table 1** and ESF-2, Figure 4. Initially, the findings indicated no significant change in the Th1/Th2 ratio among LC patients versus NC. However, as shown in **Table 3**, imputation of 12 missing studies significantly changed our results, yielding a modest yet significant effect size of 0.149.

We analyzed the effect size of Th1 using data from 45 studies as displayed in **Table 1**. Among these, 4 studies had CIs entirely below zero, while 11 showed CIs entirely above zero. Additionally, 30 studies demonstrated overlapping CIs, resulting in mixed findings: 14 reported negative SMD and 16 had positive SMD values. **Table 2 and Figure 5** show significantly increased Th1 activation in LC patients compared to NC. **Table 3** revealed that including 6 missing studies on the right side of the forest plot increased the effect size to 0.353.

**Figure 5.**
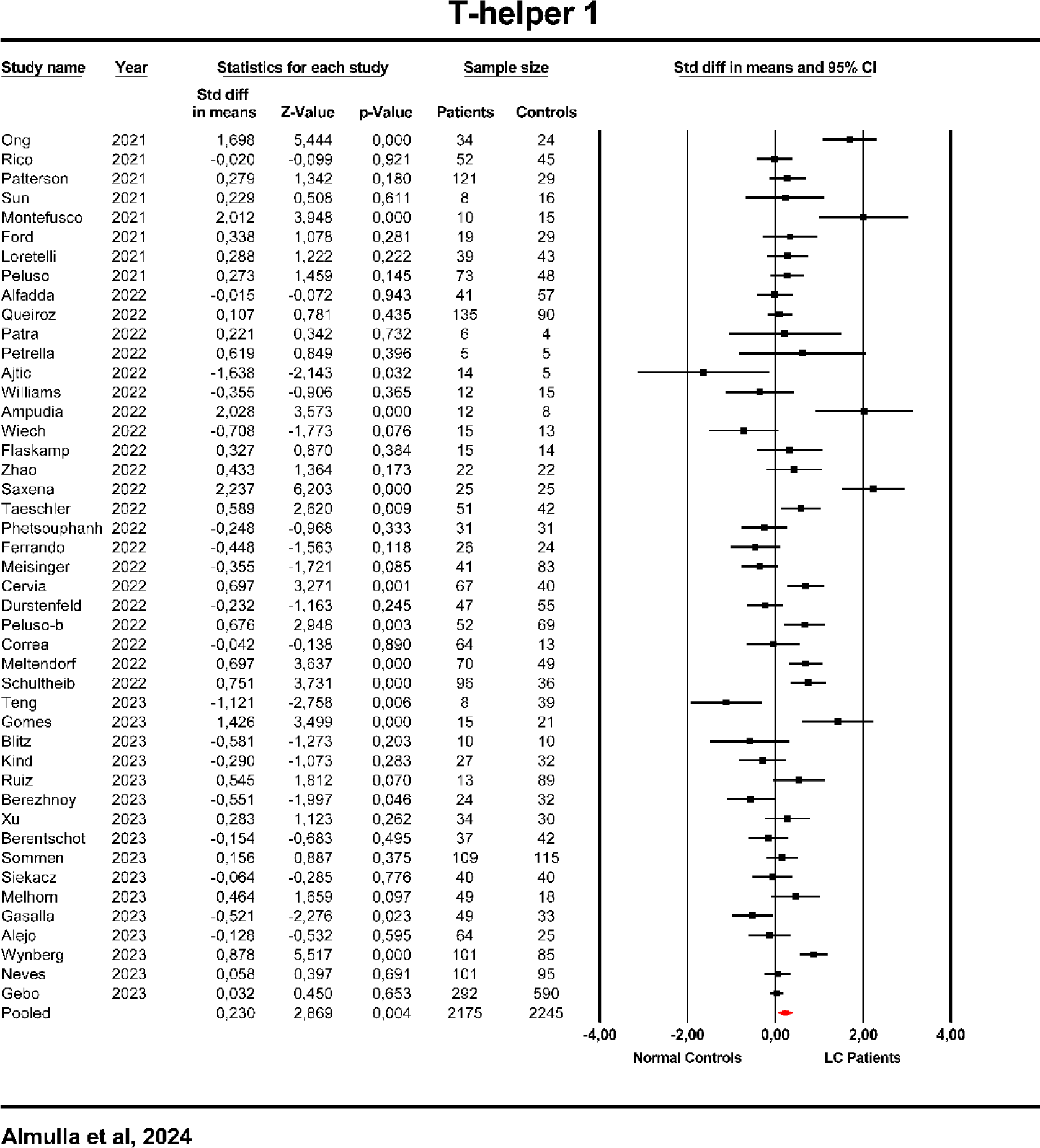
Forest plot of T helper (Th 1) in patients with Long COVID (LC) and normal controls.

The effect size of Th2 was assessed by analyzing 57 studies, as shown in **Table 1**. Of these studies, 4 displayed CIs entirely below zero, whereas 20 had CIs completely above zero. Furthermore, 33 studies exhibited overlapping CIs, leading to various outcomes: 19 studies with negative SMD and 14 with positive SMD values. **Table 2 and** ESF-2, Figure 5 indicate significant Th2 activation in LC patients compared to NC. Our bias analysis indicated no studies were missing.

**Table 1** shows that data from 55 studies was incorporated to assess Th17 levels. Within these studies, 3 had CIs entirely below zero, while 22 exhibited CIs completely above zero. Additionally, 31 studies with overlapping CIs yielded diverse results: 16 reported negative SMD, and 14 presented positive SMD values. **Table 2** and **Figure 6** show that LC patients displayed significantly activated Th17 compared to NC. Egger’s and Kendall’s tests (**Table 3**) detected 10 missing studies on the right side of the forest plot; imputing them would increase the SMD value to 0.492.

**Figure 6.**
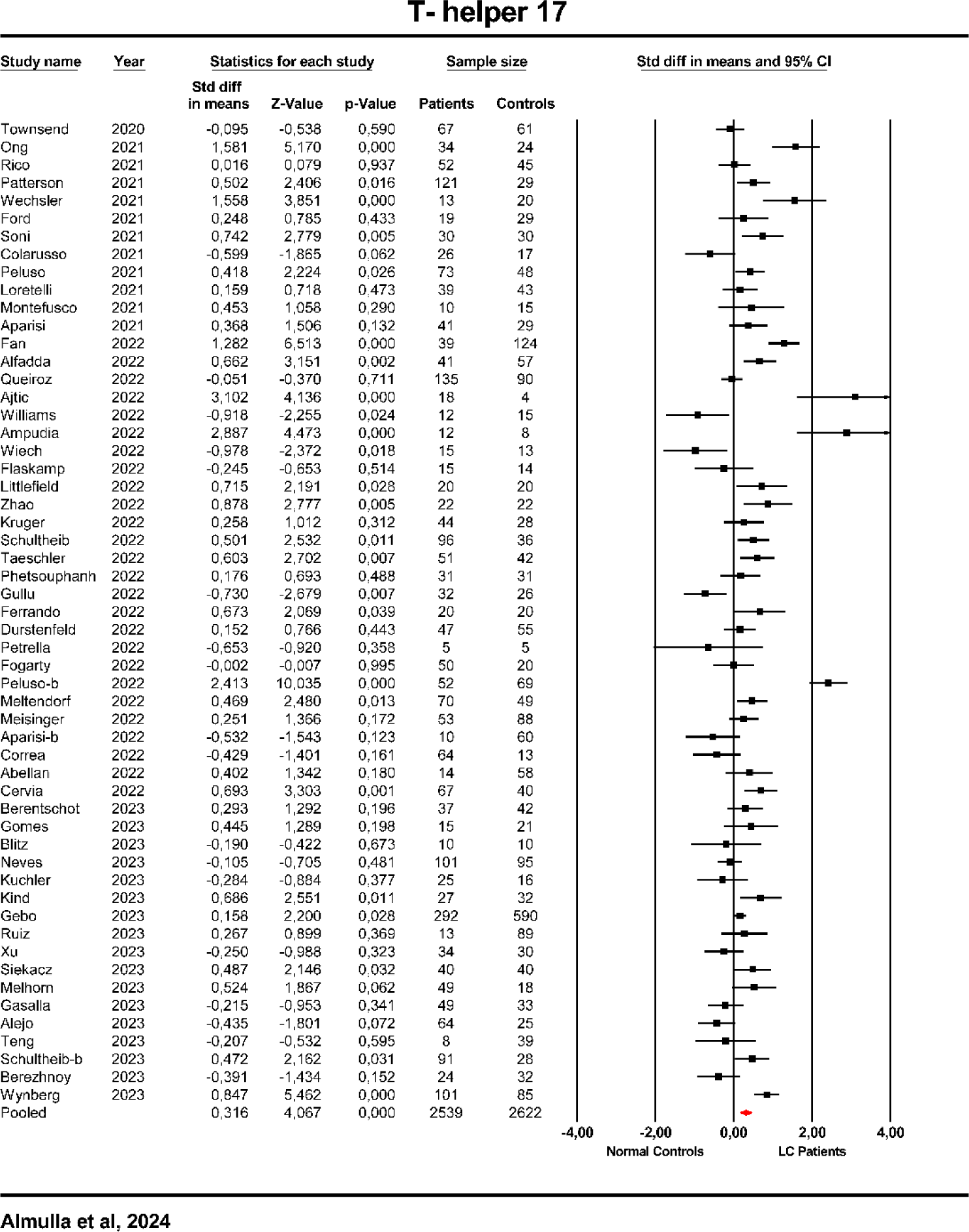
Forest plot of T helper (Th) 17 in patients with Long COVID (LC) and normal controls.

### Immune-related neurotoxicity

We assessed neurotoxicity by analyzing data from 62 studies, as detailed in **Table 1**. The findings varied across these studies: 8 had CIs entirely below zero, 22 entirely above zero, and 32 had overlapping CIs, resulting in 20 studies showing negative SMD and 12 showing positive SMD values. **Table 2 and Figure 7**, show that LC patients had significantly higher neurotoxicity index than the NC group. Table 3 identified 5 missing studies on the right side of the forest plot, and their inclusion increased the SMD value to 0.327.

**Figure 7.**
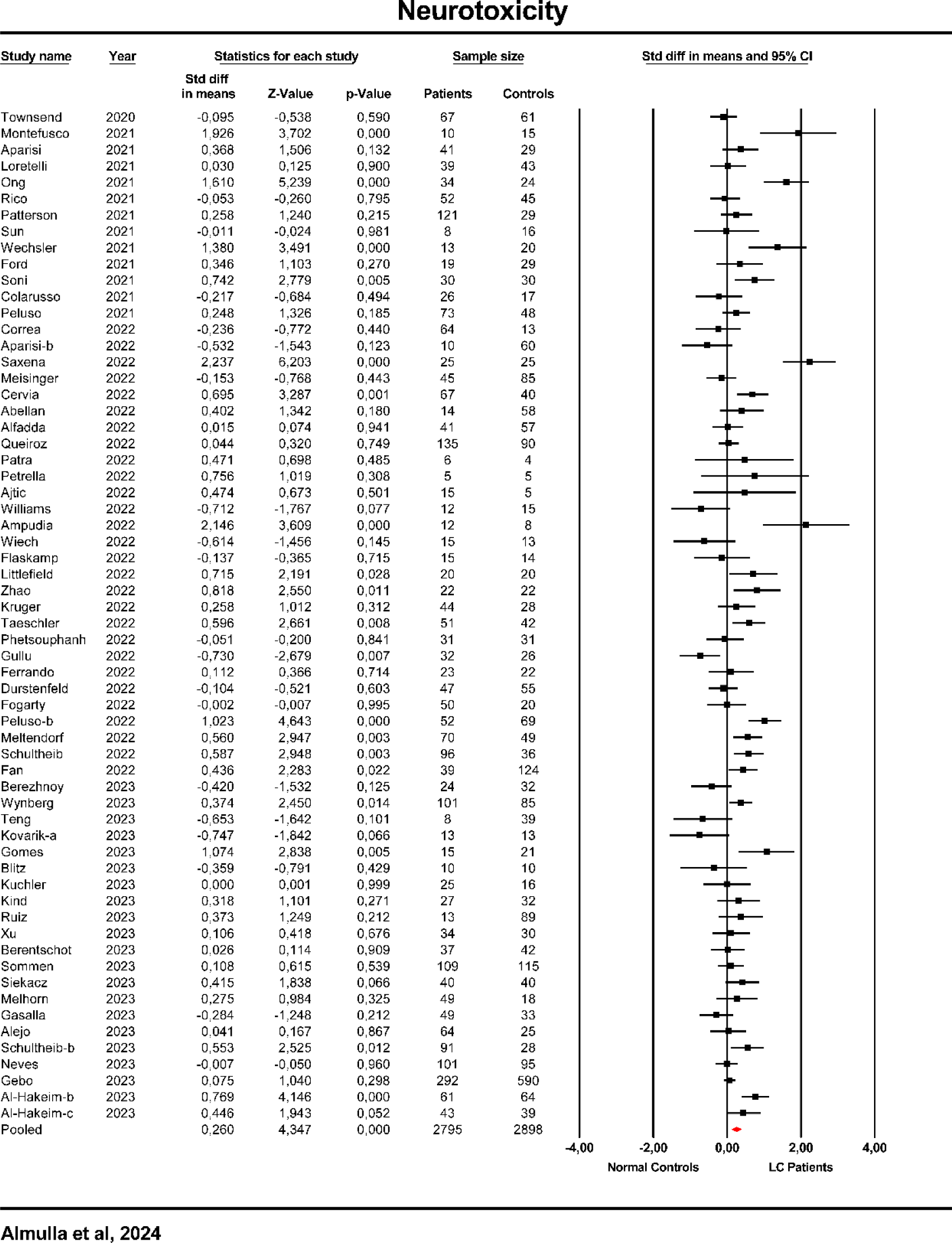
Forest plot of neurotoxicity in patients with Long COVID (LC) and normal controls.

## Secondary outcome variables

### CRP

**Table 1** displays that the CRP levels were analyzed using data from 38 studies. Of these, 2 studies had CIs entirely below zero, and 16 displayed CIs entirely above zero. A significant number of 20 studies presented overlapping CIs, leading to mixed results: 5 studies showed negative SMD, while 15 indicated positive SMD values. **Table 2 and** ESF-2 Figure 6 demonstrate a significant increase (with moderate effect size) in CRP levels in LC patients compared to the NC group. Bias analysis in **Table 3** identified 10 missing studies on the right side of the forest plot. Including these missing studies amplified the effect size to 0.776.

### IL-6

ESF-2, Table 1 indicates that IL-6 levels were evaluated across 52 studies. These studies showed a range of outcomes: 3 had CIs entirely below zero, 23 had CIs completely above zero, and 26 displayed overlapping CIs, resulting in 15 studies with negative SMD and 11 with positive SMD values. ESF-2, Table 2 and Figure 7, indicate that IL-6 levels significantly increased in LC patients compared to the NC group. ESF-2, Table 3 shows that including 7 missing studies on the right side of the forest plot raises the SMD value.

### IL-1β

Our study’s evaluation of IL-1β involved 23 studies, as shown in ESF-2, Table 1. These studies varied in their findings: 2 reported CIs entirely below zero, 8 entirely above zero, and 13 with overlapping CIs. This resulted in 6 studies showing negative SMD and 7 showing positive SMD values. ESF-2, Figure and Table 2 indicate that LC patients have significantly increased IL-1β levels compared to the NC group. ESF-2, Table 3 revealed that including 2 missing studies on the right side of the forest plot adjusts the SMD value to 0.484.

### TNF-α

The assessment of TNF-α levels involved the analysis of 41 studies, as detailed in ESF-2, Table 1 and Figure 9. The findings across these studies were diverse: 5 studies had CIs entirely below zero, 11 had CIs completely above zero, and 25 showed overlapping CIs, leading to 13 studies with negative SMD and 12 with positive SMD values. ESF-2, Table 2 reveals that LC patients have significantly higher TNF-α levels than the NC group. Egger’s and Kendall’s tests identified 8 missing studies on the right side of the forest plot, and adjusting the SMD value to these studies increased it.

### IL-2

The analysis of IL-2 levels was conducted using data from 17 studies, as shown in ESF-2, Table 1. The results from these studies varied: 2 showed CIs entirely below zero, 6 entirely above zero, and 9 had overlapping CIs. This diversity led to 5 studies reporting negative SMD and 4 reporting positive SMD values. ESF-2, Table 2 and Figure 10 reveal that IL-2 levels were significantly elevated in LC patients compared to the NC group. One missing study on the right side of the forest plot was identified, and implementing this study raises the SMD value.

### IL-22

We utilized data from four studies to assess IL-22 levels, as presented in ESF-1, Table 1. These studies showed varied results: none had CIs entirely below zero, two had CIs entirely above zero, and 2 had overlapping CIs. This resulted in 1 study indicating a negative SMD and another showing a positive SMD. ESF-2, Figure 11 and Table 2 show that LC patients exhibited significantly elevated IL-22 levels compared to the NC group. ESF-2, Table 3 highlights that incorporating 1 missing study on the right side of the forest plot amplifies the effect size to 0.482.

### IL-7

We assessed IL-7 levels using data from 12 studies, as indicated in ESF-2, Table 1. The results from these studies were diverse: 3 studies had CIs entirely below zero, 2 entirely above zero, and 7 exhibited overlapping CIs. Consequently, 3 studies reported negative SMD, while 4 showed positive SMD values. ESF-2, Table 2 and Figure 12 indicate no significant changes in IL-7 levels. Egger’s and Kendall’s tests identified 3 missing studies on the right side of the forest plot, and their inclusion led to significant results with a moderate effect size of 0.585.

### IP-10

The evaluation of IP-10 levels was conducted using data from 20 studies, as detailed in ESF-2, Table 1. These studies presented a range of outcomes: 1 had CIs entirely below zero, 4 had CIs completely above zero, and 15 showed overlapping CIs. As a result, 5 studies reported negative SMD, while 10 indicated positive SMD values. ESF-2, Table 2 and Figure 13 reveal significantly higher levels of IP-10 in LC patients compared to the NC group. ESF-2, Table 3 shows that including 1 missing study, identified on the right side of the forest plot, raised the SMD value.

### IL-8

Our research analyzed IL-8 levels by synthesizing data from 21 studies, as shown in ESF-2, Table 1. The findings varied, with 1 study having CIs entirely below zero, 8 entirely above zero, and 12 with overlapping CIs. This led to 8 studies indicating negative SMD, while 4 reported positive SMD values. ESF-2, Table 2 and Figure 14 indicate significantly elevated IL-8 levels in LC patients compared to NC. ESF-2, Table 3 shows that 2 missing studies were identified on the right side of the forest plot, which increased the SMD value after imputing them.

### VEGF

We assessed VEGF levels by integrating findings from 12 studies, as outlined in ESF-2, Table 1. The outcomes were diverse: none of the studies had CIs entirely below zero, 2 had CIs completely above zero, and 10 exhibited overlapping CIs. Consequently, this variability led to 4 studies showing negative SMD and 6 demonstrating positive SMD values. ESF-2, Table 2 and Figure 15 reveal that VEGF levels were significantly increased in LC patients versus the NC group. Incorporating two missing studies, identified on the right side of the forest plot, resulted in an increased SMD value.

### IL-13

Our analysis of IL-13 levels involved data from 11 studies, as detailed in ESF-2, Table 1. These studies showed considerable variation: no study reported CIs entirely below zero, 6 had CIs entirely above zero, and 5 presented overlapping CIs. This led to 1 study with a negative SMD and 4 with positive SMD values.ESF-2, Table 2 and Figure 16 indicate IL-13 levels significantly increased in LC patients versus NC. No bias was detected in these results.

### MIP-1α

We examined MIP-1α levels by analyzing data from 9 studies, as presented in ESF-2, Table 1. The outcomes of these studies showed significant variation: 1 study had CIs entirely below zero, 4 displayed CIs completely above zero, and 4 had overlapping CIs. This resulted in a mix of findings, with 2 studies reporting negative SMD and 2 indicating positive SMD values. ESF-2, Table 2 and Figure 17 reveal that MIP-1α levels were significantly increased in LC patients versus NC. Egger’s and Kendall’s tests found 1 missing on the right side of the forest plot, leading to increased effect size.

### IL-1Ra

IL-1Ra levels were examined using data from 8 studies, as outlined in ESF-2, Table 1 of our research. The findings from these studies varied, with none showing Cis entirely below zero, 4 with CIs completely above zero, and 4 displaying overlapping CIs. Consequently, all the included studies showed positive SMD values. ESF-2, Table 2 and Figure 18 indicate IL-1Ra levels were significantly increased (with a large effect size) in LC patients versus NC. These results were absent from bias.

### G-CSF

We evaluated G-CSF levels using data from 6 studies, as indicated in ESF-2, Table 1. These studies exhibited a range of outcomes: none had CIs entirely below zero, 3 had CIs completely above zero, and 3 showed overlapping CIs. Notably, all the studies resulted in positive SMD values. ESF-2, Table 2 and Figure 19 reveal that G-CSF levels were significantly increased in LC patients versus the NC group. ESF-2, Table 3 revealed 1 missing study on the right side of the forest plot, imputing it led to a huge SMD value of 1.027.

### VCAM-1

We analyzed VCAM-1 levels from 6 studies, as detailed in ESF-2, Table 1. The outcomes of these studies varied: none showed CIs entirely below zero, 1 had a CI completely above zero, and 5 had overlapping CIs. This resulted in 1 study with a negative SMD and 4 with positive SMD values. ESF-2, Table 2 and Figure 20 indicate that VCAM-1 levels were significantly increased in LC patients versus NC. Egger’s and Kendall’s tests revealed 3 missing studies on the left side of the forest plot, and including these studies reduced the effect size, rendering it no longer significant.

### EGF

EGF levels were examined by incorporating findings from 4 studies, as shown in ESF-2, Table 1. The results from these studies were diverse: none had CIs entirely below zero, 1 displayed a CI entirely above zero, and 3 exhibited overlapping CIs. This variability led to 1 study reporting a negative SMD and 2 indicating positive SMD values. ESF-2, Table 2 and Figure 21 indicate that EGF levels were significantly increased in LC patients versus NC. One missing study on the right side of the forest plot was detected, and imputing it raised the SMD value of 0.616.

### MCP-4

We examined MCP-4 levels using data from 3 studies, as outlined in ESF-2, Table 1. The study outcomes showed variation: none had CIs entirely below zero, 1 had a CI completely above zero, and 2 had overlapping CIs. All studies reported positive SMD values. ESF-2, Table 2 and Figure 22 reveal that MCP-4 levels were significantly increased in LC patients versus NC. Incorporating 1 missing study, identified on the right side of the forest plot and adjusted the SMD value to 0.607.

### MIP-3α

MIP-3α levels were assessed using data from 3 studies, as indicated in ESF-2, Table 1. These studies uniformly showed CIs entirely above zero. ESF-2, Table 2 and Figure 23 indicate that MIP-3α levels were significantly increased in LC patients versus NC. Including two missing studies on the left side of the forest plot, as shown in ESF-2, Table 3 led to an adjusted SMD value of 0.638.

### CXCL1

Our study’s examination of CXCL1 levels involved data from 3 studies, detailed in ESF-2, Table 1, which consistently displayed CIs entirely above zero. ESF-2, Table 2 and Figure 24 reveal that CXCL1 levels were significantly increased in LC patients versus NC. ESF-2, Table 3 indicates no bias was found.

### SCF

We analyzed SCF levels by synthesizing findings from 2 studies, as outlined in ESF-2, Table 1, where both consistently showed CIs entirely above zero. ESF-2, Table 2 and Figure 25 reveal that CXCL1 levels were significantly increased in LC patients versus NC.

This meta-analysis detected no significant differences between LC patients and NC in IL-10, MCP-1, MCP-3, CXCL11, CXCL9, Eotaxin, FGF, GM-CSF, HGF, ICAM, IFN-α, IFN-β, IFN-γ, IL-12, IL-15, IL-16, IL-17, IL-18, IL-1α, IL-21, IL-23, IL-27, IL-2Ra, IL-3, IL-33, IL-4, IL-5, IL-9, M-CSF, MIP-1β, MIP-3β, PDGF, PIGF, PECAM, RANTES, sRAGE, TGF-α, TGF-β, and TNF-β.

## Results of Meta-Regression

Meta-regression analysis was carried out to clarify the individual components contributing to the heterogeneity in research addressing immune-inflammatory profiles and isolated cytokines/chemokines/growth factors in LC patients. Our findings revealed that latitude and increasing number of hospitalized patients during the acute phase significantly impact most of the outcomes except Th2, TNF-α, IL-22, IL-7, IP-10, IL-8, IL-13, MIP-1α, G-CSF, and VCAM-1 as shown in ESF-1, Table 7. Age and the more than 6 months post-COVID period also significantly influence several outcomes. Other factors, including admission to the intensive care unit during the acute phase of illness, BMI and less than 3 months post-infection, and serum media, also show substantial contributions to the heterogeneity, as presented in ESF-1, Table 7.

## Discussion

This meta-analysis, which is currently the most extensive in its field, explores the impact of immune responses on LC disease. It offers new insights through an analysis of 82 studies related to LC, examining different immune profiles and an immune-related neurotoxicity index along with CRP and 58 individual levels of cytokines/chemokines/growth factors.

### LC is associated with activated IRS and normal CIRS pathways

The first key finding of the present meta-analysis reveals that LC is characterized by a significant increase in the IRS/CIRS ratio, although with a small effect size. The increase in this ratio is explained by activated IRS pathways, whereas the CIRS pathways were not altered in LC. These results indicate that activated immune-inflammatory pathways relative to the unchanged CIRS may play a significant role in the pathophysiology of LC disease. It is worth mentioning that this meta-analysis is the first to show upregulated IRS pathways in LC patients. However, recent meta-analyses displayed high levels of immune-inflammatory biomarkers in LC, including IL-6 (Yin, Agbana et al. 2023), CRP, and leukocytes (Yong, Halim et al. 2023).

In our study, the major constituents of IRS activation were IL-1α, IL-1β, IL-6, TNF-α, IL-12p70, IL-15, IL-16, IL-17, IL-18, CCL2, CCL3, CCL4, CCL5, CCL7, CCL11, CXCL1, CXCL8, CXCL9, CXCL10, IL-2, IFN-α, IFN-γ, TNF-β, GM-CSF, and G-CSF. We examined the solitary levels of these biomarkers, and the results show that only IL-1β, TNF-α, IP-10 (CXCL10), IL-6, IL-8, MIP-1α (CCL3), and CXCL1 were significantly elevated in LC patients compared to controls. In contrast, CIRS components include IL-4, IL-10, sIL-1RA, and sIL-2R, and our analysis shows that only IL-1Ra was significantly elevated in LC patients. This is important as increased sIL-RA levels indicate IRS and monocytic activation, although antagonizing IL-1 signaling (Maes and Carvalho 2018). These pathways are likely implicated in LC patients’ chronic fatigue and affective symptoms. Indeed, substantial evidence suggests a link between chronic fatigue, depression, and anxiety with the activation of abnormal IRS pathways (Maes, Ringel et al. 2013, Anderson, Berk et al. 2014, Maes and Carvalho 2018).

It should be noted that the current comprehensive evidence confirms our previous findings reporting that LC patients are suffering from activated immune-inflammatory responses (Al-Hakeim, Al-Rubaye et al. 2022, Al-Hakeim, Al-Rubaye et al. 2022, Al-Hakeim, Abed et al. 2023, Al-Hakeim, Al-Rubaye et al. 2023, Al-Hakeim, Khairi Abed et al. 2023, Almulla, Al-Hakeim et al. 2023, Almulla, Maes et al. 2023, Vojdani, Almulla et al. 2023). This augmentation of immune-inflammatory responses suggests a critical pathway that may contribute to the disease’s progression and the onset of neuro-psychiatric symptomatology. Consequently, targeting these immune-inflammatory responses could offer novel therapeutic avenues.

In LC disease, the persistence of immune-inflammatory reactions without activating the counter-regulatory immune system, as indicated in our study, may precipitate autoimmune responses in patients, potentially leading to more severe outcomes. Indeed, recent evidence, including findings by (Almulla, Maes et al. 2023, Vojdani, Almulla et al. 2023), supports this, showing that LC patients exhibit autoimmune reactions, as indicated by elevated levels of immunoglobulins (IgA, IgM, IgG) against self-proteins such as Activin A, myelin basic protein, myelin oligodendrocyte glycoprotein, and synapsin. Consequently, this state of neuro-autoimmunity, driven by persistently activated immune responses, can render COVID-19 survivors susceptible to neurological or mental symptoms (Strong 2023). The presence of heightened autoimmune markers is significantly linked to more advanced stages of LC, emphasizing the necessity of early identification and precise interventions for these patients.

### Increased cell-mediated immunity in LC disease

The second key finding of the present study is that LC disease is characterized by a notable rise in M1/M2 and Th1/Th2 ratios, albeit with small effect sizes that become apparent after accounting for missing studies. Furthermore, our investigation also shows a significant increase in the activation of Th1, Th2, Th17, and M1 in LC disease. Thus, the smaller alterations in the M1/M2 and Th1/Th2 ratios can be attributed to the heightened activity of these specific T lymphocytes and the macrophage M1 type. The findings imply that cell-mediated immunity may be a driving factor behind LC disease and its neuro-psychiatric symptoms. The results of our meta-analysis contrasts with those of Kovarik et al. (2023) who reported that LC disease involves a significant decrease in M1 activity and an increase in M2 activity, as indicated by cytokine secretions in their study (Kovarik, Bileck et al. 2023).

Various recent studies (Dan, Mateus et al. 2021, Peluso, Deitchman et al. 2021, Vibholm, Nielsen et al. 2021, Opsteen, Files et al. 2023) indicate a prolonged activation of SARS-CoV-2-specific T-cells, lasting up to six months post-infection. However, it’s noteworthy that many of these studies report a decreasing response of T cells over time (Dan, Mateus et al. 2021, Opsteen, Files et al. 2023). This observation points to the possible presence of persistent viral reservoirs, which might be responsible for the continued SARS-CoV-2-specific responses seen in LC patients. Indeed, we recently detected SARS-CoV-2 persistence in LC patients, triggering ongoing immune responses (Vojdani, Almulla et al. 2023). It has been hypothesized that ongoing immune activation and the consequent neuroinflammation could play a role in the neurocognitive symptoms of LC, a theory supported by the observation of heightened inflammatory markers in the cerebrospinal fluid (CSF) of patients exhibiting these symptoms (Apple, Oddi et al. 2022).

In LC patients compared to uninfected individuals, there is a significant elevation in plasma levels of cytokines and chemokines, particularly those related to macrophage response to Type 2 cytokines, including IL-5, IL-9, IL-17F, IL-18, IL-22, IL-23, IL-33, CCL2/MCP-1, and sCD163 (Schultheiß, Willscher et al. 2023). Further research highlighted significant changes in post-COVID-19 patients, especially in T-cell dynamics. Severe LC cases often show a pronounced shift toward an exhausted or senescent phenotype in CD4+ and CD8+ T cells, with noticeable disturbances in CD4+ T regulatory cells, indicating the intricate nature of immune responses in LC disease (Wiech, Chroscicki et al. 2022, Berentschot, Drexhage et al. 2023). Additionally, Ruiz et al. (2023) found a significant increase in CD57 expression on CD8+ T cells, suggesting a shift towards a pro-inflammatory state that could contribute to the persistence of LC symptoms (Torres-Ruiz, Lomelín-Gascón et al. 2023). These findings suggest that low-grade persistent inflammatory responses in LC might be predominantly driven by cell-mediated immunity.

### CRP and the acute phase response in LC

The present study’s third key finding reveals that patients with LC exhibit significantly elevated CRP levels compared to the control group with a large effect size. This observation aligns with and builds upon the findings of previous meta-analysis (Yong, Halim et al. 2023) and research (Wallis, Heiden et al. 2021, Al-Hakeim, Al-Rubaye et al. 2022, Littlefield, Watson et al. 2022, Al-Hakeim, Al-Rubaye et al. 2023, Al-Hakeim, Khairi Abed et al. 2023), which consistently report elevated CRP levels in LC patients, suggesting a pronounced acute phase response in this group. In contrast to the previous meta-analysis by (Yong, Halim et al. 2023), which synthesized effect sizes from only 10 studies, the current study encompasses 38 studies, thereby enhancing the precision in the calculation of the effect sizes. Importantly, we previously detected that peak body temperature during the acute COVID stage could significantly predict CRP in LC patients (Al-Hakeim, Al-Rubaye et al. 2022).

An acute phase response is a hallmark of LC, where inflammatory cytokines like interleukin-6 (IL-6) prompt the liver to increase CRP production (Almulla, Al-Hakeim et al. 2023). Other conditions in LC, including autoimmunity (Almulla, Maes et al. 2023), gut dysbiosis (Zhang, Zhou et al. 2023), and thrombosis (Dasari, Banga et al. 2022) may also trigger the production of CRP. CRP plays a role in compromising the integrity of the blood-brain barrier (BBB), through its action in enhancing vascular permeability, stimulating microglia, increasing oxidative stress, and enabling the infiltration of inflammatory cells and cytokines into the central nervous system, possibly resulting in neuroinflammation and neurodegeneration (Kuhlmann, Librizzi et al. 2009, Closhen, Bender et al. 2010, Hsuchou, Kastin et al. 2012). Overall, high CRP levels in LC patients are probably partly responsible for the manifestations of CFS and affective symptoms. Indeed, we recently detected that CRP is one of the best predictors for neuropsychiatric symptoms in LC disease (Al-Hakeim, Khairi Abed et al. 2023, Almulla, Maes et al. 2023, Vojdani, Almulla et al. 2023).

### Immune-associate neurotoxicity

Another key finding of our study is the observation of increased immune-associated neurotoxicity in LC disease, calculated by aggregating neuro-immunotoxic mediators including IL-1β, IL-6, TNF-α, IL-2, IFN-γ, IL-12p70, IL-16, IL-17, CCL2, CCL3, CCL5, CCL11, CXCL1, CXCL8, CXCL10, GM-CSF, and M-CSF. This builds on our previous discovery that LC patients exhibit heightened immune-associated neurotoxicity, as measured by a composite based on the sum of inflammatory, insulin resistance, and oxidative stress biomarkers, which associates with a reduced quality of life in these patients (Al-Hakeim, Al-Naqeeb et al. 2023, Almulla, Ali Abbas Abo et al. 2023a). These studies have consistently found a significant association between chronic fatigue, affective symptoms (major LC symptoms), and immune-associated neurotoxicity. Further research indicates that certain products of M1 (e.g., IL-1β, IL-6, IL-15, TNF-α, CCL2, CCL5, CXCL10), Th1 (like IL-2, IFN-γ, IL-16), Th17 (such as IL-6 and IL-17), and even Th2 (for instance, IL-4), can impair neuronal functions related to mood regulation, leading to neuro-affective toxicity and potentially resulting in depression (Maes, Rachayon et al. 2022).

Those findings suggest that LC disease may be regarded as a complex exogenous neuro-psychiatric disorder with relative nosological specificity (Di Nicola and Stoyanov 2021). In that theoretical framework, exogenous neurotoxic neuro-psychiatric conditions are caused by environmental agents which realize pathogenic effects based on relatively specific pathways and the clinical manifestations associated with them.

### Heterogeneity issues

Our meta-regression analysis reveals a significant inverse association between age and various inflammatory biomarkers, including IRS/CIRS, neurotoxicity, M1/M2, Th17, and CRP. This suggests a more pronounced inflammatory state in younger COVID-19 survivors. Such findings align with the concept of immunosenescence, which involves the gradual weakening of the immune system with aging, leading to alterations in immune responses like cytokine production variability (Ventura, Casciaro et al. 2017). Additionally, the onset of LC after more than 6 months shows a significant direct correlation with regulatory immune responses, including CIRS, M2 macrophage, and Th2, while an early onset (less than 3 months) is associated with Th17 and IL-17. Furthermore, IRS, neurotoxicity, M1, Th1/Th2, Th1, Th17, CRP, IL-1β, and IL-2 display an inverse relationship with the latitude of the countries where patients reside. These results suggest that the proinflammatory state of LC is more pronounced in patients living closer to the equator. Other important factors influencing these biomarkers are detailed in the ESF-1 Table 7.

## Limitations

Despite the novel insights offered by this meta-analysis, it is important to acknowledge certain limitations that may affect the interpretation of its findings. The findings of this study would have been further enriched if we had the opportunity to investigate the interplay between neuro-oxidative and neuroimmune-inflammatory pathways in the context of LC. Due to the lack of medication-related information in most of the studies we analyzed, this study did not assess the impact of medications. Therefore, future research should include details on treatments administered during the acute phase or in LC cases. Our study did not examine the effects of COVID-19 vaccinations on inflammatory biomarkers due to the absence of relevant data in the studies we included. Future research should, therefore, include information on vaccination status. Our study could not conduct meta-analyses for CXCL12, IL-29, IL-31, MCP-2, MCP-3, TNFR2, TNFSF9, TNFSF10, TNFSF12, TNFSF14, and sVEGFR, because only one study was available.

## Conclusion

This comprehensive and large-scale meta-analysis, the first of its scale incorporating 82 studies, indicates that LC patients typically exhibit heightened immune-inflammation, enhanced cell-mediated immunity, and immune-related neurotoxicity. These findings open up potential new treatment avenues targeting these specific pathways for LC management.

## Declaration of Competing Interests

The authors affirm that no conflicts of interest are associated with this manuscript.

## Ethical approval and consent to participate

Not applicable.

## Consent for publication

Not applicable.

## Availability of data and materials

Upon full utilization by all authors, the last author, MM, will address legitimate inquiries for the dataset used in this meta-analysis, which will be shared in an Excel file format.

## Funding

The study was funded by the C2F program, Chulalongkorn University, Thailand, No. 64.310/436/2565 to AFA, an FF66 grant and a Sompoch Endowment Fund (Faculty of Medicine), MDCU (RA66/016) to MM, and Grant № BG-RRP-2.004-0007-С01 „Strategic Research and Innovation Program for the Development of MU - PLOVDIV– (SRIPD-MUP)“, Creation of a network of research higher schools, National plan for recovery and sustainability, European Union – NextGenerationEU.

## Author’s contributions

AA and MM were responsible for designing the present study, while YT and AA collected the data. YT and AA independently verified the data accuracy. AA and MM conducted the statistical analysis. All authors have actively participated in the drafting and revising of the manuscript and have provided their approval for the submission of the final version.

## Acknowledgments

Not applicable.

## Notes

### Competing Interest Statement

The authors have declared no competing interest.

