## supplementary file for "Immune activation and immune-associated neurotoxicity in Long-COVID: A systematic review and meta-analysis of 82 studies comprising 58 cytokines/chemokines/growth factors"

SHORT TITLE: Immune response system in Long COVID disease

1. Sichuan Provincial Center for Mental Health, Sichuan Provincial People’s Hospital, School of Medicine, University of Electronic Science and Technology of China, Chengdu 610072, China

2. Key Laboratory of Psychosomatic Medicine, Chinese Academy of Medical Sciences, Chengdu, 610072, China

3. Department of Psychiatry, Faculty of Medicine, Chulalongkorn University, Bangkok, Thailand.

4. Medical Laboratory Technology Department, College of Medical Technology, The Islamic University, Najaf, Iraq.

5. Immunosciences Lab, Inc., Los Angeles, CA 90035, USA.

6. Cyrex Laboratories, LLC, Phoenix, AZ 85034, USA.

7. Cognitive Impairment and Dementia Research Unit, Faculty of Medicine, Chulalongkorn University, Bangkok, Thailand.

8. Cognitive Fitness and Biopsychological Technology Research Unit, Faculty of Medicine. Chulalongkorn University, Bangkok, 10330, Thailand, Bangkok, 10330, Thailand.

9. Department of Psychiatry, Medical University of Plovdiv, Plovdiv, Bulgaria.

10. Research Institute, Medical University of Plovdiv, Plovdiv, Bulgaria.

11. Strategic Research and Innovation Program for the Development of MU - PLOVDIV–(SRIPD-MUP), European Union – NextGenerationEU.

12. Kyung Hee University, 26 Kyungheedae-ro, Dongdaemun-gu, Seoul 02447, Korea.

* **Corresponding author:**

Prof. Dr. Michael Maes, M.D., Ph.D.

Sichuan Provincial Center for Mental Health

Sichuan Provincial People’s Hospital,

School of Medicine,

University of Electronic Science and Technology of China

Chengdu 610072

China

e-mail addresses:

<https://scholar.google.co.th/citations?user=1wzMZ7UAAAAJ&hl=th&oi=ao>

Highly cited author: 2003-2023 (ISI, Clarivate)

ScholarGPS: Worldwide #1 in molecular neuroscience; #1/4 in pathophysiology

Expert worldwide medical expertise ranking, Expertscape (December 2022), worldwide:

#1 in CFS, #1 in oxidative stress, #1 in encephalomyelitis, #1 in nitrosative stress, #1 in nitrosation, #1 in tryptophan, #1 in aromatic amino acids, #1 in stress (physiological), #1 in neuroimmune; #2 in bacterial translocation; #3 in inflammation, #4-5: in depression, fatigue and psychiatry.

**ESF, Table 1**. Overview of the cytokines, chemokines and growth factors measured in the current study.

| Protein abbreviations | Gene Symbol | Protein name / alias |
| --- | --- | --- |
| IFN-γ | **IFNG** | Interferon-γ |
| IL-1α | **IL1A** | Interleukin-1α |
| IL-1β | **IL1B** | Interleukin-1β |
| sIL-1RA | **IL1RN** | Soluble interleukin-1 receptor antagonist |
| IL-2 | **IL2** | Interleukin-2 |
| IL-2Ra | **IL2RA** | Interleukin-2 receptor |
| IL-3 | **IL3** | Interleukin-3 |
| IL-4 | **IL4** | Interleukin-4 |
| IL-5 | **IL5** | Interleukin-5 |
| IL-6 | **IL6** | Interleukin-6 |
| IL-7 | **IL7** | Interleukin-7 |
| IL-9 | **IL9** | Interleukin-9 |
| IL-10 | **IL10** | Interleukin-10 |
| IL-13 | **IL13** | Interleukin-13 |
| IL-15 | **IL15** | Interleukin-15 |
| IL-16 | **IL16** | Interleukin-16 |
| IL-17 | **IL17A** | Interleukin-17 |
| IL-18 | **IL18** | Interleukin-18 |
| TNF-α | **TNF** | Tumor necrosis factor-α |
| TNF-β | **LTA** | Tumor necrosis factor-β or lymphotoxin-alpha (LT-α) |
| G-CSF | **CSF3** | Granulocyte colony stimulating factor (G-CSF) or colony stimulating factor 3 (CSF3) |
| M-CSF | **CSF1** | Macrophage colony-stimulating factor (M-CSF) or colony stimulating factor 1 (CSF1) |
| GM-CSF | **CSF2** | Granulocyte-macrophage colony-stimulating factor (GM-CSF) or colony-stimulating factor 2 (CSF2) |
| CCL2 or MCP1 | **CCL2** | C-C motif chemokine ligand 2 (CCL2) or monocyte chemoattractant protein 1 (MCP1) |
| CCL3 or MIP-1α | **CCL3** | C-C motif Chemokine ligand 3 (CCL3) or macrophage inflammatory protein 1-alpha (MIP-1α) |
| CCL4 or MIP-1β | **CCL4** | C-C motif chemokine ligand 4 (CCL4) or macrophage inflammatory protein 1β (MIP-1β) or lymphocyte activation gene 1 protein |
| CCL5 or RANTES | **CCL5** | C-C motif chemokine ligand 5 (CCL5) or regulated upon activation, normally T-expressed, and presumably Secreted (RANTES) |
| CCL7 or MCP3 | **CCL7** | C-C motif chemokine ligand 7 (CCL7) or monocyte-chemotactic protein 3 (MCP3). |
| CCL11 or Eotaxin | **CCL11** | C-C motif chemokine ligand 11 (CCL11) or eosinophil chemotactic protein |
| CXCL1 or GRO-α | **CXCL1** | C-X-C motif chemokine 1 (CXCL1) or growth-regulated alpha protein (GRO) |
| CXCL8 or IL-8 | **CXCL8** | C-X-C motif chemokine ligand 8 (CXCL8) or interleukin-8 (IL-8) |
| CXCL9 or MIG | **CXCL9** | C-X-C motif chemokine ligand 9 (CXCL9) or monokine induced by gamma interferon (MIG) |
| CXCL10 or IP10 | **CXCL10** | C-X-C motif chemokine ligand 10 (CXCL10) or Interferon gamma-induced protein 10 (IP10) |
| FGF | **FGF2** | Fibroblast growth factor 2 (FGF) or basic fibroblast growth factor |
| HGF or SF | **HGF** | Hepatocyte growth factor (HGF) or scatter factor (SF) |
| PDGF | **PDGFA** | Platelet derived growth factor (PDGF) |
| SCF | **KITLG** | Stem cell factor (SCF) or Kit ligand (KITLG) |
| VEGF | **VEGFA** | Vascular endothelial growth factor |
| IL-22 | **IL22** | Interleukin-22 |
| VCAM1 | **VCAM1** | vascular cell adhesion molecule-1 |
| EGF | **EGF** | Epidermal growth factor |
| CCL13 or MCP-4 | **CCL13** | Monocyte Chemoattractant Protein-4 |
| CCL20 or MIP-3α | **CCL20** | Macrophage Inflammatory Protein-3 alpha |
| CXCL11 | **CXCL11** | Chemokine (C-X-C motif) ligand 11 |
| ICAM-1 | **ICAM1** | Intercellular Adhesion Molecul |
| IFN-α | **IFNA** | Interferon alpha |
| IFN-β | **IFNB1** | Interferon beta |
| IL-12 | **IL-12A gene encodes the p35 subunit. IL-12B gene encodes the p40 subunit.** | Interleukin-12 |
| IL-21 | **IL21** | Interleukin-21 |
| IL-23 | **IL12B gene encodes the p40 subunit. IL23A gene encodes the p19 subunit.** | Interleukin-23 |
| IL-27 | **IL27** | Interleukin-27 |
| IL-33 | **IL33** | Interleukin-33 |
| CCL19 or MIP-3β | **CCL19** | Macrophage Inflammatory Protein-3 beta |
| PECAM-1 | **PECAM1** | Platelet Endothelial Cell Adhesion Molecule-1 |
| sRAGE | **AGER** | soluble form of the Receptor for Advanced Glycation End-products |
| TGF-α | **TGFA** | Transforming Growth Factor alpha |
| TGF-β | **TGFB** | Transforming Growth Factor beta |
| PIGF | **PGF** | Placental Growth Factor |

**ESF, Table 2**. Composite scores used in the present study.

| Immune Profile | Members |
| --- | --- |
| M1 macrophage | IL-1β, IL-6, TNF-α, IL-12p70, IL-15, CCL2, CCL5, CXCL1, CXCL8, CXCL9, CXCL10 |
| M2 macrophages | IL-10, IL-4, IL-13, VEGF, PDGF, sIL-1RA |
| z M1 – z M2 | zM1 – zM2 |
| T helper (Th)-1 | IL-2, sIL-2R, IFN-a, IFN-γ, IL-12p70, IL-16, TNF-α, TNF-β |
| Th-2 | IL-4, IL-5, IL-9, IL-13, IL-10, IL-6 |
| z Th- z Th-2 | zTh-1 – zTh-2 |
| Th-17 | IL-6, IL-17 |
| IRS | IL-1α, IL-1β, IL-6, TNF-α, IL-12p70, IL-15, IL-16, IL-17, IL-18, CCL2, CCL3, CCL4, CCL5, CCL7, CCL11, CXCL1, CXCL8, CXCL9, CXCL10, IL-2, IFN-α, IFN-γ, TNF-α, TNF-β, TRAIL, GM-CSF, M-CSF, G-CSF, SCGF |
| CIRS | IL-4, IL-10, sIL-1RA, sIL-2R |
| z IRS – z CIRS | z IRS – z CIRS |
| Neurotoxicity | IL-1α, IL-1β, IL-6, TNF-α, TRAIL, IL-2, IFN-γ, IL-12p70, IL-16, IL-17, CCL2, CCL3, CCL5, CCL11, CXCL1, CXCL8, CXCL10 |

IRS: immune-inflammatory response system; CIRS: compensatory immunoregulatory system

Adapted from:

*Maes M, Rachayon M, Jirakran K, Sodsai P, Klinchanhom S, Gałecki P, Sughondhabirom A, Basta-Kaim A. The Immune Profile of Major Dysmood Disorder: Proof of Concept and Mechanism Using the Precision Nomothetic Psychiatry Approach. Cells. 2022 Mar 31;11(7):1183. doi: 10.3390/cells11071183. PMID: 35406747; PMCID: PMC8997660.*

*Kalayasiri R, Dadwat K, Supaksorn T, Sirivichayakul S, Maes M. Methamphetamine (MA) use, MA dependence, and MA-induced psychosis are associated with increasing aberrations in the compensatory immunoregulatory system and interleukin-1α and CCL5 levels. medRxiv 2023.03.26.23287766; doi: https://doi.org/10.1101/2023.03.26.23287766*

**ESF, Table 3.** Search sentences and terms were used in each database.

| **Database Name** | **Search Sentence** | **No. of Articles** |
| --- | --- | --- |
| **PubMed/Medline** | ("Long COVID" [Title/Abstract] OR "Post-Acute Sequelae of COVID-19" [Title/Abstract]) AND ("Inflammation Mediators" [MeSH Terms] OR "C-Reactive Protein" [MeSH Terms] OR "Cytokines" [MeSH Terms] OR "Chemokines" [MeSH Terms]) | **108** |
|  | ("Long COVID" [Title/Abstract] OR "Post-Acute Sequelae of COVID-19" [Title/Abstract]) AND ("Acute-Phase Proteins" [MeSH Terms] OR "Interleukins" [MeSH Terms] OR "Interleukin-1beta" [MeSH Terms] OR "IL-1beta" [Title/Abstract] OR "Interleukin-1" [MeSH Terms] OR "IL-alpha" [Title/Abstract] OR "Interleukin-6" [MeSH Terms] OR "IL-6" [Title/Abstract] OR "Interleukin-2" [MeSH Terms] OR "IL-2" [Title/Abstract] OR "Interleukin-10" [MeSH Terms] OR "IL-10" [Title/Abstract] OR "Tumor Necrosis Factor" [MeSH Terms] OR "TNF" [Title/Abstract] OR "Chemokines" [MeSH Terms]) | **139** |
|  | ("Post COVID-19 Syndrome" [Title/Abstract] OR "Post-Acute Sequelae of COVID-19" [Title/Abstract] OR "Post COVID" [Title/Abstract]) AND ("Acute-Phase Proteins" [MeSH Terms] OR "Interferon-gamma" [MeSH Terms] OR "IFN-gamma" [Title/Abstract] OR "Interferon-alpha" [MeSH Terms] OR "IFN-alpha" [Title/Abstract] OR "Vascular Endothelial Growth Factors" [MeSH Terms] OR "VEGF" [Title/Abstract] OR "Interleukins" [MeSH Terms] OR "IL-7" [Title/Abstract] OR "IL-8" [Title/Abstract] OR "IL-16" [Title/Abstract] OR "IL-22" [Title/Abstract] OR "IL-9" [Title/Abstract] OR "IL-12" [Title/Abstract] OR "IL-5" [Title/Abstract] OR "Monocyte Chemoattractant Proteins" [MeSH Terms] OR "MCP" [Title/Abstract] OR "Eotaxin" [Title/Abstract]) | **123** |
| **Google Scholar** | ("Post COVID" OR "Post COVID-19 Syndrome" OR "Post-Acute Sequelae of COVID-19") AND ("inflammatory mediators") AND ("acute phase proteins" OR "IFN-gamma" OR "Interferon gamma" OR "IFN-alpha" OR "Interferon alpha" OR "VEGF" OR "Vascular Endothelial Growth Factor" OR "IL-7" OR "Interleukin-7" OR "IL-8" OR "Interleukin-8" OR "MCP" OR "Monocyte Chemoattractant Protein" OR "Eotaxin" OR "IL-16" OR "Interleukin-16" OR "IL-22" OR "Interleukin-22" OR "IL-9" OR "Interleukin-9" OR "IL-12" OR "Interleukin-12" OR "IL-5" OR "Interleukin-5"). | **991** |
|  | ("Long COVID" OR "Post-Acute Sequelae of COVID-19" OR "Post-COVID-19 Syndrome") AND ("inflammatory mediators") AND ("C-reactive protein" OR "CRP") AND ("cytokines") AND ("chemokines") | **445** |
|  | ("Long COVID" OR "Post-Acute Sequelae of COVID-19" OR "Post-COVID-19 Syndrome") AND ("inflammatory mediators") AND ("acute phase proteins" OR "IL-1beta" OR "Interleukin-1beta" OR "IL-alpha" OR "Interleukin-alpha" OR "TNF" OR "Tumor Necrosis Factor" OR "IL-6" OR "Interleukin-6" OR "IL-2" OR "Interleukin-2" OR "IL-10" OR "Interleukin-10" OR "chemokines") | **1970** |
| **SciFinder** | ("Long COVID") AND "C-reactive protein" AND "cytokines" AND "chemokines" | **11** |

**ESF, Table 4.** Immune cofounder’s scale (ICS) applied from Andrés-Rodríguez, et al., 2019

| **Methodological quality of the study** | |
| --- | --- |
| **1** | Study sample ≥ 128 participants including patients and HC (1= Yes, 0 = No) |
| **2** | Did the study control results for potential confounders (e,g, age, BMI, gender, race)? (1= Yes, 0 = No) |
| **3** | Were participants with Long COVID and HCs age- and-gender-matched or statistically controlled? (1= Yes, 0 = No) |
| **4** | Was the time of sample collection specified (e,g, morning vs, evening)? (1= Yes, 0 = No) |
| **5** | Were participants with Long COVID free of immunomodulatory drugs including anti-cytokines, corticoids, immunoglobulins, and immunosuppressants, been through a medication washout or the intake was statistically controlled? (1= Yes, 0 = No) |
| **6** | Were participants with Long COVID free of antidepressants and mood stabilizers or statistically controlled? (1= Yes, 0 = No) |
| **7** | Reporting of either the manufacturer of the test or its parameters (detection limit and coefficient of variation) (1= Yes, 0 = No) |
| **8** | Reporting how data under detection limit was handled (1 = Yes, 0 = No) |
| **9** | Reporting % of the sample under detection limit (1=Yes, 0= No) |
| **10** | Reporting blood fraction (serum, plasma, culture supernatant or whole blood) (1= Yes, 0 = No) |
| **Total quality score (10 points)** | |
| **Biomarker confounders red points**  *The red points should not be given if the item is statistically controlled for* | |
| **1** | 3 red points for comorbid illnesses such as autoimmune disorders & other immune disorders including RA, psoriasis, IBD, COPD, MS |
| **2** | **3 red points** for use of recreational drugs such as methamphetamine or opioids (Not applicable if psychiatric disorders are excluded) |
| **3** | **2 red points** for comorbidity with Long COVID (any psychiatric or mood disorder) |
| **4** | **2 red points** when groups were not matched for age |
| **5** | **2 red points** for sex |
| **6** | **2 red points** for medication use as for example immunomodulators |
| **7** | **2 red points** for early traumatic life events |
| **8** | **2 red points** for shift work and primary sleep disorders |
| **9** | 1,5 red points for antidepressants or antipsychotics |
| **10** | 1 red point for other neuro-psychiatric comorbidities, as for example schizophrenia, autism, GAD, PTSD |
| **11** | 1 red point for more common systemic immune disorders including diabetes type 1/2, essential hypertension, metabolic syndrome |
| **12** | **1 red point** for not fasting (8 hours before blood extraction) |
| **13** | **1 red point** for use of omega-3 and antioxidant supplements |
| **14** | **1 red point** for BMI |
| **15** | **1 red point** for physical activity or sedentary life |
| **16** | **1 red point** for smoking |
| **17** | **1 red point** for use of oral contraceptives or NSAIDs |
| **18** | 0,5 red points for ethnicity in countries such as US, Brazil (not China or Japan) |
|  | **Total red point score (26 points)** |

**ESF, Table 5.** PRISMA checklist

| **Section/topic** | **#** | **Checklist item** | **Reported on page #** |
| --- | --- | --- | --- |
| **TITLE** | | | |
| Title | 1 | Identify the report as a systematic review, meta-analysis, or both. | 1 |
| **ABSTRACT** | | | |
| Structured summary | 2 | Provide a structured summary including, as applicable: background; objectives; data sources; study eligibility criteria, participants, and interventions; study appraisal and synthesis methods; results; limitations; conclusions and implications of key findings; systematic review registration number. | 3 |
| **INTRODUCTION** | | | |
| Rationale | 3 | Describe the rationale for the review in the context of what is already known. | 6,7 |
| Objectives | 4 | Provide an explicit statement of questions being addressed with reference to participants, interventions, comparisons, outcomes, and study design (PICOS). | 7 |
| **METHODS** | | | |
| Protocol and registration | 5 | Indicate if a review protocol exists, if and where it can be accessed (e.g., Web address), and, if available, provide registration information including registration number. | 8 |
| Eligibility criteria | 6 | Specify study characteristics (e.g., PICOS, length of follow-up) and report characteristics (e.g., years considered, language, publication status) used as criteria for eligibility, giving rationale. | 8,9 |
| Information sources | 7 | Describe all information sources (e.g., databases with dates of coverage, contact with study authors to identify additional studies) in the search and date last searched. | 8, ESF, Table 3 |
| Search | 8 | Present full electronic search strategy for at least one database, including any limits used, such that it could be repeated. | 10 |
| Study selection | 9 | State the process for selecting studies (i.e., screening, eligibility, included in systematic review, and, if applicable, included in the meta-analysis). | 10 |
| Data collection process | 10 | Describe method of data extraction from reports (e.g., piloted forms, independently, in duplicate) and any processes for obtaining and confirming data from investigators. | 10 |
| Data items | 11 | List and define all variables for which data were sought (e.g., PICOS, funding sources) and any assumptions and simplifications made. | 11 |
| Risk of bias in individual studies | 12 | Describe methods used for assessing risk of bias of individual studies (including specification of whether this was done at the study or outcome level), and how this information is to be used in any data synthesis. | 11,12 |
| Summary measures | 13 | State the principal summary measures (e.g., risk ratio, difference in means). | 11,12 |
| Synthesis of results | 14 | Describe the methods of handling data and combining results of studies, if done, including measures of consistency (e.g., I^2^) for each meta-analysis. | 11,12,13 |
| Risk of bias across studies | 15 | Specify any assessment of risk of bias that may affect the cumulative evidence (e.g., publication bias, selective reporting within studies). | 11,12 |
| Additional analyses | 16 | Describe methods of additional analyses (e.g., sensitivity or subgroup analyses, meta-regression), if done, indicating which were pre-specified. | 11,13 |
| **RESULTS** | | |  |
| Study selection | 17 | Give numbers of studies screened, assessed for eligibility, and included in the review, with reasons for exclusions at each stage, ideally with a flow diagram. | 13 |
| Study characteristics | 18 | For each study, present characteristics for which data were extracted (e.g., study size, PICOS, follow-up period) and provide the citations. | 13, ESF, Table 4 |
| Risk of bias within studies | 19 | Present data on risk of bias of each study and, if available, any outcome level assessment (see item 12). | Table 3 |
| Results of individual studies | 20 | For all outcomes considered (benefits or harms), present, for each study: (a) simple summary data for each intervention group (b) effect estimates and confidence intervals, ideally with a forest plot. | Table 1 |
| Synthesis of results | 21 | Present results of each meta-analysis done, including confidence intervals and measures of consistency. | Table 2 |
| Risk of bias across studies | 22 | Present results of any assessment of risk of bias across studies (see Item 15). | Table 3 |
| Additional analysis | 23 | Give results of additional analyses, if done (e.g., sensitivity or subgroup analyses, meta-regression [see Item 16]). | ESF, Table 5 |
| **DISCUSSION** | | |  |
| Summary of evidence | 24 | Summarize the main findings including the strength of evidence for each main outcome; consider their relevance to key groups (e.g., healthcare providers, users, and policy makers). | 27-33 |
| Limitations | 25 | Discuss limitations at study and outcome level (e.g., risk of bias), and at review-level (e.g., incomplete retrieval of identified research, reporting bias). | 33 |
| Conclusions | 26 | Provide a general interpretation of the results in the context of other evidence, and implications for future research. | 34 |
| **FUNDING** | | |  |
| Funding | 27 | Describe sources of funding for the systematic review and other support (e.g., supply of data); role of funders for the systematic review. | 35 |

**ESF, table 6.** Characteristics of the studies included in the systematic reviews and meta-analysis

| **NO** | **Authors, years** | **Setting** | **Post-COVID period** | **Type of case** | **Type of Control** | **Sample Size** | | | **Age** | | **Assessed Biomarkers and Findings** | **Specimen** | **Method** | **Quality score** | **Red point score** |
| --- | --- | --- | --- | --- | --- | --- | --- | --- | --- | --- | --- | --- | --- | --- | --- |
|  |  |  |  |  |  | **Cases**  **M/F** | **Control**  **M/F** | **Total**  **M/F** | **Case-Mean (SD) or Median (IQR)** | **Control- Mean(SD) or Median (IQR)** |  |  |  |  |  |
| 1 | (Teng, Song et al. 2023) | China | 3-6 months | Change in renal functions | No change in renal functions | 8 3/5 | 39 27/12 | 47 30/17 | 42 (15,18) | 49,1 (14,36) | TGF-β*, IL-1β*, IL-2*, IL-4*, IL-5*, IL-6*, IL-8#,IL-10*,IL-12p70*, IL-17A*, IL-17F#, IL-22#, TNF-α*, TNF-β*, IFN-γ* | Serum | Flow Cytometry | 5 | 10,5 |
| 2 | (Berentschot, Drexhage et al. 2023) | Netherlands | 3-6 months | Long COVID | Healthy control | 37 24/13 | 42 26/16 | 79 50/29 | 58 (55.0-66.0) | 62 (51.8-68.3) | BDNF#,CCL2#,CCL7#, CXCL10#,CXCL9#,GM-CSF*,IFN-β#,IFN-γ#,IL-10#,IL-6,IL-7#,TNF-α# | Serum | Luminex assay | 8 | 11,5 |
| 3 | (Kuchler, Günthner et al. 2023) | Germany | 13.8 months | ME/CFS | No ME/CFS | 25 6/19 | 16 4/12 | 41 10/31 | 40,6 (12,2) | 44,7 (12,2) | CXCL10#, IL-6#, IL-8*, MCP1* | Serum | Flow Cytometry | 5 | 10,5 |
| 4 | (Neves, Quaresma et al. 2023) | Brazil | 3-6 months | Long COVID | Healthy control | 202 77/125 | 95 34/61 | 297 111/186 | 46,5 (38–54) | 39 (34–45) | IFN-γ*, IFN-γ_1*, IL-10*, IL-10_1*,IL-17*, IL-17_1*, IL-2*, IL-2_1*, IL-4#, IL-4_1#, IL-6#, IL-6_1#, TNF-α*, TNF-α_1* | Serum | Flow Cytometry | 6 | 12 |
| 5 | (Elke, Alvin et al. 2023) | Netherlands | 5-6 months | Long COVID | Healthy control | 101 45/56 | 85 24/61 | 186 69/117 | 55 | 48 | CRP*, CRP_1*, IL-6*, IL-6_1*, IP-10#, IP-10_1*, MCP1#, MCP1_1#, TNF-alpha*, TNF-alpha_1* | Serum | Luminex assay | 6 | 11,5 |
| 6 | (Patel, Knauer et al. 2022) | Canada | 3-6 months | Long COVID | Healthy control | 23 13/10 | Unknown | Unknown | 61(15) | Unknown | PECAM*, VEGF*, ICAM*, VEGF*, VEGF*, VCAM* VEGF* | Plasma | Luminex assay | 5,5 | 13,5 |
| 7 | (Ferrando, Dornbush et al. 2022) | USA | 6-8 months | Post-COVID | No Post-COVID | 32 7/25 | 28 12/16 | 60 19/41 | 48,1 (12.8) | 33.7 (11.0) | CRP*, IL-6*, TNF-α# | Serum | Unknown | 4,5 | 11,5 |
| 8 | (Ahearn-Ford, Lunjani et al. 2021) | Ireland | 3-6 months | Post-COVID | Healthy control | 24 13/11 | 29 15/14 | 53 28/25 | 53,5 | 43.2 | CRP*, CRP_1*, Eotaxin*, Eotaxin_1#, Eotaxin_2*, Eotaxin_3#, FGF*, GM-CSF_2*, GM-CSF_3*, ICAM-1*, ICAM-1_2*, IFN-γ*, IFN-γ_2#, IL-10*, IL-10_1*, IL-12/23p40* , IL-12/23p40_1*, IL-12/p70*, IL-12/p70_1*, IL-13*, IL-13_1*, IL-15*, IL-15_1*, IL-16*, IL-16_1*, IL-17*, IL-17_1*, IL-17_2#, IL-17_3*, IL-17_5*, IL-17_6#, IL-17_7#, IL-17_8*, IL-1α*, IL-1α_1*, IL-1β*, IL-1β_1*, IL-1Ra*, IL-1Ra_1*, IL-2*, IL-2_1*, IL-21*, IL-21_1*, IL-22*, IL-22_1*, IL-23*, IL-23_1*, IL-27#, IL-27_1#, IL-3*, IL-3_1#, IL-31* , IL-31_1*, IL-4*, IL-4_1*, IL-5*, IL-5_1*, IL-6*, IL-6_1*, IL-7#, IL-7_1*, IL-8*, IL-8_1*, IL-9*, IL-9_1*, IP-10*, IP-10_1*, MCP-1*, MCP-1_1*, MCP-4*, MCP-4_1*, MIP-1α*, MIP-1α_1*, MIP-1β*, MIP-1β_1*, MIP-3α*, MIP-3α_1*, PIGF*, TNF-α*, TNF-α_1*, TNF-β*, TNF-β_1*, VCAM-1*, VCAM-1_1*, VEGF*, VEGF_2*, VEGF_3* | Serum | Multiplex assay | 4,5 | 11,5 |
| 9 | (Julia, Stephan et al. 2023) | Germany | 3 months | Post-COVID | Healthy control | 27 9/18 | 32 15/17 | 59 24/35 | 37 | 36 | CXCL1*, EGF*, Eotaxin#, IFN-γ*, IL-10*, IL-12/p70#, IL-13*, IL-18*, IL-1α*, IL-1β*, IL-1Ra*, IL-4*, IL-5*, IL-6*, IL-7*, MCP-1#, MIP-3β*, PDGF*, RANTES*, TGF-α*, TNF-α*, VEGF* | Plasma | Luminex assay | 3,5 | 11,5 |
| 10 | (Elizabeth, Thomas et al. 2022) | USA | 3 months | Post-COVID | Healthy control | 12 0/12 | 15 7/8 | 27 7/20 | Unknown | Unknown | IL-1β#, IL-2#, sIL-2R#, IL-4*, IL-5*, IL-6*, IL-8*, IL-10*, IL-12*, IL-13#, IL-17*, TNF-α* | plasma | Multiplex assay | 2,5 | 12 |
| 11 | (Fogarty, Ward et al. 2022) | Ireland | 1.5 months | Long COVID | Healthy control | 50 30/20 | Unknown | 50 30/20 | 50 (36–63) | Unknown | IL-6* | Plasma | Elisa | 3 | 12,5 |
| 12 | (Schultheiß, Willscher et al. 2022) | Germany | 8-10 months | Post-COVID | Healthy control | 91 26/65 | 28 11/17 | 119 37/82 | 51 | 50 | IL-17*, IL-18*, IL-22*, IL-23*, IL-33*, IL-5*, IL-9*, MCP-1* | Plasma | Flow Cytometry | 4,5 | 11,5 |
| 13 | (Scott, Pearmain et al. 2023) | UK | 9 months | long COVID | Healthy control | 68 43/25 | 70 44/26 | 138 87/51 | 59,1 (1.5) | 57.4 (1.6) | CRP* | Serum | Unknown | 5,5 | 10,5 |
|  |  |  |  |  |  | 63 40/23 | 73 46/27 | 136 86/50 | 58,1 (1.5) | 58.5 (1.6) | CRP_1* | Serum |  |  |  |
| 14 | (Vazquez-Alejo, Tarancon-Diez et al. 2023) | Spain | 6 months | Long COVID | Healthy control | 64 41/23 | Unknown | 64 41/23 | 57 (25–98) | Unknown | CXCL1*, Eotaxin*, IFN-α*, IFN-γ*, IL-10*, IL-12/p70*, IL-17*, IL-18*, IL-1β*, IL-23*, IL-33*, IL-6*, IL-8*, IL-8_1*, IP-10*, MCP-1*, MCP-1_1*, MIG*, MIP-1α*, MIP-1β*, MIP-3α*, TNF-α* | Plasma | Flow Cytometry | 3,5 | 13,5 |
| 15 | (Soni, Soni et al. 2021) | India | 1 month | Post-COVID | Healthy control | 30 21/9 | 30 20/10 | 60 41/19 | 53,9 (16.1) | 46.2 (15.1) | CRP*, IL-6* | Serum | Unknown | 3,5 | 12,5 |
| 16 | (Utrero-Rico, González-Cuadrado et al. 2021) | Spain | 6 months | Post-COVID | Healthy control | Unknown | Unknown | Unknown | 65 (50-74) | 29 (23) | CXCL9*, G-CSF*, GM-CSF#, IFN-α*, IFN-γ*, IL-10#, IL-17*, IL-1α*, IL-1β*, IL-1Ra*, IL-2#, IL-21*, IL-6#, IL-7#, IL-8#, IP-10*, MCP-1#, MIP-1α* TNF-α#, VEGF# | Plasma | Luminex assay | 6,5 | 13,5 |
| 17 | (Kelly, Sonya et al. 2023) | USA | 3 months | Post-COVID | No post-COVID | 292 87/205 | 590 289/301 | 882 376/506 | Unknown | Unknown | IFN-α#, IFN-γ*, IL-10*, IL-15*, IL-16#, IL-17#, IL-18#, IL-1Ra*, IL-21#, IL-22#, IL-29#, IL-2Ra*, IL-6*, IL-7*, IL-8*, IP-10*, MCP-1*, MIP-1α#, TNF-α#, VCAM-1#, VEGF* | Plasma | Multiplex assay | 7,5 | 12,5 |
| 18 | (Xu, Zheng et al. 2023) | China | 10-13 months | Post-COVID | Healthy control | 34 16/18 | 30 12/18 | 64 28/36 | 59 (39.0–67.2) | 52.0 (46.3–57.7) | IL-10*, IL-10_1*, IL-2*, IL-2_1*, IL-4*, IL-4_1*, IL-6*, IL-6_1*, TNF-α*, TNF-α_1* | Plasma | Flow Cytometry | 6,5 | 12,5 |
| 19 | (Melhorn, Alamoudi et al. 2023) | UK | 5.0(±1.9) months | Post-COVID | Healthy control | 49 36/13 | 18 16/2 | 67 52/15 | 60 (9) | 47 (16) | CRP*, GM-CSF*, ICAM-1*, IFN-γ*, IL-10*, IL-12/23p40*, IL-17*, IL-1β*, IL-4*, IL-6*, IL-7#, IP-10*, MCP-1*, MCP-4*, MIP-1α#, MIP-1β#, PIGF*, TNF-α*, VCAM-1*, VEGF_1#, VEGF_2#, VEGF_3* | Plasma | Multiplex assay | 3,5 | 12,5 |
| 20 | (Alfadda, Rafiullah et al. 2022) | Saudi Arabia | 6 months | Post-COVID | No post-COVID | Unknown | Unknown | Unknown | 51,34 (18.2) | 46.44 (16.8) | CRP#, EGF*, IFN-γ*, IL-10#, IL-1α*, IL-1β#, IL-2#, IL-4#, IL-6*, IL-8#, MCP-1#, TNF-α*, VEGF# | Serum | High-Sensitivity Array | 3,5 | 13,5 |
| 21 | (Patra and Ray 2022) | USA | Unknown | Post-COVID | Healthy control | Unknown | Unknown | Unknown | Unknown | Unknown | TNF-α*, IL-6*, IL-8*, IL-1β#, IL-1α*, IL-10*, IL-4*, IL-13*, TGF-β* | Serum | Elisa | 3,5 | 15 |
| 22 | (Arslan, Aksakal et al. 2022) | Turkey | 3-12 weeks | Post-COVID lung sequelae | Healthy control | 32 25/7 | 26 8/18 | 58 33/25 | 63.656 (8.450) | 39.385 (10.346) | VEGF*, IL-17# | serum | Elisa | 4,5 | 11,5 |
| 23 | (Townsend, Dyer et al. 2020) | Ireland | At least 6 weeks | Post-COVID fatigue | No fatigue | 67 22/45 | 61 37/24 | 128 59/69 | 49.3 (14.3) | 49.7 (16) | CRP*, IL-6* | serum | Elisa | 5,5 | 10,5 |
| 24 | (Allan-Blitz, Goodrich et al. 2023) | USA | At least 28 days | Neurological post-COVID | Healthy control | 20 10/10 | 20 10/10 | 40 20/20 | 42,9 (12.6) | 44.4 (13.6) | CXCL10#, CXCL11#, CXCL12*, CXCL9#, EGF#, Eotaxin*, G-CSF*, HGF*, IFN-γ*, IL-10*, IL-15*, IL-17#, IL-18#, IL-1β*, IL-27#, IL-6*, IL-7*, IL-8#, MCP-1#, MCP-2#, MCP-3#, MCP-4*, M-CSF#, MIP-1α#, MIP-1β#, MIP-3β#, TNFSF10, TNFSF12*, VEGF# | Plasma | Olink Proteomics | 4,5 | 12 |
| 25 | (Meltendorf, Vogel et al. 2022) | Germany | 100(±38) days | Post-COVID | Healthy control | 70 29/41 | 49 16/33 | 119 45/74 | 48 (12) | 46 (13) | IFN-γ*, IL-10*, IL-13*, IL-17*, IL-17_1*, IL-22*, IL-33*, IL-4*, IL-5*, IL-6*, TNF-α* | Serum | Flow Cytometry | 7,5 | 11,5 |
| 26 | (Gomes, Brito et al. 2023) | Brazil | More than 1 month | Long COVID | Healthy control | 15 15/0 | 21 7/14 | 36 22/14 | 47,5 (19.5) | 41 (13) | IFN-γ*, IL-10*, IL-12*, IL-17*, IL-1β*, IL-2*, IL-4*, IL-6*, IL-8*, IP-10*, TNF-α* | Plasma | Flow Cytometry and Elisa | 4,5 | 11 |
| 27 | (Park, Dean et al. 2023) | USA | 6 months | Post-COVID | No post-COVID | 11 8/3 | 10 6/4 | 21 14/7 | 53 (44, 58) | 55 (44, 58.2) | FGF#, IL-1Ra*, PDGF#, TGF-α# | Plasma | Multiplex assay | 4,5 | 11 |
| 28 | (Taeschler, Adamo et al. 2022) | Switzerland | 6 months | Post-COVID-mild | Healthy controls | 79 40/39 | 42 18/24 | 121 58/63 | 36 (29-53) | 32 (28-52) | TNF-α*, IL-6*, CRP* | serum | Flow Cytometry | 7,5 | 11,5 |
|  |  |  |  | Post-COVID-severe |  | 37 23/14 | 42 18/24 | 79 41/38 | 64 (57-73) |  | TNF-α_1*, IL-6_1*, CRP_1* |  |  |  |  |
|  |  |  | 12-month follow-up | Post-COVID-mild |  | 64 30/34 | 42 18/24 | 106 48/58 | 35 (28-45) |  | TNF-α_2*, IL-6_2*, CRP_2* |  |  |  |  |
|  |  |  |  | Post-COVID-severe |  | 26 18/8 | 42 18/24 | 68 36/32 | 65 (53-72) |  | TNF-α_3*, IL-6_3*,CRP_3*, |  |  |  |  |
| 29 | (Nádasdi, Sinkovits et al. 2022) | Hungary | Unknown | Post-COVID | Healthy controls | 26 15/11 | 39 0/39 | 65 15/50 | 45 (34.0–54.0) | 37.0  (34.0–40.0) | CRP#, CRP_1# | Serum | Routine tests | 4,5 | 11,5 |
| 30 | (Garcia-Gasalla, Berman-Riu et al. 2023) | Spain | 8–12 weeks | Post-COVID | fully recovered controls | 49 20/29 | 33 22/11 | 82 42/40 | 56.6 (12.5) | 60.0 (15.5) | CRP, IL-6, IL-10, IFN-γ, IL-1β, IL-1Ra, IL-8, IL-17, IL-18, IL-22, TNF-α | serum | Immunoassay and Nephelometry | 4,5 | 11,5 |
| 31 | (Torres-Ruiz, Lomelín-Gascón et al. 2023) | Mexico | 6 months | Post-COVID | No post-COVID | 33 17/16 | 83 39/44 | 116 56/60 | 56,9 (14.9) | 40.3 (17.4) | EGF*, Eotaxin*, G-CSF*, GM-CSF*, IFN-γ*, IL-10*, IL-12/p70*, IL-12p40*, IL-13*, IL-15*, IL-17*, IL-1α*, IL-1β#, IL-1Ra*, IL-2*, IL-3*, IL-4*, IL-6*, IL-7*, IL-8*, IP-10*, MCP-1*, MIP-1β*, TNF-α*, TNF-β*, VEGF* | Serum | Multiplex assay | 4,5 | 11,5 |
| 32 | (Berezhnoy, Bissinger et al. 2023) | Germany | More than 1 month | Long COVID | Healthy control | 33 17/16 | 83 39/44 | 116 56/60 | 56,9 (14.9) | 40.3 (17.4) | IFN-α, IFN-γ, IL-10, IL-12/p70, IL-17, IL-18, IL-1β, IL-23, IL-33, IL-6, IL-8, MCP-1, TNF-α | Plasma | Flow Cytometry | 7,5 | 11 |
| 33 | (Meisinger, Goßlau et al. 2022) | Germany | 279 days | Post-COVID fatigue | No fatigue | 72 0/72 | 80 0/80 | 152 0/152 | 47 (34;55.5) | 46 (32; 57.5) | CRP*, IFN-γ#, IL-2#, IL-6* | Serum, PBMC | ECLIA (ElektroChemiLuminescence ImmunoAssay ELISpot | 8,5 | 11 |
|  |  |  |  |  |  | 33 33/0 | 96 96/0 | 129 129/0 | 37,57 (51) | 52.5 (36.5; 60) | CRP_1*,IFN-γ_1*,IL-2_1#, IL-6_1* |  |  |  |  |
| 34 | (Schultheiß, Willscher et al. 2023) | Germany | 8 months | Post-COVID | Healthy controls | 258 97/161 | 36 14/22 | 294 111/183 | 51,2 (15–83) | 50 (17–81) | IFN-α*, IL-13*, IL-17*, IL-1β*, IL-4*, IL-6*, IL-8*, TNF-α* | Plasma | Flow Cytometry | 4,5 | 11,5 |
| 35 | (Zhao, Schank et al. 2022) | USA | 3 months | Post-COVID | Healthy controls | 22 5/17 | 22 16/6 | 44 21/23 | 50,04 (13,1) | 43,39 (11,6) | CCL23*, CXCL10*, CXCL11*, CXCL9*, IL-12*, IL-17*, IL-17_1*, IL-1α#, IL-2#, IL-33#, IL-6*, IL-7#, MCP-1*, MCP-3*, M-CSF*, MIP-1α*, MIP-1β*, MIP-3α*, MIP-3β*, SCF*, TGF-α*, TNF-α*, TNF-β*, TNFRSF9* | Plasma | Flow Cytometry | 4,5 | 11,5 |
| 36 | (Wiech, Chroscicki et al. 2022) | Italy | 3- 6 months | Post-COVID-moderate | Healthy control | 17 17/0 | 13 13/0 | 30 30/0 | 43,5 (27-63) | 47 (29-59) | TGF-β#, IFN-γ#, TNF-α#, MCP-1#, IP-10*, IL-1β#, IL-2#, IL-4#, IL-6#, IL-8*, IL-10#, IL-17# | Plasma | Flow Cytometry | 4,5 | 11,5 |
|  |  |  |  | Post-COVID-mild |  | 13 13/0 | 13 13/0 | 26 26/0 | 52 (36-64) |  | TGF-β_1#, IFN-γ_1#, TNF-α_1#, MCP-1_1#, IP-10_1*, IL-1β_1#, IL-2_1#, IL-4_1#, IL-6_1#, IL-8_1#, IL-10_1#, IL-17_1# |  |  |  |  |
| 37 | (Sun, Tang et al. 2021) | USA | 1-3 months | Neurological post-COVID | Healthy control | 8 2/6 | 12 8/4 | 20 10/10 | 45,6 (12.3) | 52.3 (12.4) | IFN-γ*, IL-10*, IL-1β$, IL-4$, IL-6$, IL-8#, TNF-α* | Plasma | Multiplex assay | 4,5 | 12 |
| 38 | (Ong, Fong et al. 2021) | Singapore | 3-6 months | Post-COVID | Healthy controls | Unknown | 24 10/14 | 24 10/14 | 44 (33–56) | 55 (33 – 64.5) | BDNF*, BDNF_1*, BDNF_2*, HGF#, HGF_1*, HGF_2*, IL-12/p70*, IL-12/p70_1*, IL-12/p70_2*, IL-17*, IL-17_1*, IL-17_2*, IL-1β*, IL-1β_1*, IL-1β_2*, MIP-1β*, MIP-1β_1*, MIP-1β_2*, SCF*, SCF_1*, SCF_2*, VEGF*, VEGF_1*, VEGF_2*, VEGF_3*, VEGF_4*, VEGF_5* | Plasma | Multiplex assay | 7,5 | 12,5 |
| 39 | (Loretelli, Abdelsalam et al. 2021) | Italy | 3 months | Post-COVID | Healthy controls | 39 29/10 | 43 18/25 | 82 47/35 | 53,3 (2.4) | 47.3 (2.1) | Eotaxin#, G-CSF*, IFN-γ*, IL-10*, IL-13*, IL-17*, IL-1β*, IL-1Ra*, IL-4*, IL-6*, IL-7*, IL-8*, IL-9#, IP-10#, MCP-1*, MIP-1α*, MIP-1β#, PDGF#, RANTES#, TNF-α# | Serum | Multiplex assay | 4,5 | 12,5 |
| 40 | (Acosta-Ampudia, Monsalve et al. 2022) | Colombia | 7–11 months | Post-COVID | Healthy controls | 12 6/6 | 6 3/3 | 18 9/9 | 50,5 (49.75 to 55.5) | 35 () | G-CSF*, GM-CSF*, IFN-α*, IFN-γ$, IL-10*, IL-12/p70*, IL-13$, IL-17*, IL-1β*, IL-2*, IL-4*, IL-5*, IL-6*, IL-7*, IL-8*, IL-9$, IP-10$, MCP-1*, RANTES$, TNF-α* | Serum | Flow Cytometry | 4,5 | 11,5 |
| 41 | (Fan, Wong et al. 2022) | Singapore | 12.7(±3.6) months | Post-COVID | Healthy controls | 39 28/11 | 124 61/63 | 163 89/74 | 43 (32, 56) | 43 (21–65) | ICAM-1#, IL-6*, IP-10# | Plasma | ELISA | 5,5 | 10,5 |
| 42 | (Sommen, Havdal et al. 2023) | Norway | 179-341 days | Post-COVID | No post-COVID | Unknown | Unknown | Unknown | Unknown | Unknown | CRP*, CRP_1*, Eotaxin#, Eotaxin_1*, IL-9*, IL-9_1*, IP-10*, IP-10_1*, MCP-1*, MCP-1_1*, MIP-1β*, MIP-1β_1*, RANTES*, RANTES_1*, TNF-α#, TNF-α_1* | Plasma | Multiplex assay | 7,5 | 14,5 |
| 43 | (Wu, Deng et al. 2021) | China | 6 months | Post-COVID cardic injury | No cardic injury | 13 4/9 | 14 4/10 | 27 8/19 | 63 (59, 70) | 63 (57, 70) | CRP*, IL-6$ | serum | Unknown | 7,5 | 12,5 |
| 44 | (Wallis, Heiden et al. 2021) | UK | 3 months | Post-COVID chest abnormality | Normal Convalescence | 32 18/14 | 69 36/33 | 101 54/47 | 57.0 (50.3–63.8) | 52.0 (42.5–62.5) | CRP* | serum | Unknown | 7,5 | 11,5 |
| 45 | (Stavileci, Özdemir et al. 2022) | Turkey | 6 months | Post-COVID cardic abnormality | No cardic abnormality | 102 44/58 | 146 50/96 | 248 94/154 | 36,47 (8.72) | 33.64  (9.22) | CRP* | Serum | Unknown | 5,5 | 12,5 |
| 46 | (Sibila, Perea et al. 2022) | Spain | 6 months | Post-COVID lung abnormality | No lung abnormality | 90 59/31 | 125 71/54 | 215 130/85 | 56,9 (12.7) | 65.3? (10.4) | CRP* | Plasma | Unknown | 8,5 | 11,5 |
| 47 | (Kruger, Vlok et al. 2022) | South Africa | 221(± 99) days | Long COVID | Healthy control | 66 21/45 | 29 9/20 | 95 30/65 | 51 (40–60) | 52 (41–57) | IL-6* | Serum | Unknown | 4,5 | 11 |
|  |  |  |  |  |  | 24 5/19 | 29 9/20 | 53 14/39 | 45 (31–58) |  | IL-6_1* |  |  |  |  |
| 48 | (Fernández-de-Las-Peñas, Ryan-Murua et al. 2022) | Spain | 6-months | Post-COVID fatigue | No post-COVID fatigue | 300 140/160 | 112 73/39 | 412 213/199 | 63.0 (15.0) | 59.5 (17.0) | CRP_1#, CRP_2#, CRP_3#, CRP_4# | serum | Unknown | 5,5 | 10,5 |
| 49 | (Durstenfeld, Peluso et al. 2022) | USA | 3 months | CNS-post COVID symptoms | No symptoms | 47 22/25 | 55 38/17 | 102 60/42 | 51,9 (11.9) | 50.3 (12.4) | CRP, IFN-γ, IL-10, IL-6, TNF-α | Plasma | Multiplex assay | 4,5 | 11 |
| 50 | (Díaz-Salazar, Navas et al. 2022) | Spain | 3 months | Post-COVID | No Post-COVID | 36 11/25 | 85 42/43 | 121 53/68 | 46,7 (14) | 45.1 (17) | CRP# | Serum | Immunonephelometric | 7,5 | 12 |
| 51 | (Corrêa, Deus et al. 2022) | Brazil | 11 months | Long COVID | No Long COVID | Unknown | Unknown | Unknown | 67.11 (2.99) | 65.31 (4.77) | TNF-α*, IL-10*, IL-6* | plasma | Elisa | 7,5 | 13,5 |
| 52 | (Aparisi, Ybarra-Falcón et al. 2021) | Spain | 3 months | Long COVID with dyspnea | without dyspnea | 41 11/30 | 29 14/15 | 70 25/45 | 54,9 (10.5) | 54.6 (13.9) | CRP*, IL-6* | Serum | Immunoassay | 4,5 | 11,5 |
| 53 | (Saxena, Saxena et al. 2022) | India | Unknown | Post-COVID | Healthy control | 25 11/14 | 25 13/12 | 50 24/26 | 42 (6.0) | 40 (6.2) | CRP*, TNF-α* | Serum | Elisa | 4,5 | 9,5 |
| 54 | (Colarusso, Maglio et al. 2021) | Italy | 1-3 months | Post-COVID lung abnormality | Healthy control | 52 32/20 | 17 17/0 | 69 49/20 | 50 (10) | 50 (10) | CRP*, CRP_1*, CRP_2*, CRP_3*, IFN-β*, IFN-β_1*, IFN-β_2*, IFN-β_3*, IL-1α*, IL-1α_1*, IL-1α_2*, IL-1α_3*, IL-6#, IL-6_1#, IL-6_2#, IL-6_3#, IP-10*, IP-10_1*, IP-10_2*, IP-10_3*, TGF-β#, TGF-β_1*, TGF-β_2*, TGF-β_3* | Plasma | Elisa | 4,5 | 11,5 |
| 55 | (Phetsouphanh, Darley et al. 2022) | Australia | 4 months | Long COVID | Healthy control | 31 15/16 | 31 15/16 | 62 30/32 | 49,6 (14.9) | 48.9 (12.8) | IL-10#, IL-12/p70#, IL-13*, IL-18*, IL-33#, IL-5#, IL-6*, IL-9#, IP-10*, MCP-1#, PECAM-1#, TGF-β*, TNF-α*, VCAM-1* | Serum | Flow Cytometry | 7,5 | 11,5 |
| 56 | (Petrella, Nenna et al. 2022) | Italy | 1 months | Long COVID | Healthy control | 5 5/0 | 5 5/0 | 10 10/0 | Unknown | Unknown | TGF-β*, IFN-α*, IL-10*, IL-12*, TNF-α*, MCP-1*, IL-2*, IL-6* | serum | Elisa | 3,5 | 12,5 |
|  |  |  |  |  |  | 5 0/5 | 5 0/5 | 10 0/10 |  |  | TGF-β_1*, IFN-α_1*, IL-10_1*, IL-12_1*, TNF-α_1*, MCP-1_1*, IL-2_1*, IL-6_1* |  |  |  |  |
| 57 | (Stepanova, Korol et al. 2023) | Ukrania | 3.3-4.6 months | Post-COVID | No Post-COVID | 31 11/20 | 32 24/8 | 63 35/28 | 48,6 (10.9) | 53.8 (13.5) | CRP* | Serum | Unknown | 4,5 | 10,5 |
| 58 | (Siekacz, Kumor-Kisielewska et al. 2023) | Poland | 3 months | Post-COVID pulmonary complications | No Complications | 40 31/9 | 40 21/19 | 80 52/28 | 56 (12.21) | 53 (11.3) | IFN-α#, IL-6*, sRAGE*, TNF-α* | Serum | Elisa | 7,5 | 10,5 |
| 59 | (Mitrović-Ajtić, Đikić et al. 2023) | Serbia | 2.5 months | Post-COVID | Healthy control | 14 0/14 | 7 0/7 | 21 0/21 | 59,9 (11.3) | 39.4 (9.4) | ICAM-1$, IFN-γ$, IL-10*, IL-1β$, IL-6$, IL-8$, MCP-1$, TGF-β$, TNF-α$, VCAM-1$, | Plasma | Elisa | 4,5 | 11,5 |
|  |  |  |  |  |  | 25 25/0 | 3 3/0 | 28 28/0 | 57,6 (13.2) | 46.3 (5) | ICAM-1_1$, IFN-γ_1*, IL-10_1*,IL-1β_1$,IL-6_1$,IL-8_1$,MCP-1_1$,TGF-β_1$,TNF-α_1$,VCAM-1_1$, |  |  |  |  |
|  |  |  | 5 months |  |  | 9 0/9 | 7 0/7 | 16 0/16 | 64,6 (11.7) | 39.4 (9.4) | ICAM-1_2$, IFN-γ_2#,IL-10_2$,IL-1β_2$, IL-6_2$,IL-8_2$,MCP-1_2$,TGF-β_2$,TNF-α_2$,VCAM-1_2$, |  |  |  |  |
|  |  |  |  |  |  | 13 13/0 | 3 3/0 | 16 16/0 | 52,7 (14.4) | 46.3 (5) | ICAM-1_3$, IFN-γ_3#,IL-10_3$,IL-1β_3$, IL-6_3$,IL-8_3$,MCP-1_3$, TGF-β_3$, TNF-α_3$,VCAM-1_3$ |  |  |  |  |
| 60 | (Johannes, Andrea et al. 2022) | Austria | 3-10 months | Long COVID | Healthy control | 13 4/9 | 13 6/7 | 26 10/16 | 33 (21-53) | 30 (25-43) | CRP*, IL-18#, MCP-1#, TNFR2 # | Serum | Multiplex assay | 7,5 | 11,5 |
| 61 | (Simone, Caitlin et al. 2022) | South Africa | Unknown | Long COVID | No Long COVID | 25 9/16 | 15 1/14 | 40 10/30 | 52 (14) | 42 (8) | PECAM-1* | serum | Elisa | 7,5 | 11,5 |
| 62 | (García-Abellán, Fernández et al. 2022) | Spain | 6-12 months | Long COVID | No Long COVID | 14 5/9 | 58 39/19 | 72 44/28 | 59.5 (53-71) | 60 (52-71) | CRP*, IL-6* | Serum | Immunoassay | 4,5 | 12,5 |
| 63 | (Gameil, Marzouk et al. 2021) | Egypt | 3-6 months | Long COVID | Healthy control | 120 67/53 | 120 69/51 | 240 136/104 | 38,29 (5.27) | 37.25 (4.87) | CRP* | Serum | Unknown | 5,5 | 9,5 |
| 64 | (Taha, Samaan et al. 2021) | Egypt | 6 months | Post-COVID-19 arthritis-positive | Post-COVID-19 arthritis-negative | 37 23/14 | 63 38/25 | 100 61/39 | 63.06 (12.33) | 50.93 (19.95) | IL-6$ | serum | Unknown | 4,5 | 12 |
| 65 | (Queiroz, Neves et al. 2022) | Brazil | At least 1 months | Post-COVID | No Post-COVID | Unknown | Unknown | Unknown | Unknown | Unknown | IFN-γ#, IL-10*, IL-17*, IL-2*, IL-4#, IL-6#, TNF-α# | Plasma | Flow Cytometry | 4,5 | 14 |
| 66 | (Montefusco, Ben Nasr et al. 2021) | Italy | 2 months | Post-COVID | Healthy control | 10 7/3 | 15 10/5 | 25 17/8 | 46,9 (3.8) | 47.2 (3.1) | G-CSF*, IFN-γ*, IL-10*, IL-13*, IL-17*, IL-1β*, IL-2*, IL-4*, IL-5*, IL-6*, IL-7*, IL-8*, MCP-1* | Serum | Multiplex assay | 5 | 11 |
| 67 | (Dugani, Mehta et al. 2022) | India | Unknown | Post-COVID | Healthy control | 69 37/32 | 66 45/21 | 135 82/53 | 51,71 (15.9) | 46.33 (18.17) | CRP*, IL-6$ | Serum | Unknown | 5,5 | 12 |
| 68 | (Cervia, Zurbuchen et al. 2022) | Switzerland | 6 months | Long COVID | Healthy Control | 89 43/46 | 40 17/23 | 129 60/69 | 34 (27–49) | 33 (29–53) | CRP*, IL-6*, TNF-α*, | Serum | Luminex Assay | 5,5 | 9,5 |
|  |  |  |  |  |  | 45 32/13 | 40 17/23 | 85 49/36 | 64 (54–74) |  | CRP_1*, IL-6_1*, TNF-α_1* | Serum |  |  |  |
| 69 | (Magdy, Eid et al. 2022) | Egypt | Unknown | Post-COVID | No Post-COVID | 45 15/30 | 45 16/29 | 90 31/59 | 43 (15.85) | 45.71 (13.89) | CRP* | Serum | Unknown | 4,5 | 12,5 |
| 70 | (Peluso, Sans et al. 2022) | USA | 3 months | CNS-post COVID symptoms | No symptoms | 52 15/37 | 69 40/29 | 121 55/66 | 48 (38.5 – 57) | 43 (36 – 55) | IL-6*, GFAP*, NfL*, MCP-1*, IFN-γ*, IL-10*, TNF-α*, IP-10*, IL-6_1*, GFAP_1*, NfL_1*, MCP-1_1#, IFN-γ_1*, IL-10_1#, TNF-α_1*, IP-10_1* | plasma | Multiplex assay | 4,5 | 13 |
| 71 | (Patterson, Guevara-Coto et al. 2021) | USA | More than 3 months | Post-COVID | Healthy control | Unknown | Unknown | Unknown | Unknown | Unknown | TNF-α*, IL-4*, IL-13*, IL-2*, GM-CSF#, RANTES*, MIP-1α*, IL-6*, IL-10*, IFN-γ* | plasma | Flow Cytometry | 4,5 | 15 |
| 72 | (Flaskamp, Roubal et al. 2022) | Germany | 4.3–11.6 months | Post-COVID | Healthy control | 17 1/16 | 14 2/12 | 31 3/28 | 42 (27–66) | 45 (31–58) | IL-10*, IL-18#, IL-6*, MCP-1#, PIGF*, sRAGE#, sVEGFR#, TGF-β*, TNF-α*, TNFSF14*, VEGF* | Serum | Flow Cytometry and Elisa | 7,5 | 11,5 |
|  |  |  | 8.2–11.1 months |  |  | 13 2/11 |  | 27 4/23 | 43 (24–59) |  | IL-10_1*,IL-18_1#,IL-6_1#,MCP-1_1#,PIGF_1*, sRAGE_1#, sVEGFR_1#, TGF-β_1$, TNF-α_1*, TNFSF14_1*, VEGF_1# |  |  |  |  |
| 73 | (Peluso, Lu et al. 2021) | USA | More than 6 months | Post-COVID | Healthy control | 73 28/45 | 48 27/21 | 121 55/66 | 44 (36–56) | 44.5 (37–58.5) | IL-6*, MCP-1*, IFN-γ*, IL-10*, TNF-α*, IP-10*, IL-6_1*, MCP-1_1#, IFN-γ_1*, IL-10_1*, TNF-α_1*, IP-10_1* | Plasma | Immunoassay | 4,5 | 11,5 |
| 74 | (Wechsler, Butuci et al. 2022) | USA | At least 1 months | Post-COVID | Healthy control | 13 1/12 | 20 14/6 | 33 15/18 | 49 (24–73) | 36 (18–51) | IL-6*, CXCL1* | serum | Flow Cytometry | 4,5 | 12 |
| 75 | (Aparisi, Ybarra-Falcón et al. 2022) | Spain | 3 months | Post-COVID headache | No headache | 10 0/10 | 60 25/35 | 70 25/45 | 46,9 (8.45) | 56.13 (11.9) | CRP*, IL-6* | Serum | Immunoassay | 4,5 | 10,5 |
| 76 | (Martone, Tosato et al. 2022) | Italy | 3 months | Post-COVID sarcopenia | No sarcopenia | 106 42/64 | 435 225/210 | 541 267/274 | 59,9 (15.9) | 51.5 (14.6) | CRP* | Serum | Unknown | 5,5 | 11 |
| 77 | (Littlefield, Watson et al. 2022) | USA | 6 months | Post-COVID | No Post-COVID | 20 10/10 | 20 10/10 | 40 20/20 | 53 (22–69) | 50 (27–73) | CRP*, IL-6* | Plasma | Elisa | 4,5 | 12 |
| 78 | (Al-Hakeim, Khairi Abed et al. 2023) | Iraq | 3-6 months | Long COVID | Healthy control | 20 12/8 | 30 16/14 | 50 28/22 | 32.5 (8.4) | 29.2 (8.1) | CRP* | Serum | Elisa | 7 | 7,5 |
| 79 | (Al-Hakeim, Al-Rubaye et al. 2023) | Iraq | 3-6 months | Long COVID | Healthy control | 67 42/25 | 36 30/6 | 103 72/31 | 29,3 (7.2) | 30.9 (8.3) | CRP* | Serum | Elisa | 7 | 8,5 |
|  |  |  |  |  |  | 51 43/8 |  | 87 73/14 | 33,1 (8.6) |  | CRP_1* |  |  |  |  |
| 80 | (Al-Hakeim, Al-Rubaye et al. 2023c) | Iraq | 3-6 months | Long COVID | Healthy control | 30 22/8 | 57 42/15 | 87 64/23 | 29.8 (7.3) | 28.3 (7.6) | Caspase 1#, CRP*, IL-10*,IL-18*, IL-1β* | Serum | Elisa | 7 | 7,5 |
|  |  |  |  |  |  | 56 40/16 |  | 113 82/31 | 27.6 (5.4) |  | Caspase 1_1#,CRP_1*, IL-10_1*, IL-18_1#, IL-1β_1* |  | Elisa |  |  |
| 81 | (Al-Hakeim, Al-Rubaye et al. 2022b) | Iraq | 3-6 months | Long COVID | Healthy control | 86 62/24 | 39 24/15 | 125 86/39 | 28.4 (6.2) | 28.1 (7.6) | Caspase 1*, CRP*, IL-18*, IL-1β* | Serum | Elisa | 7 | 7,5 |
| 82 | (Al-Hakeim, Abed et al. 2023a) | Iraq | 3-6 months | Long COVID | Healthy control | Unknown | Unknown | Unknown | Unknown | Unknown | CRP* | Serum | Elisa | 7 | 9,5 |

*: Indicates that Long COVID patients have increased mean level of the measured inflammatory biomarker compared to controls

^#:^ Indicates that Long COVID patients have reduced mean level of the measured inflammatory biomarker compared to controls

$: There is much difference between mean levels of Long COVID patients versus controls.

IL: Interleukin, CRP: C-reactive protein, MCP-1: Monocyte Chemoattractant Protein-1, TNF: Tumor necrosis factor, TGF: Transforming growth factor, IFN: Interferon, PIGF: Placental growth factor, sRAGE: Soluble receptor for advanced glycation end products, sVEGFR: Soluble vascular endothelial growth factor receptor-1, TNFSF: Tumor necrosis factor superfamily, VEGF: Vascular endothelial growth factor, GFAP: Glial fibrillary acidic protein, NfL: Neurofilament light polypeptide, RANTES: Regulated upon Activation, Normal T Cell Expressed and Presumably Secreted, MIP: Macrophage inflammatory protein, GM-CSF: Granulocyte-macrophage colony-stimulating factor, IP-10: Interferon gamma-induced protein 10, G-CSF: Granulocyte colony-stimulating factor, VCAM: Vascular cell adhesion molecule-1, ICAM: Intercellular Adhesion Molecule 1, PECAM: Platelet endothelial cell adhesion molecule, PDGF: Platelet-derived growth factor, IL-1Ra: Interleukin-1 receptor antagonist protein, SCF: Stem cell factor, HGF: Hepatocyte growth factor, FGF: Fibroblast growth factors, BDNF: Brain-derived neurotrophic factor.

**ESF. Table 7**. Results of Meta-regression

| **Variables** | **No. of Studies** | **Covariates** | **1-sided p-value** | **Z-Value** |
| --- | --- | --- | --- | --- |
| IRS/CIRS | 28 | Age | 0.004 | -2.64 |
| IRS | 60 | Latitude | 0.003 | -2.66 |
| CIRS | 32 | More than 6 Months | 0.009 | 2.35 |
|  | 6 | # of patients hospitalized during AP | 0.026 | 1.93 |
| M1/M2 | 6 | # of patients hospitalized during AP | 0.047 | -1.67 |
|  | 30 | Age | 0.001 | -2.91 |
| M1 Macrophage | 59 | Latitude | 0.017 | -2.12 |
| M2 Macrophage | 33 | More than 6 Months | 0.023 | 1.98 |
|  | 6 | # of patients hospitalized during AP | 0.002 | 2.78 |
| Th1/Th2 | 45 | Latitude | 0.000 | -4.11 |
| Th1 | 45 | Latitude | 0.010 | -2.31 |
| Th2 | 50 | More than 6 Months | 0.019 | 2.23 |
| Th17 | 55 | Latitude | 0.025 | -1.95 |
|  | 49 | Less than 3 months | 0.050 | -1.64 |
|  | 9 | # of patients hospitalized during AP | 0.048 | -1.66 |
|  | 5 | # of patients admitted to ICU | 0.044 | -1.70 |
|  | 43 | Age | 0.015 | -2.17 |
| Neurotoxicity | 62 | Latitude | 0.0054 | -2.55 |
|  | 47 | Age | 0.028 | -1.91 |
| CRP | 9 | # of patients hospitalized during AP | 0.044 | -1.70 |
|  | 7 | # of patients admitted to ICU | 0.008 | -2.38 |
|  | 30 | Age | 0.012 | -2.23 |
|  | 38 | Latitude | 0.036 | -1.80 |
| IL-6 | 9 | # of patients hospitalized during AP | 0.029 | -1.88 |
|  | 5 | # of patients admitted to ICU | 0.044 | -1.70 |
|  |  | Fasting | 0.003 | -2.66 |
| IL-1β | 23 | Latitude | 0.022 | -2 |
| TNF-α | 6 | Smoking | 0.004 | -2.58 |
| IL-2 | 17 | Latitude | 0.019 | -2.06 |
| IL-22 | 4 | Sample size | 0.023 | 1.99 |
| IL-7 | 11 | Less than 3 months | 0.012 | 2.24 |
|  | 12 | Serum | 0.003 | 2.66 |
| IP-10 | 4 | BMI | 0.0004 | 3.35 |
| IL-8 | 21 | More than 6 months | 0.039 | 1.76 |
|  |  | Serum | 0.001 | 3.05 |
| IL-13 | 11 | Serum | 0.011 | 2.29 |
| MIP-1α | 9 | Serum | 0.050 | 1.64 |
| IL-1Ra | 8 | Latitude | 0.0004 | 3.33 |
|  | 7 | Age | 0.0017 | -2.93 |
| G-CSF | 6 | More than 6 months | 0.011 | 2.28 |
|  |  | Serum | 0.048 | 1.66 |
|  | 5 | Female | 0.042 | -1.73 |
| VCAM | 6 | Sample size | 0.0004 | -3.34 |
|  |  | Serum | 0.036 | 1.79 |

**M1 Macrophage:** IL-1β, IL-6, TNF-α, IL-12p70, IL-15, CCL2, CCL5, CXCL1, CXCL8, CXCL9, CXCL10

**M2 Macrophage:** IL-10, IL-4, IL-13, VEGF, PDGF, sIL-1RA

**T helper (Th)-1:** IL-2, sIL-2R, IFN-a, IFN-γ, IL-12p70, IL-16, TNF-α, TNF-β

**T helper (Th)-2:** IL-4, IL-5, IL-9, IL-13, IL-10, IL-6

**T helper (Th)-17:** IL-6, IL-17

**IRS:** IL-1α, IL-1β, IL-6, TNF-α, IL-12p70, IL-15, IL-16, IL-17, IL-18, CCL2, CCL3, CCL4, CCL5, CCL7, CCL11, CXCL1, CXCL8, CXCL9, CXCL10, IL-2, IFN-α, IFN-γ, TNF-β, GM-CSF, G-CSF.

**CIRS:** IL-4, IL-10, sIL-1RA, sIL-2R

**Neurotoxicity:** IL-1β, IL-6, TNF-α, IL-2, IFN-γ, IL-12p70, IL-16, IL-17, CCL2, CCL3, CCL5, CCL11, CXCL1, CXCL8, CXCL10, GM-CSF, M-CSF

IRS: Immune-response system, CIRS: Compensatory immune-response system, M: Macrophage, Th: T helper, CRP: C-reactive protein, IL: interleukin, TNF: Tumor necrosis factor, IP-10: Interferon gamma-induced protein 10, VEGF: Vascular endothelial growth factor, MIP-1α: Macrophage inflammatory protein-1 alpha, IL-1Ra: Interleukin-1 receptor antagonist, G-CSF: Granulocyte colony-stimulating factor, VCAM: Vascular cell adhesion molecule.

**Abbreviation:**

IRS: Immune-response system, CIRS: Compensatory immune-response system, M: Macrophage, Th: T helper, IL: Interleukin, CRP: C-reactive protein, MCP-1: Monocyte Chemoattractant Protein-1, TNF: Tumor necrosis factor, TGF: Transforming growth factor, IFN: Interferon, PIGF: Placental growth factor, sRAGE: Soluble receptor for advanced glycation end products, sVEGFR: Soluble vascular endothelial growth factor receptor-1, TNFSF: Tumor necrosis factor superfamily, VEGF: Vascular endothelial growth factor, GFAP: Glial fibrillary acidic protein, NfL: Neurofilament light polypeptide, RANTES: Regulated upon Activation, Normal T Cell Expressed and Presumably Secreted, MIP: Macrophage inflammatory protein, GM-CSF: Granulocyte-macrophage colony-stimulating factor, IP-10: Interferon gamma-induced protein 10, G-CSF: Granulocyte colony-stimulating factor, VCAM: Vascular cell adhesion molecule-1, ICAM: Intercellular Adhesion Molecule 1, PECAM: Platelet endothelial cell adhesion molecule, PDGF: Platelet-derived growth factor, IL-1Ra: Interleukin-1 receptor antagonist protein, SCF: Stem cell factor, HGF: Hepatocyte growth factor, FGF: Fibroblast growth factors, BDNF: Brain-derived neurotrophic factor.
