## Supplementary file for "Immune activation and immune-associated neurotoxicity in Long-COVID: A systematic review and meta-analysis of 82 studies comprising 58 cytokines/chemokines/growth factors"

**Table 1:** The outcomes and number of patients with Long COVID (LC) disease and healthy control along with the side of standardized mean difference (SMD) and the 95% confidence intervals with respect to zero SMD.

| **Outcome profiles** | **n studies** | **Side of 95% confidence intervals** | | | | **Patient**  **Cases** | **Control**  **Cases** | **Total number of participants** |
| --- | --- | --- | --- | --- | --- | --- | --- | --- |
|  |  | **< 0** | **Overlap 0**  **and SMD < 0** | **Overlap 0 and SMD > 0** | **> 0** |  |  |  |
| IL-6 | 52 | 3 | 15 | 11 | 23 | 2382 | 2544 | 5926 |
| IL-1β | 23 | 2 | 6 | 7 | 8 | 715 | 697 | 1412 |
| TNF-α | 41 | 5 | 13 | 12 | 11 | 2079 | 2115 | 4194 |
| IL-2 | 17 | 2 | 5 | 4 | 6 | 656 | 685 | 1341 |
| IL-22 | 4 | 0 | 1 | 1 | 2 | 188 | 145 | 333 |
| IL-7 | 12 | 3 | 3 | 4 | 2 | 582 | 943 | 1525 |
| IP-10 | 20 | 1 | 5 | 10 | 4 | 1083 | 1452 | 2535 |
| IL-8 | 21 | 1 | 8 | 4 | 8 | 820 | 1141 | 1961 |
| VEGF | 12 | 0 | 4 | 6 | 2 | 607 | 968 | 1575 |
| IL-13 | 11 | 0 | 1 | 4 | 6 | 444 | 372 | 816 |
| MIP-1α | 9 | 1 | 2 | 2 | 4 | 668 | 811 | 1479 |
| IL-1Ra | 8 | 0 | 0 | 4 | 4 | 502 | 871 | 1373 |
| G-CSF | 6 | 0 | 0 | 3 | 3 | 136 | 210 | 346 |
| VCAM-1 | 6 | 0 | 1 | 4 | 1 | 428 | 709 | 1137 |
| EGF | 4 | 0 | 1 | 2 | 1 | 91 | 188 | 279 |
| MCP-4 | 3 | 0 | 0 | 2 | 1 | 78 | 57 | 135 |
| MIP-3α | 3 | 0 | 0 | 0 | 3 | 105 | 76 | 181 |
| CXCL1 | 3 | 0 | 0 | 0 | 3 | 104 | 77 | 174 |
| SCF | 2 | 0 | 0 | 0 | 2 | 56 | 46 | 102 |

IL: interleukin, TNF: Tumor necrosis factor, IP-10: Interferon gamma-induced protein 10, VEGF: Vascular endothelial growth factor, MIP-1α: Macrophage inflammatory protein-1 alpha, IL-1Ra: Interleukin-1 receptor antagonist, G-CSF: Granulocyte colony-stimulating factor, VCAM: Vascular cell adhesion molecule, EGF: Epidermal growth factor, MCP-4: Monocyte chemoattractant protein, SCF: Stem cell factor

| **Outcome feature sets** | **n** | **Groups** | **SMD** | **95% CI** | **z** | **p** | **Q** | **df** | **p** | **I^2^ (%)** | **τ^2^** | **Τ** |
| --- | --- | --- | --- | --- | --- | --- | --- | --- | --- | --- | --- | --- |
| IL-6 | 52 | Overall | 0.333 | 0.167;0.499 | 3.930 | <0.0001 | 336.010 | 51 | <0.0001 | 84.822 | 0.289 | 0.538 |
| IL-1β | 23 | Overall | 0.380 | 0.099;0.660 | 2.655 | 0.008 | 119.940 | 22 | <0.0001 | 81.657 | 0.356 | 0.597 |
| TNF-α | 41 | Overall | 0.225 | 0.037;0.414 | 2.343 | 0.019 | 282.685 | 40 | <0.0001 | 85.850 | 0.296 | 0.544 |
| IL-2 | 17 | Overall | 0.312 | -0.065;0.689 | 1.642 | 0.104 | 136.926 | 16 | <0.0001 | 88.315 | 0.501 | 0.708 |
| IL-22 | 4 | Overall | 0.421 | 0.130;0.711 | 2.839 | 0.005 | 4 | 3 | 0.261 | 25.001 | 0.022 | 0.149 |
| IL-7 | 12 | Overall | 0.179 | -0.273;0.631 | 0.777 | 0.437 | 116.147 | 11 | <0.0001 | 90.529 | 0.531 | 0.729 |
| IP-10 | 20 | Overall | 0.208 | 0.030;0.386 | 2.292 | 0.022 | 64.441 | 19 | <0.0001 | 70.516 | 0.101 | 0.318 |
| IL-8 | 21 | Overall | 0.290 | 0.019;0.561 | 2.095 | 0.036 | 106.056 | 20 | 0.000 | 81.142 | 0.278 | 0.527 |
| VEGF | 12 | Overall | 0.258 | 0.010;0.507 | 2.042 | 0.041 | 37.114 | 11 | 0.000 | 70.361 | 0.118 | 0.344 |
| IL-13 | 11 | Overall | 0.591 | 0.187;0.995 | 2.870 | 0.004 | 58.848 | 10 | 0.000 | 83.007 | 0.360 | 0.600 |
| MIP-1α | 9 | Overall | 0.372 | -0.099;0.843 | 1.547 | 0.122 | 88.952 | 8 | 0.000 | 91.006 | 0.447 | 0.668 |
| IL-1Ra | 8 | Overall | 0.936 | 0.407;1.464 | 3.468 | 0.001 | 80.708 | 7 | 0.000 | 91.327 | 0.499 | 0.706 |
| G-CSF | 6 | Overall | 0.867 | 0.240;1.494 | 2.711 | 0.007 | 27.816 | 5 | 0.000 | 82.025 | 0.470 | 0.686 |
| VCAM-1 | 6 | Overall | 0.331 | 0.034;0.628 | 2.183 | 0.029 | 11.787 | 5 | 0.038 | 57.582 | 0.070 | 0.265 |
| EGF | 4 | Overall | 0.406 | -0.152;0.964 | 1.427 | 0.154 | 11.226 | 3 | 0.011 | 73.277 | 0.230 | 0.480 |
| MCP-4 | 3 | Overall | 0.476 | 0.034;0.918 | 2.110 | 0.035 | 2.684 | 2 | 0.261 | 25.497 | 0.040 | 0.200 |
| MIP-3α | 3 | Overall | 0.776 | 0.454;1.099 | 4.713 | 0.000 | 0.784 | 2 | 0.676 | 0.000 | 0.000 | 0.000 |
| CXCL1 | 3 | Overall | 1.226 | 0.753;1.698 | 5.083 | <0.0001 | 3.745 | 2 | 0.154 | 46.599 | 0.081 | 0.285 |
| SCF | 2 | Overall | 1.095 | 0.347;1.844 | 2.868 | 0.004 | 3.110 | 1 | 0.078 | 67.843 | 0.198 | 0.445 |

IL: interleukin, TNF: Tumor necrosis factor, IP-10: Interferon gamma-induced protein 10, VEGF: Vascular endothelial growth factor, MIP-1α: Macrophage inflammatory protein-1 alpha, IL-1Ra: Interleukin-1 receptor antagonist, G-CSF: Granulocyte colony-stimulating factor, VCAM: Vascular cell adhesion molecule, EGF: Epidermal growth factor, MCP-4: Monocyte chemoattractant protein, SCF: Stem cell factor

**Table 3:** Results on publication bias.

| **Outcome feature sets** | **Fail safe n** | **Z** **Kendall’s τ** | **p** | **Egger’s t test (df)** | **p** | **Missing studies (side)** | **After Adjusting** |
| --- | --- | --- | --- | --- | --- | --- | --- |
| IL-6 | 9.516 | 0.102 | 0.459 | 0.540(50) | 0.295 | 7 (Right) | 0.465 (0.290;0.641) |
| IL-1β | 5.747 | 0.158 | 0.437 | 0.884 (21) | 0.193 | 2 (Right) | 0.484 (0.196;0.772) |
| TNF-α | 5.746 | 0.213 | 0.415 | 0.664(39) | 0.255 | 8 (Right) | 0.419(0.222;0.616) |
| IL-2 | 3.373 | 0.453 | 0.325 | 0.916(15) | 0.187 | 1 (Right) | 0.444 (0.046;0.843) |
| IL-22 | 3.013 | 1.698 | 0.044 | 6.094(2) | 0.012 | 1 (Right) | 0.482 (0.183;0.782) |
| IL-7 | 1.693 | 0.342 | 0.365 | 0.235(10) | 0.409 | 3 (Right) | 0.585 (0.070;1.100) |
| IP-10 | 3.868 | 1.654 | 0.049 | 1.311(18) | 0.103 | 1 (Right) | 0.234 (0.055;0.414) |
| IL-8 | 4.378 | 0.392 | 0.347 | 0.735 (19) | 0.235 | 2 (Right) | 0.408 (0.125;0.690) |
| VEGF | 3.489 | 1.165 | 0.121 | 0.865 (10) | 0.203 | 2 (Right) | 0.370 (0.104;0.635) |
| IL-13 | 6.610 | 0.934 | 0.175 | 0.909(9) | 0.193 | 0 | - |
| MIP-1α | 4.165 | 0.521 | 0.301 | 1.330(7) | 0.112 | 1 (Right) | 0.516 (0.013;1.018) |
| IL-1Ra | 9.430 | 1.360 | 0.086 | 2.588(6) | 0.020 | 0 | - |
| G-CSF | 5.963 | 1.502 | 0.066 | 1.024(4) | 0.181 | 1 (Right) | 1.027 (0.406;1.649) |
| VCAM-1 | 3.138 | 0.751 | 0.226 | 6.825 (4) | 0.001 | 3 (Left) | 0.149 (-0.089;0.338) |
| EGF | 2.906 | 0.339 | 0.367 | 0.336(2) | 0.384 | 1 (Right) | 0.616 (0.019;1.213) |
| MCP-4 | 2.361 | 0.000 | 0.500 | 0.525(1) | 0.345 | 1 (Right) | 0.607 (0.155;1.058) |
| MIP-3α | 4.776 | 0.000 | 0.500 | 1.942 (1) | 0.151 | 2 (Left) | 0.638 (0.377;0.899) |
| CXCL1 | 7.054 | 0.000 | 0.500 | 0.545(1) | 0.341 | 0 | - |

IL: interleukin, TNF: Tumor necrosis factor, IP-10: Interferon gamma-induced protein 10, VEGF: Vascular endothelial growth factor, MIP-1α: Macrophage inflammatory protein-1 alpha, IL-1Ra: Interleukin-1 receptor antagonist, G-CSF: Granulocyte colony-stimulating factor, VCAM: Vascular cell adhesion molecule, EGF: Epidermal growth factor, MCP-4: Monocyte chemoattractant protein.

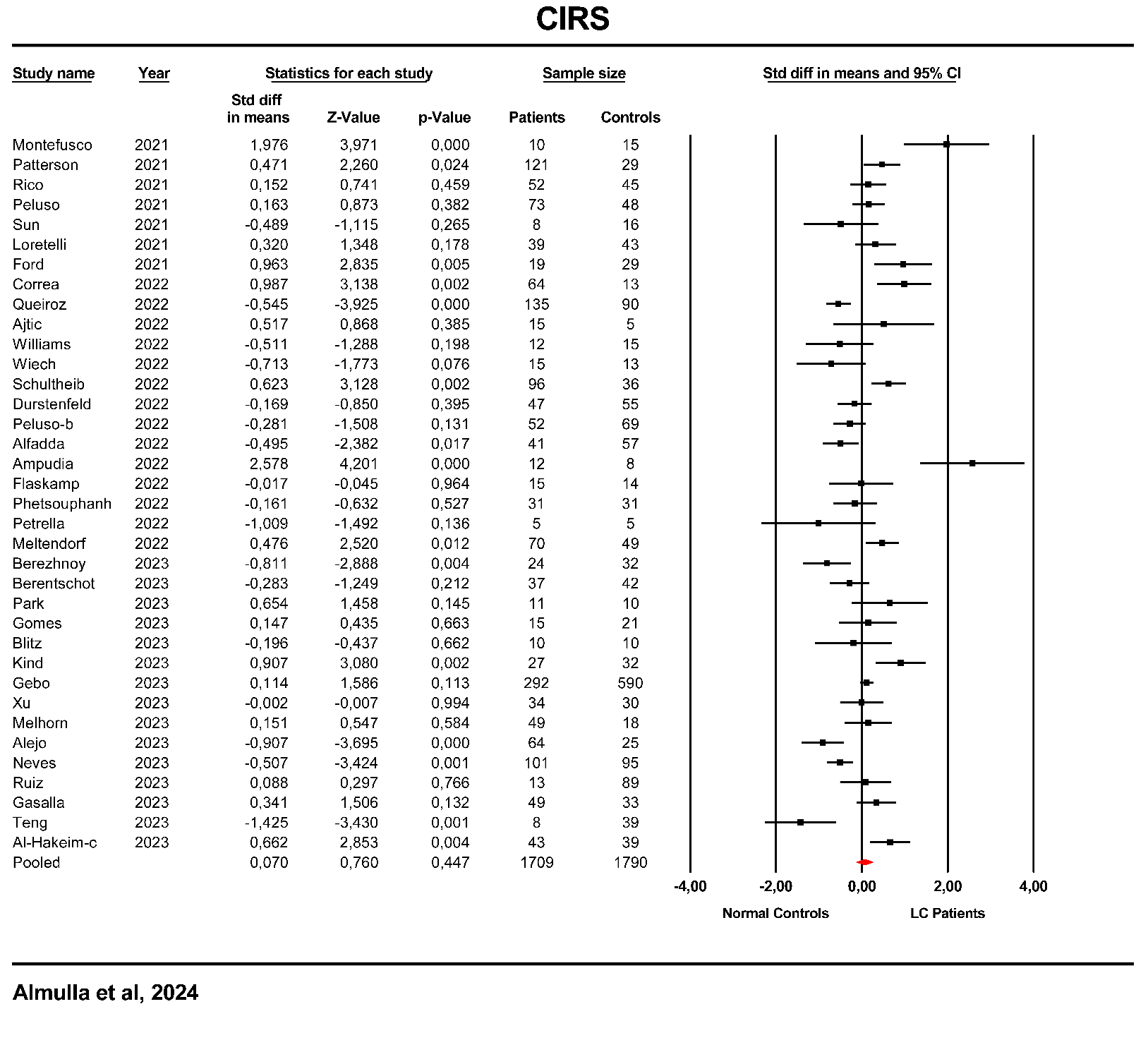

**ESF, Figure 1**. Forest plot of compensatory immune response system (CIRS) in patients with Long COVID (LC) and normal controls.

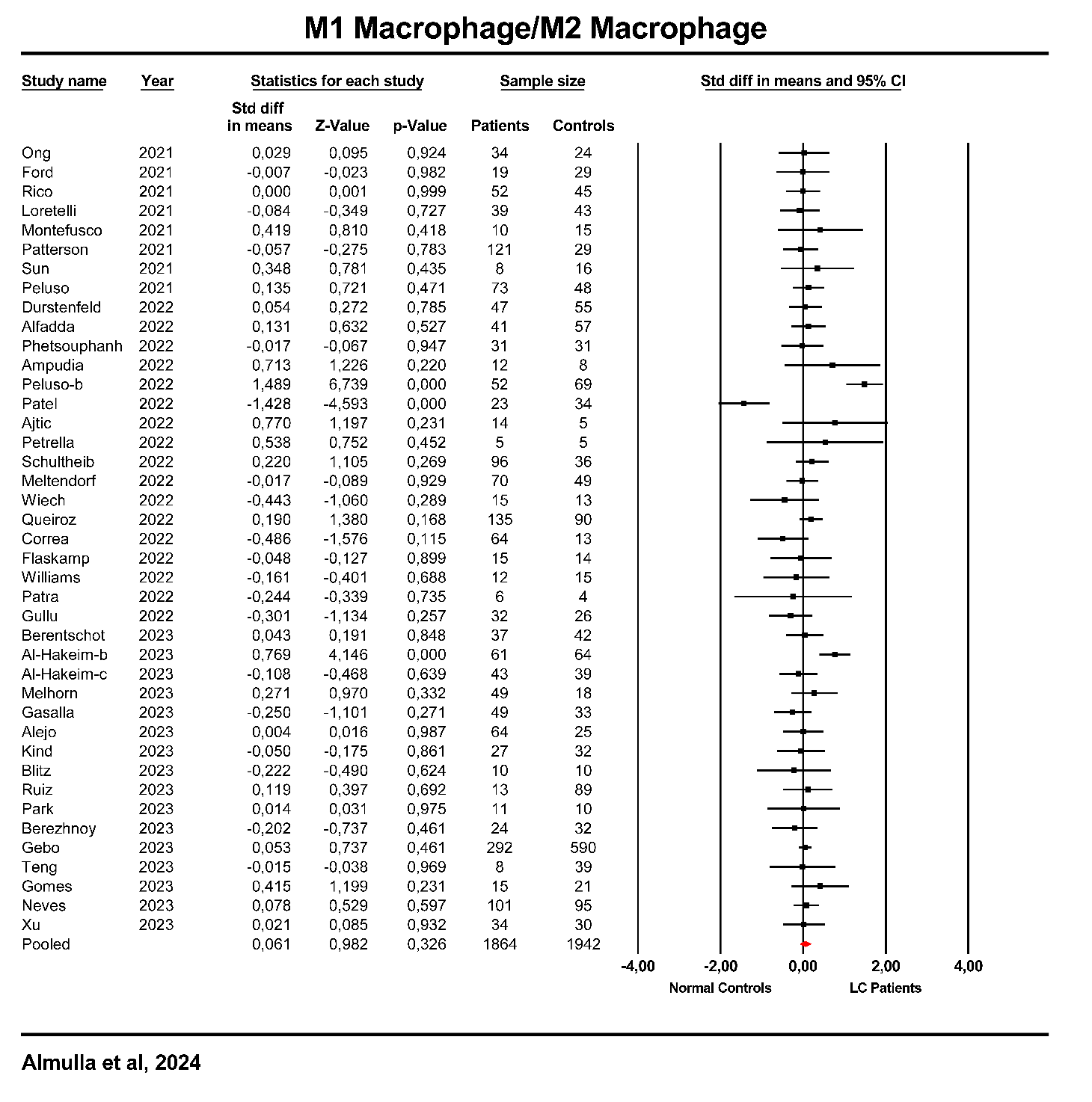

**ESF, Figure 2**. Forest plot of M1 macrophage/ M2 macrophage ratio in patients with Long COVID (LC) and normal controls.

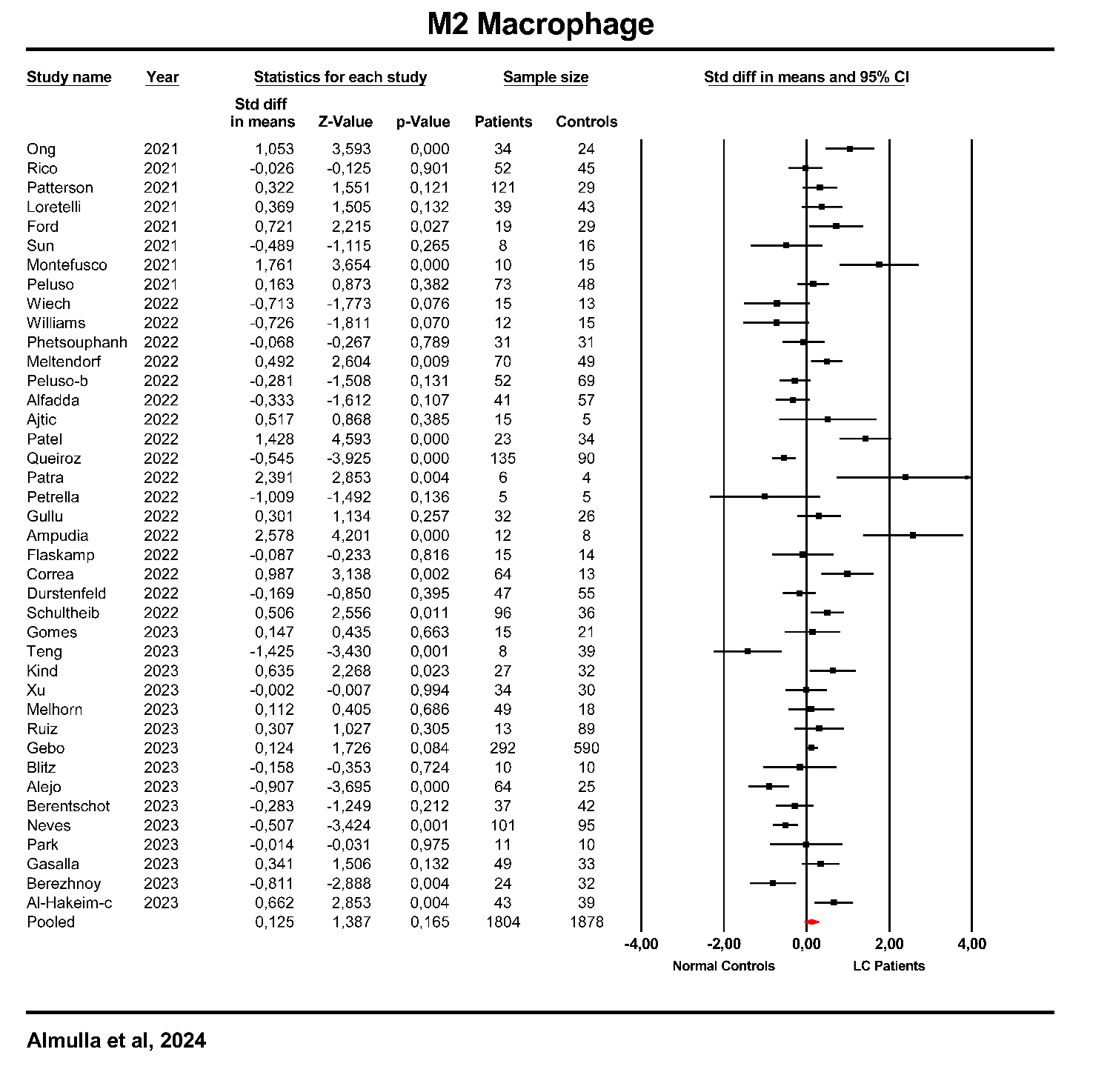

**ESF, Figure 3**. Forest plot of M2 macrophage ratio in patients with Long COVID (LC) and normal controls.

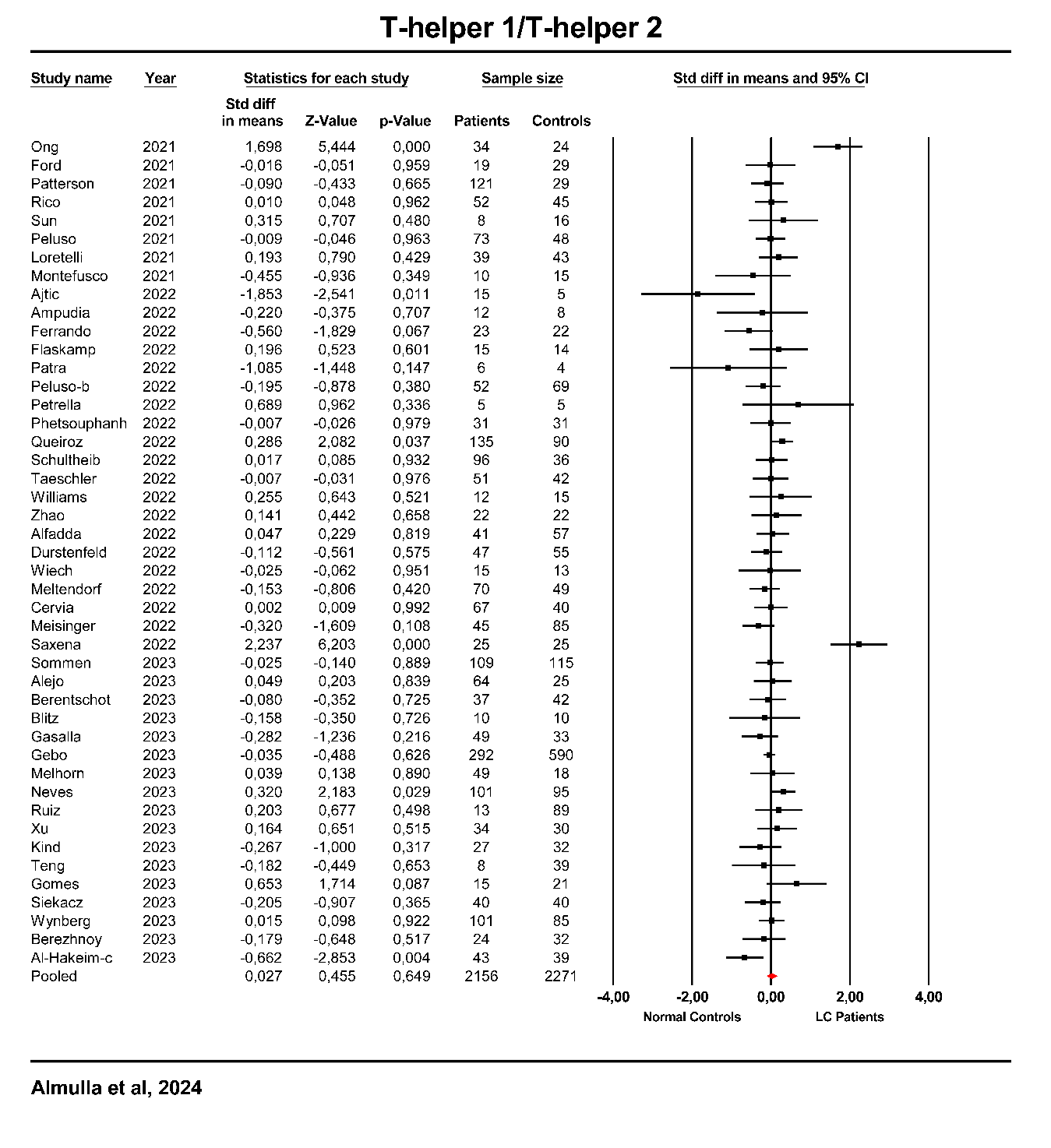

**ESF, Figure 4**. Forest plot of T-helper 1/T-helper 2 ratio in patients with Long COVID (LC) and normal controls.

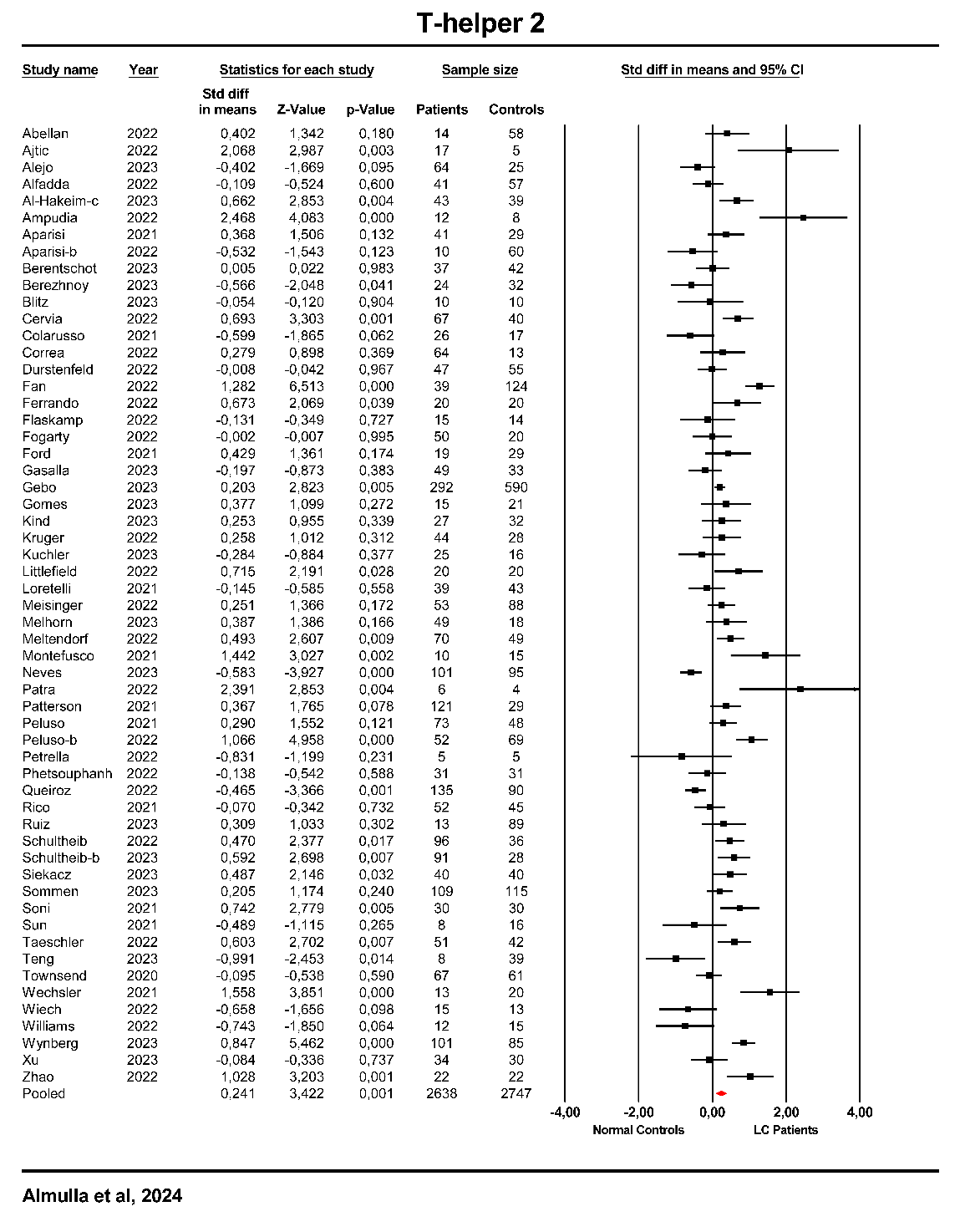

**ESF, Figure 5**. Forest plot of T-helper 2 ratio in patients with Long COVID (LC) and normal controls.

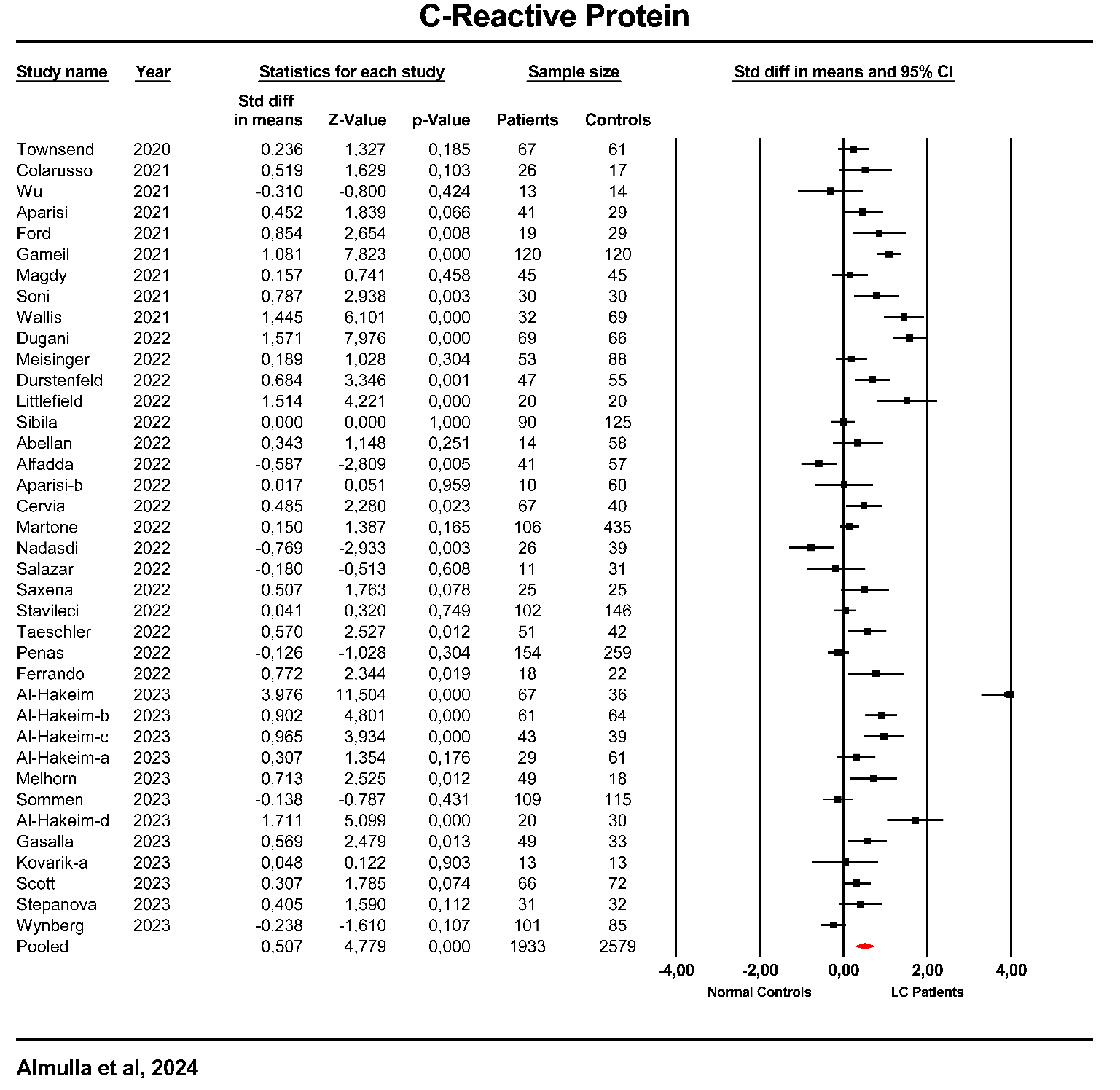

**ESF, Figure 6**. Forest plot of C-reactive protein (CRP) ratio in patients with Long COVID (LC) and normal controls.

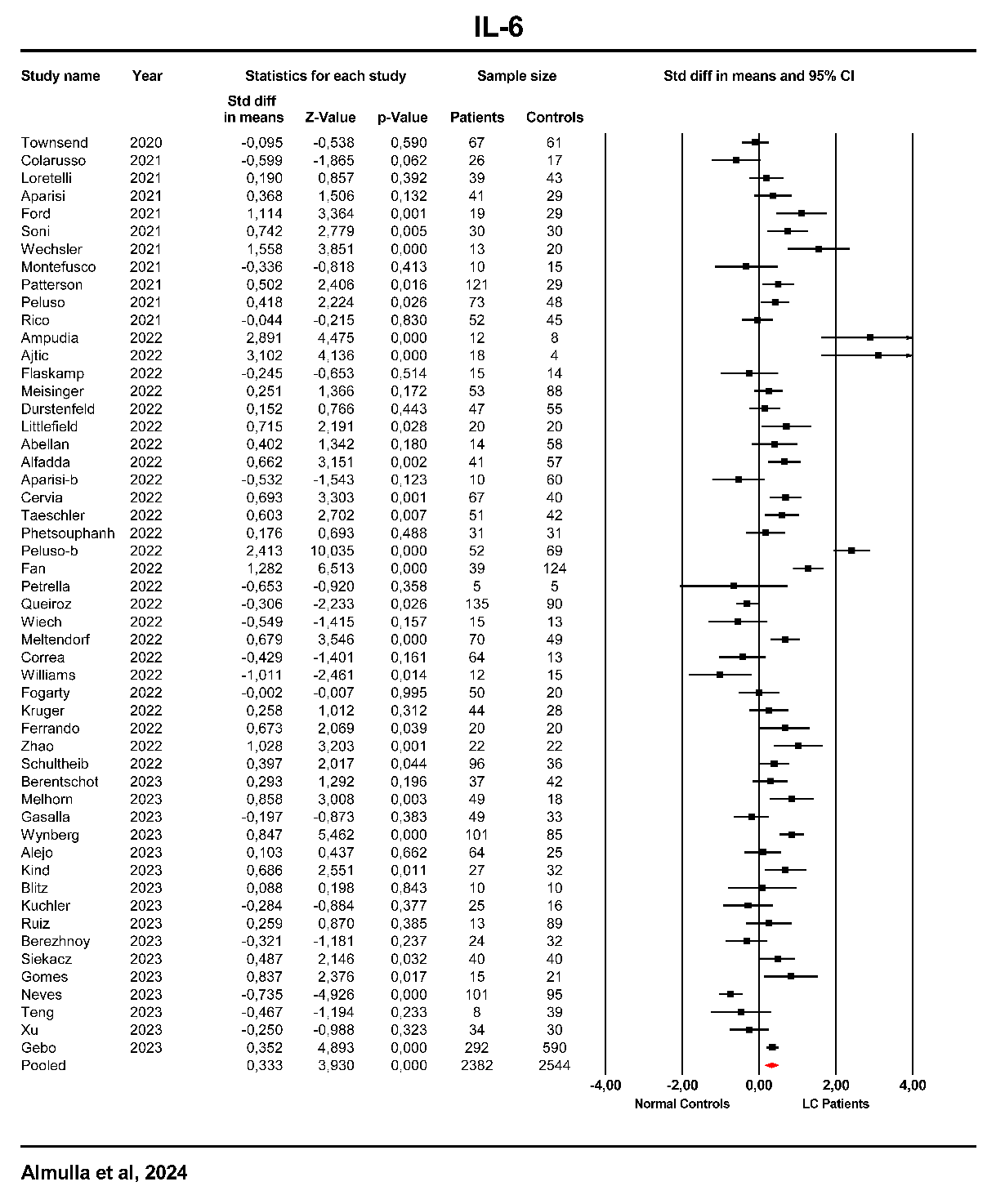

**ESF, Figure 7**. Forest plot of interleukin (IL)-6 in patients with Long COVID (LC) and normal controls.

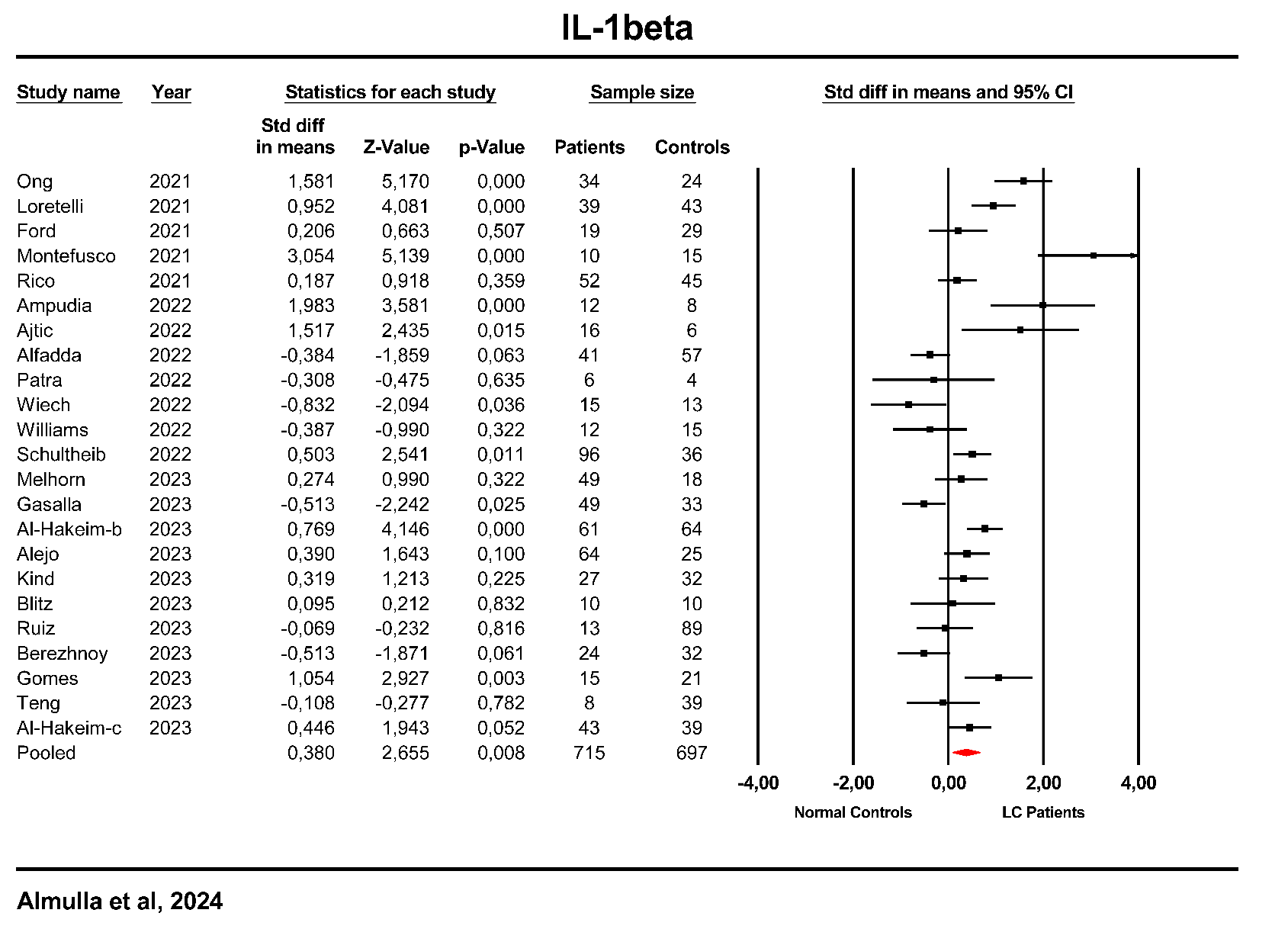

**ESF, Figure 8**. Forest plot of interleukin (IL)-1β in patients with Long COVID (LC) and normal controls.

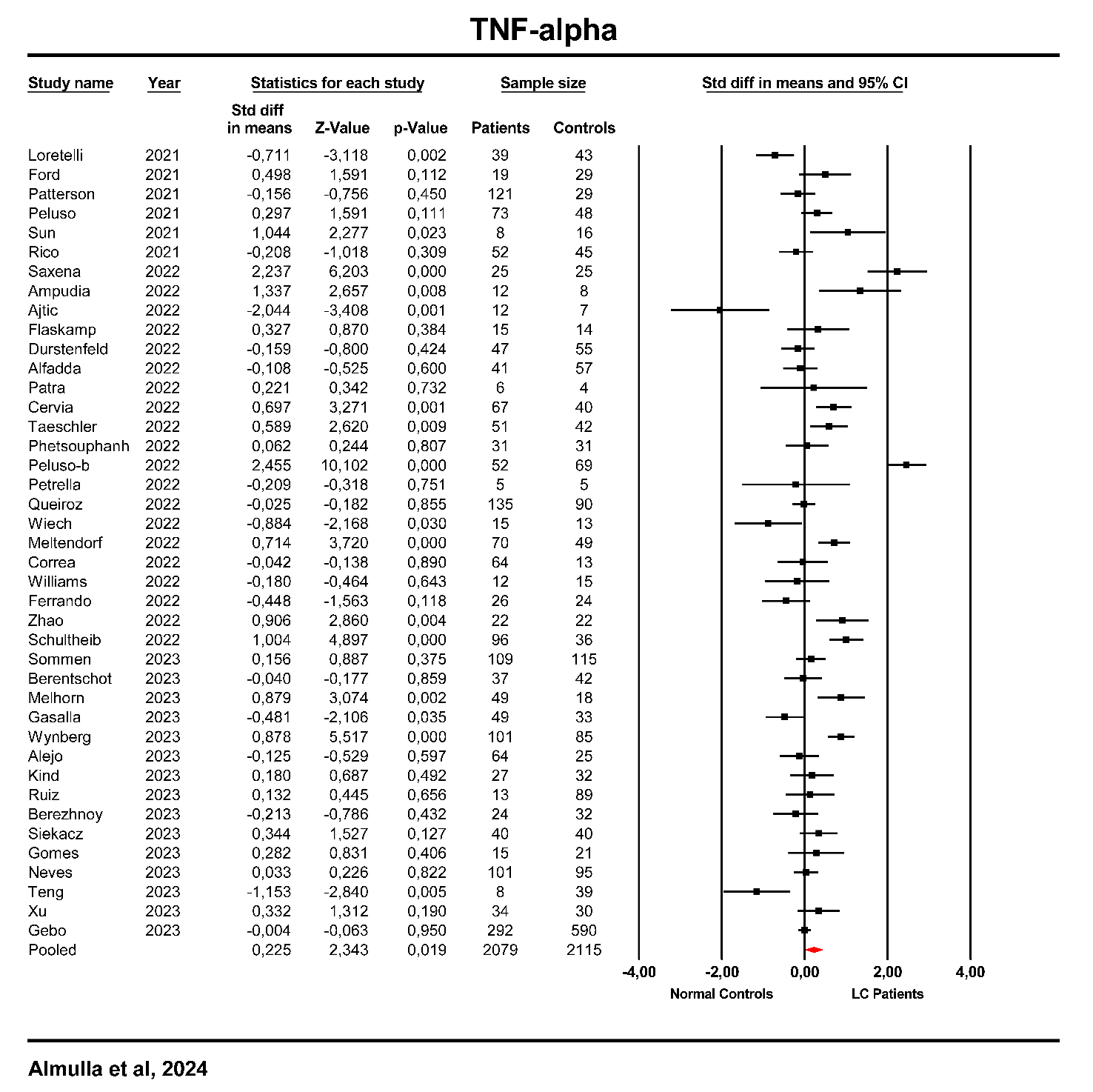

**ESF, Figure 9**. Forest plot of Tumor necrosis factor (TNF)-α in patients with Long COVID (LC) and normal controls.

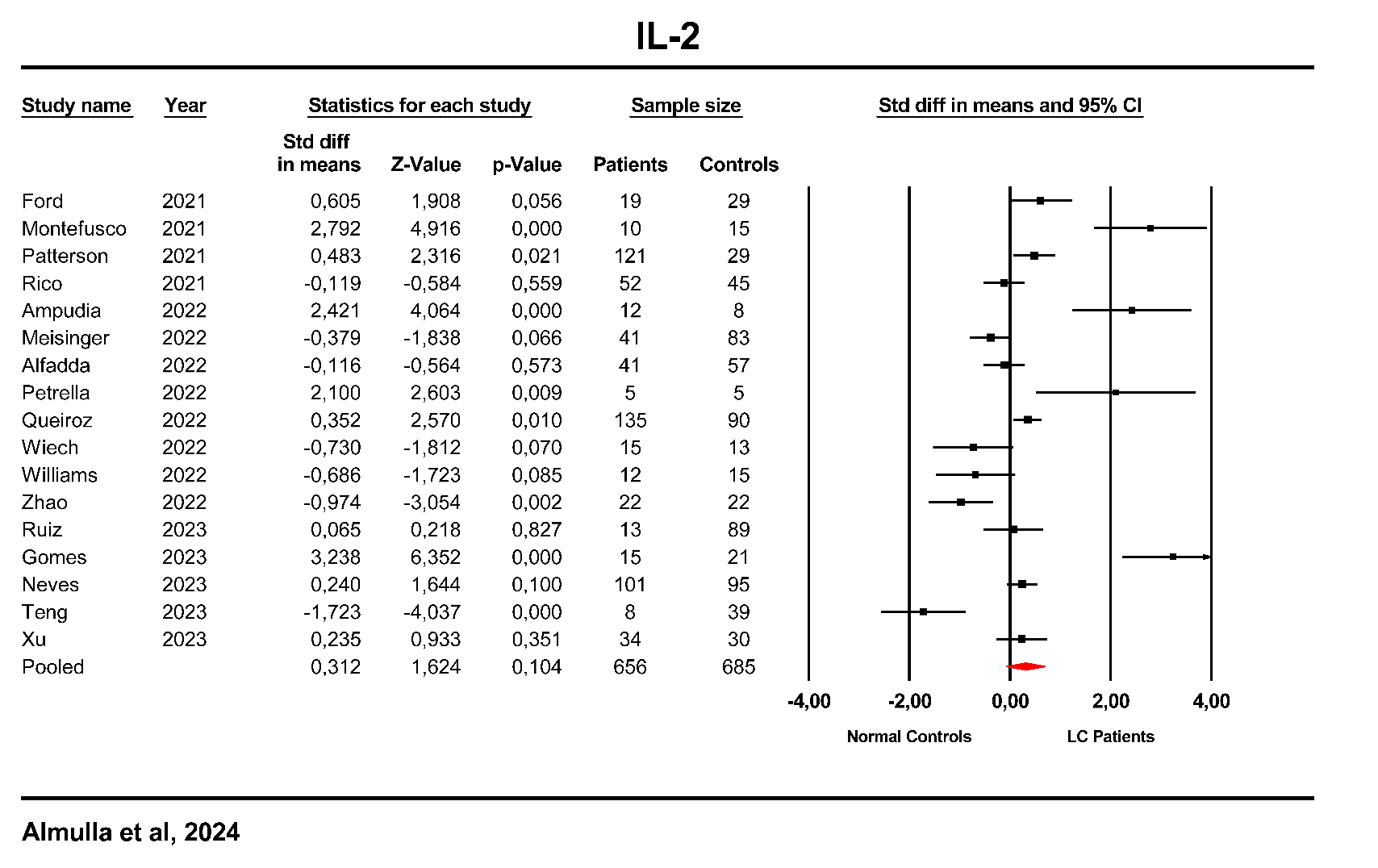

**ESF, Figure 10**. Forest plot of interleukin (IL)-2 in patients with Long COVID (LC) and normal controls.

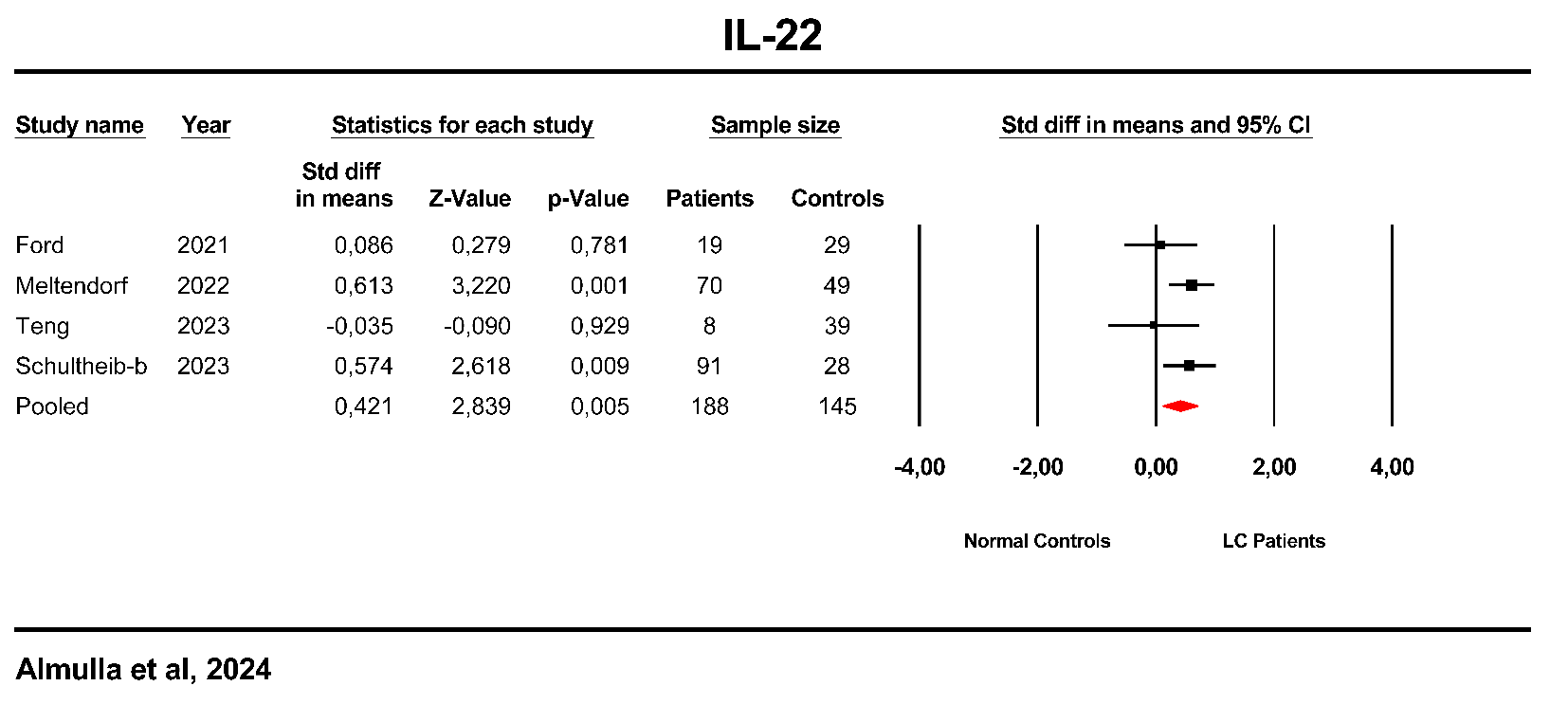

**ESF, Figure 11**. Forest plot of interleukin (IL)-22 in patients with Long COVID (LC) and normal controls.

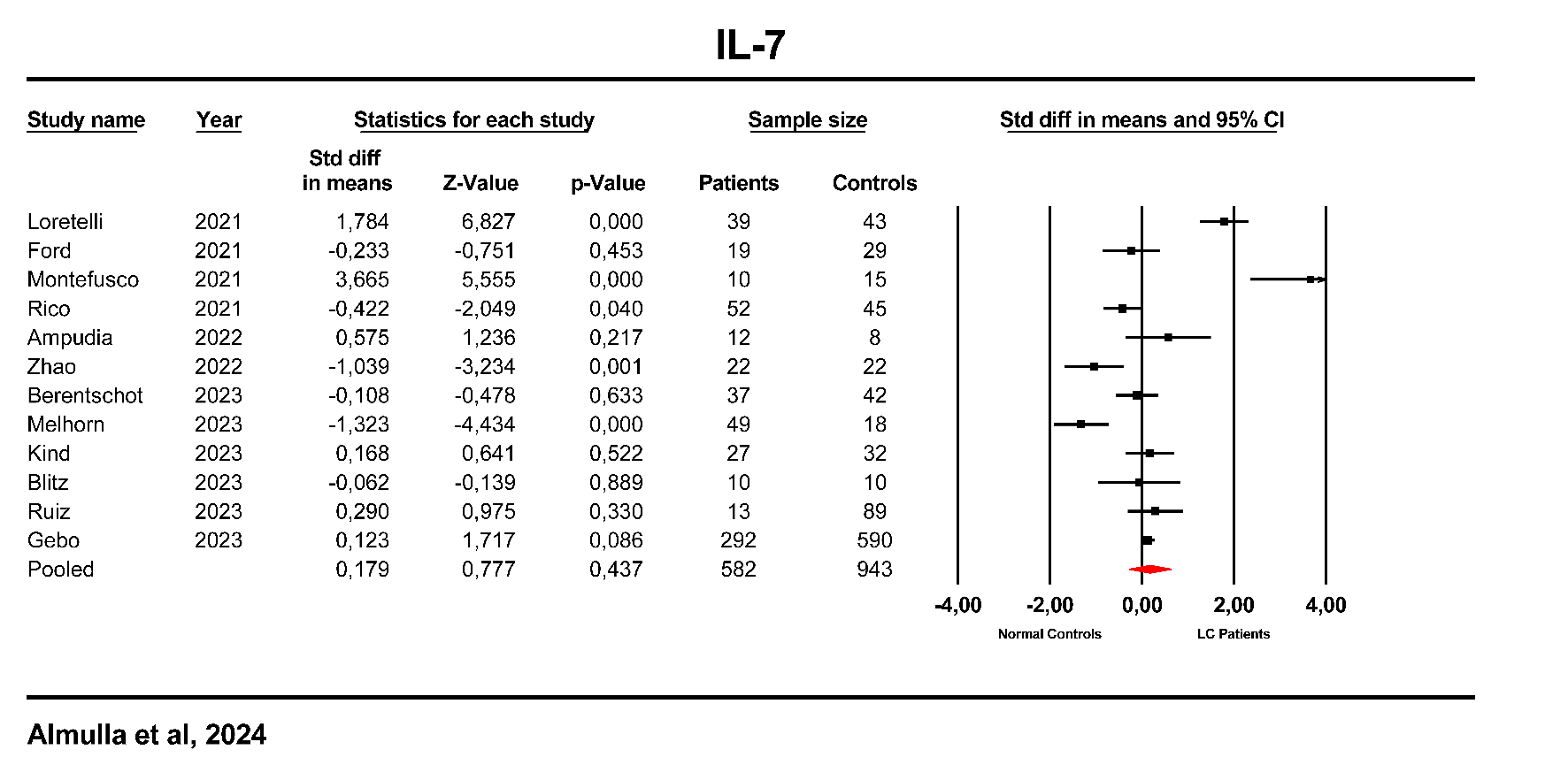

**ESF, Figure 12**. Forest plot of interleukin (IL)-7 in patients with Long COVID (LC) and normal controls.

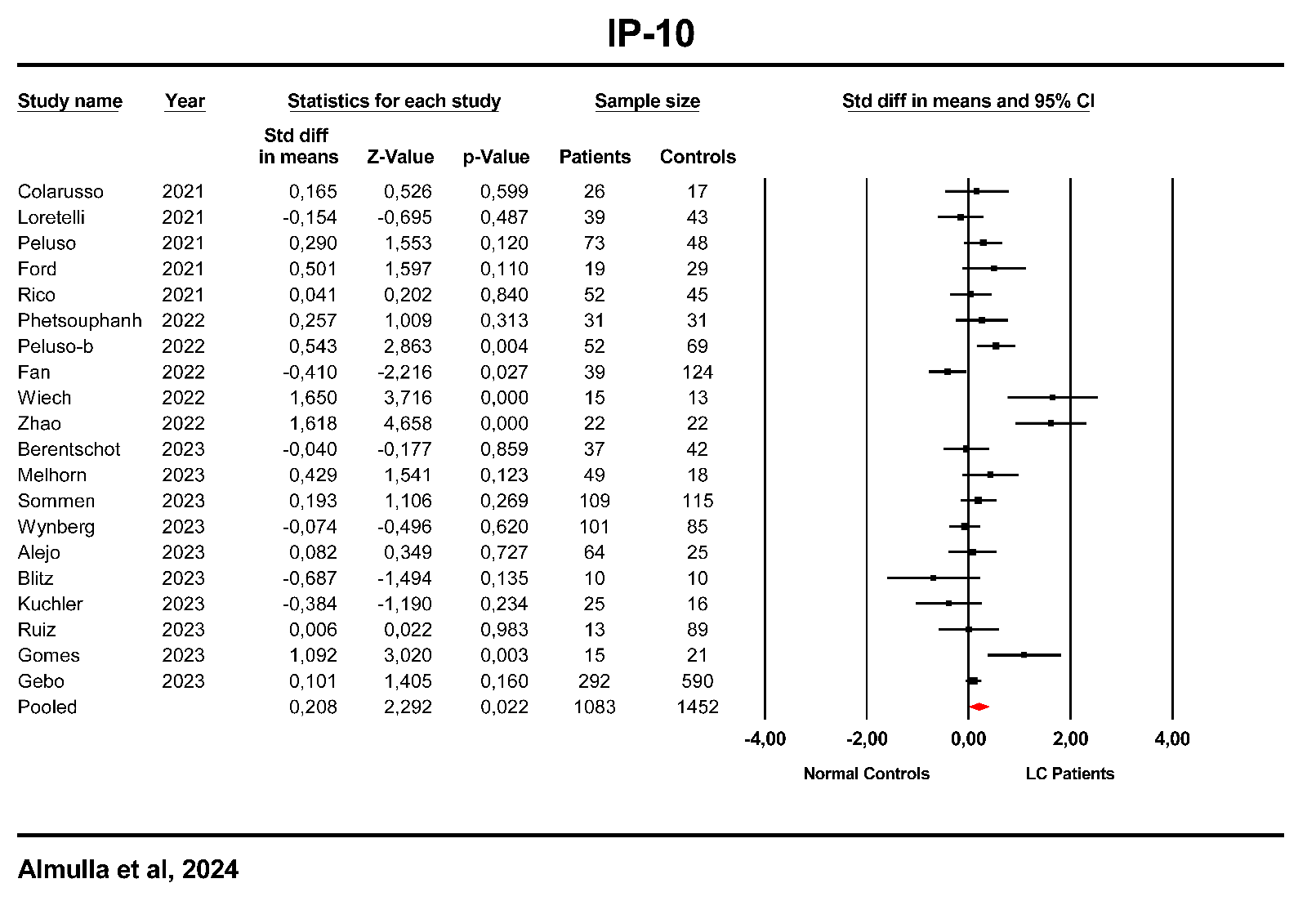

**ESF, Figure 13**. Forest plot of Interferon gamma-induced protein 10 (IP-10) in patients with Long COVID (LC) and normal controls.

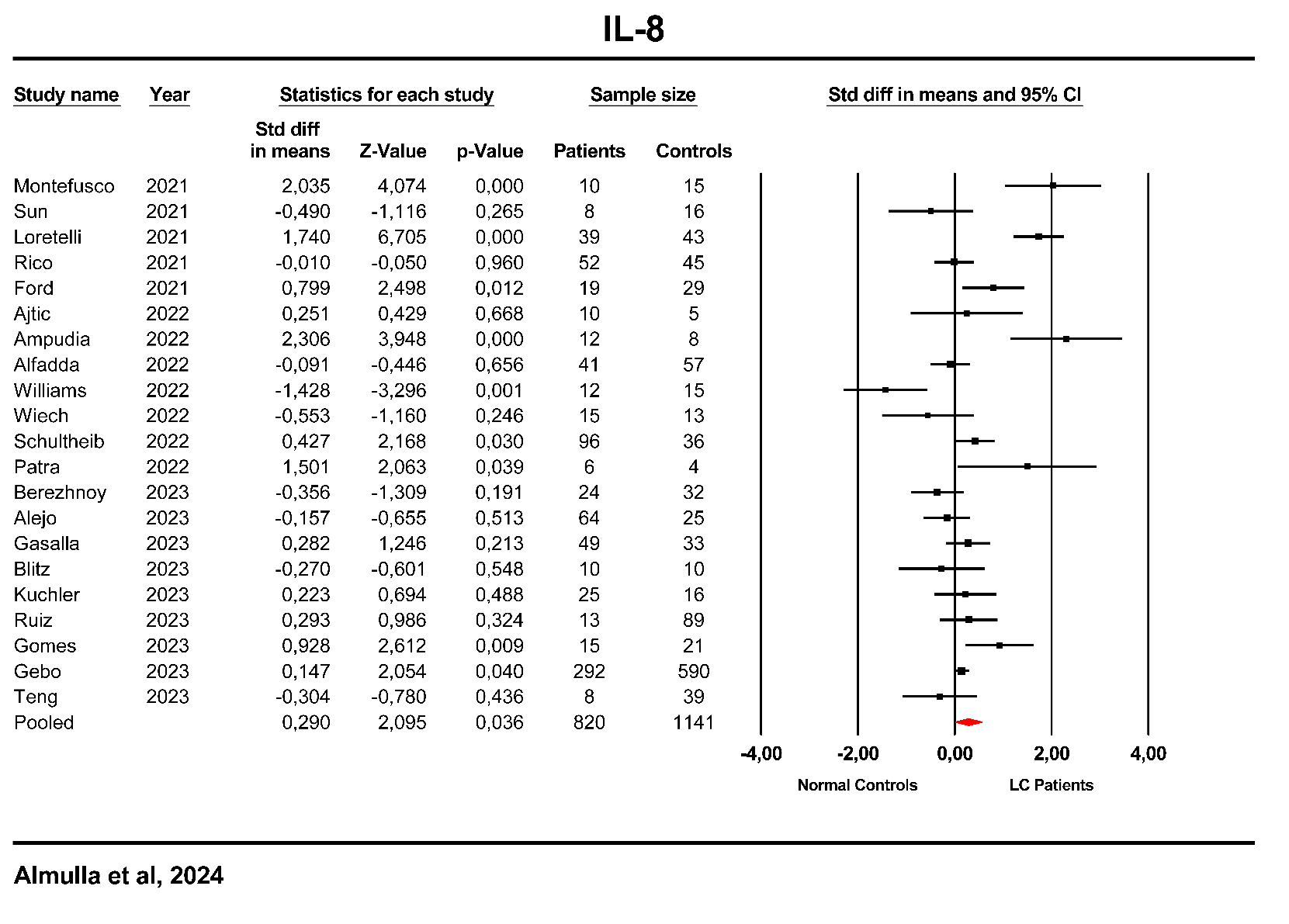

**ESF, Figure 14**. Forest plot of interleukin (IL)-8 in patients with Long COVID (LC) and normal controls.

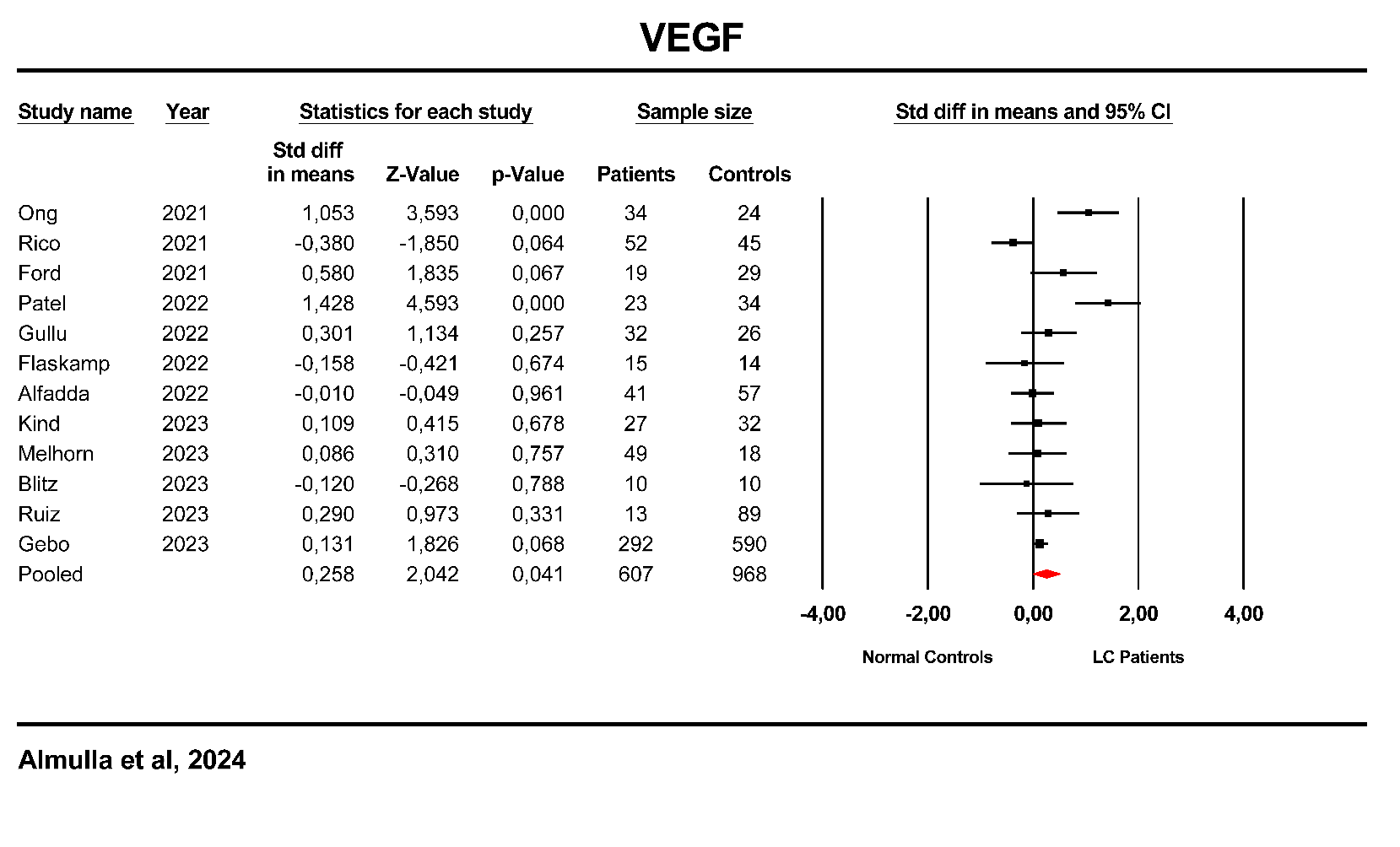

**ESF, Figure 15**. Forest plot of Vascular endothelial growth factor (VEGF) in patients with Long COVID (LC) and normal controls.

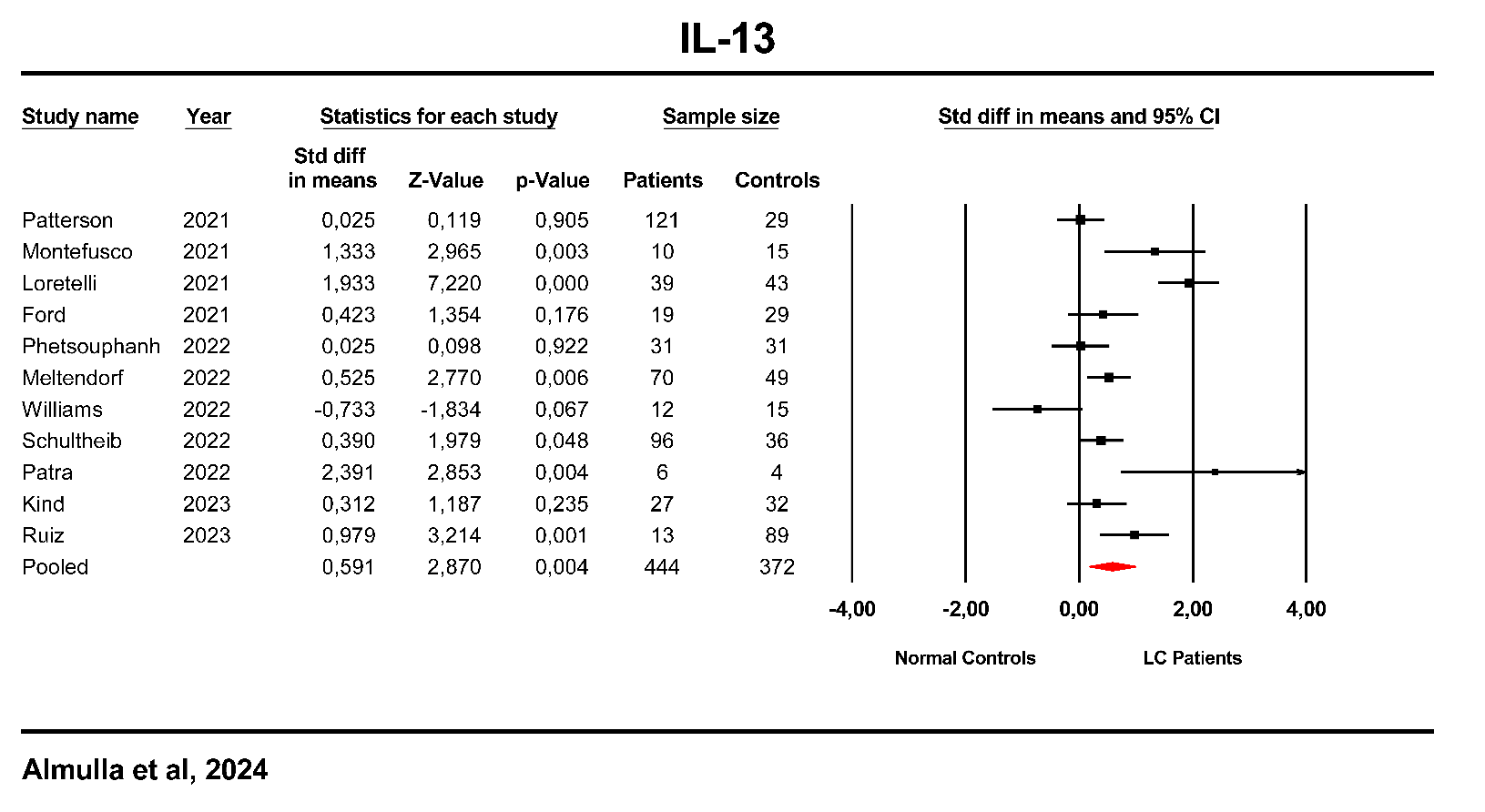

**ESF, Figure 16**. Forest plot of interleukin (IL)-13 in patients with Long COVID (LC) and normal controls.

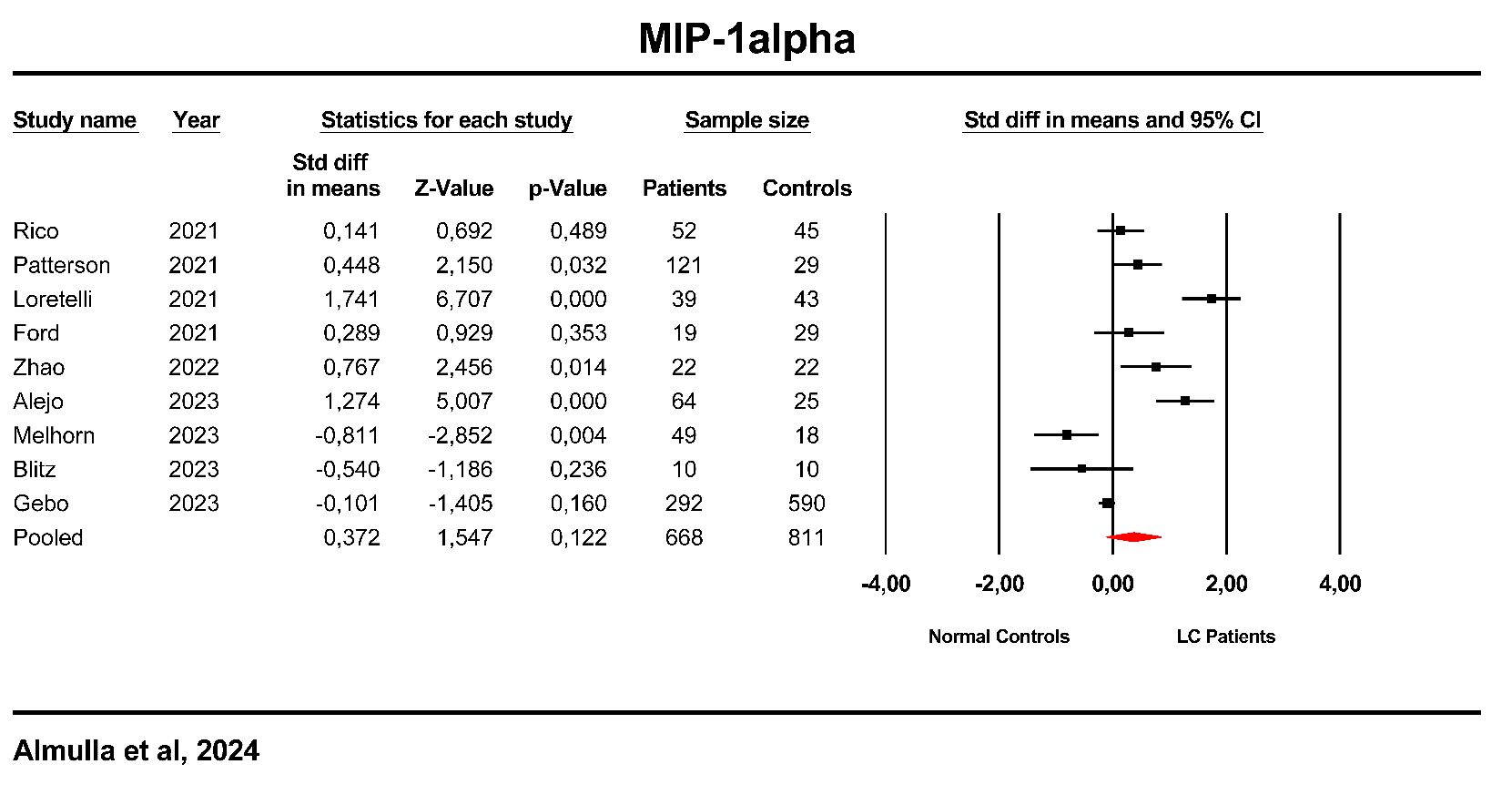

**ESF, Figure 17**. Forest plot of Macrophage inflammatory protein-1alpha (MIP-1α) in patients with Long COVID (LC) and normal controls.

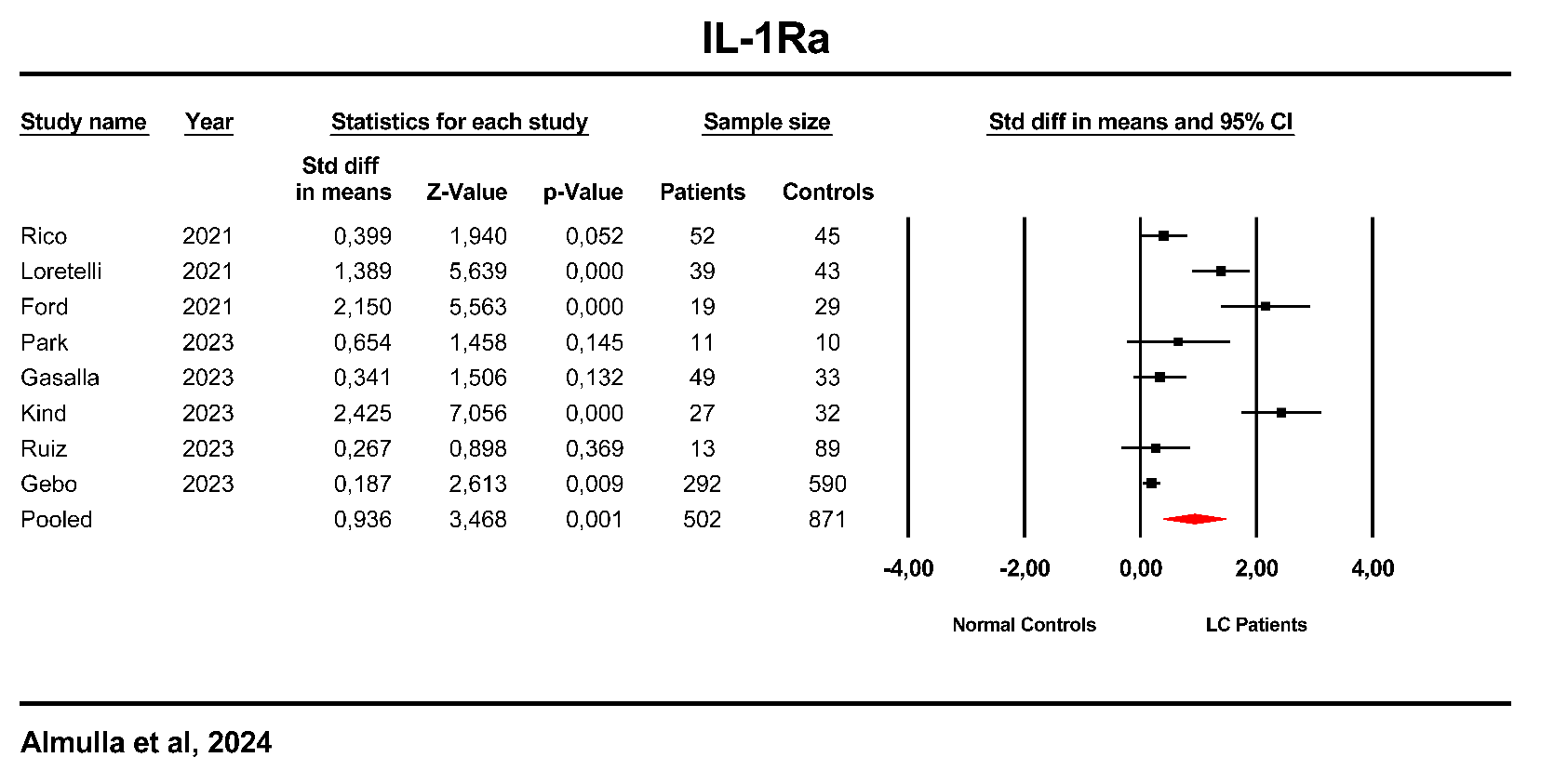

**ESF, Figure 18**. Forest plot of Interleukin-1 Receptor Antagonist (IL-1Ra) in patients with Long COVID (LC) and normal controls.

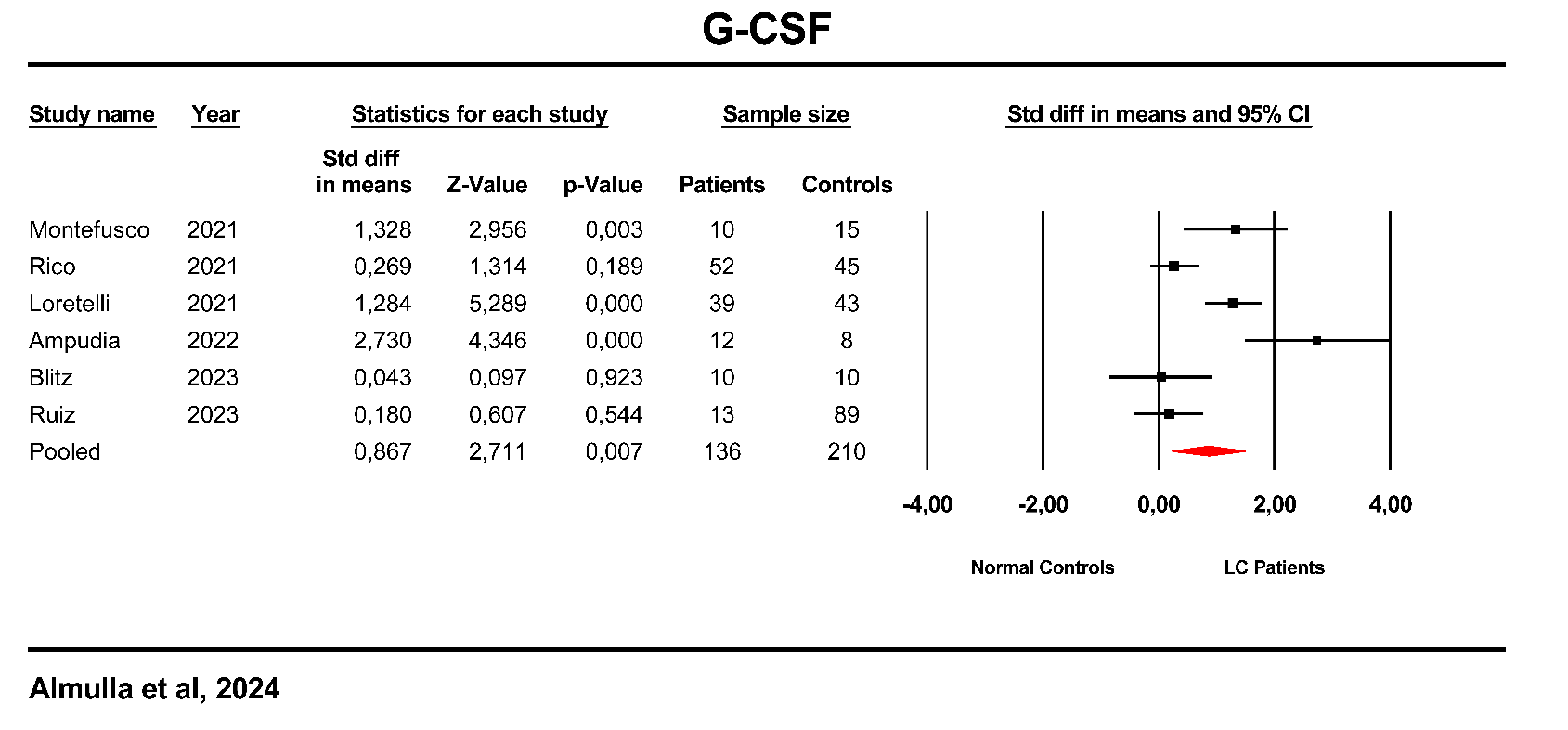

**ESF, Figure 19**. Forest plot of Granulocyte colony-stimulating factor (G-CSF) in patients with Long COVID (LC) and normal controls.

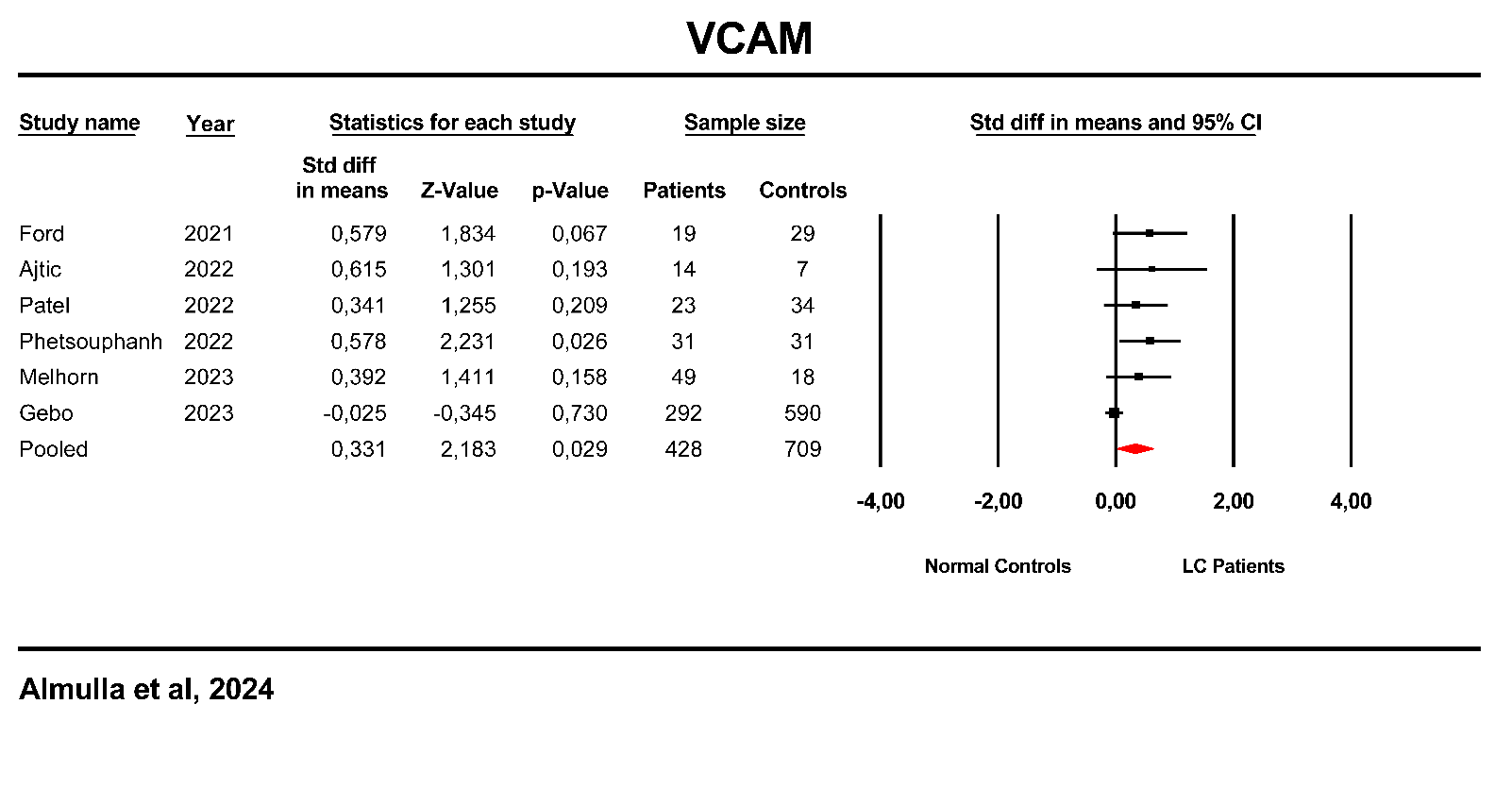

**ESF, Figure 20**. Forest plot of Vascular cell adhesion molecule-1 (VCAM) in patients with Long COVID (LC) and normal controls.

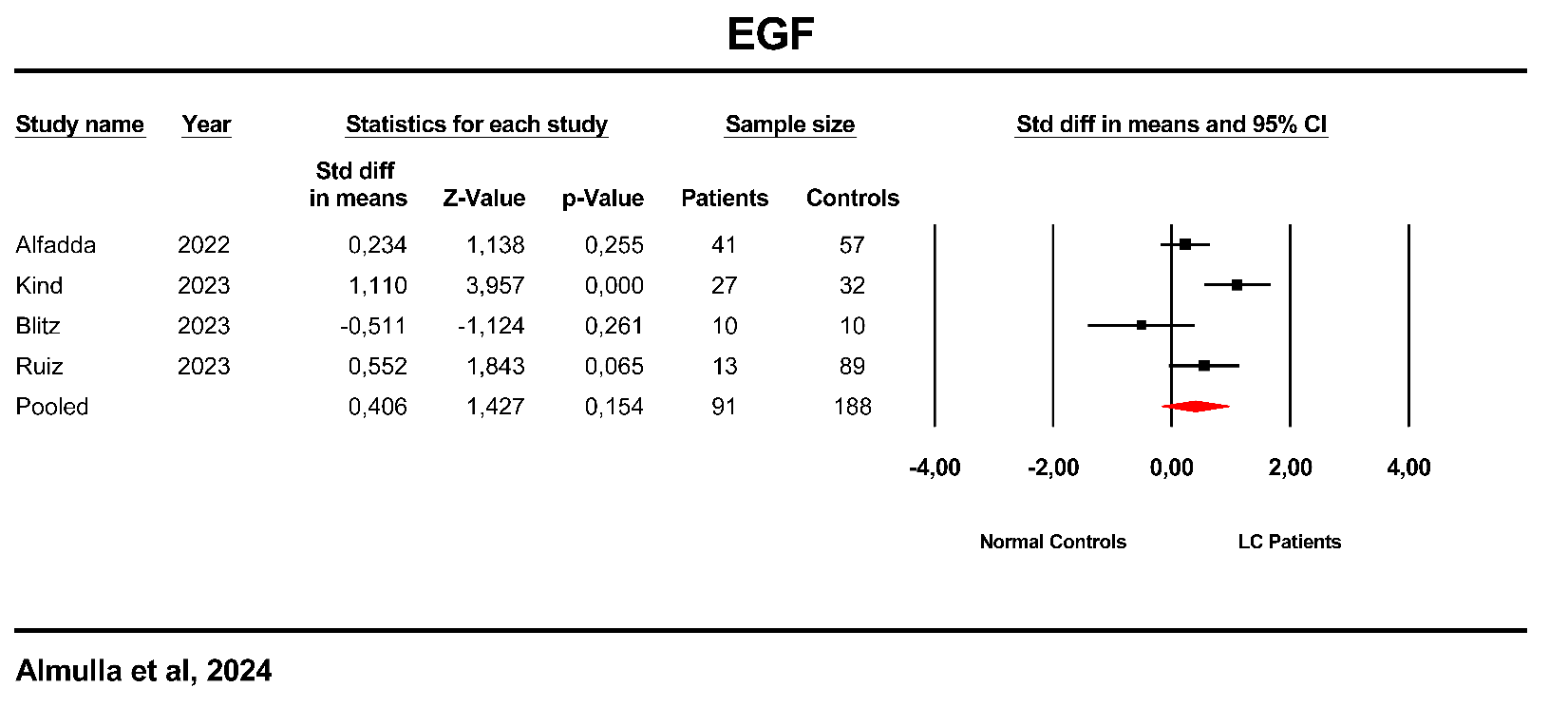

**ESF, Figure 21**. Forest plot of Epidermal growth factor (EGF) in patients with Long COVID (LC) and normal controls.

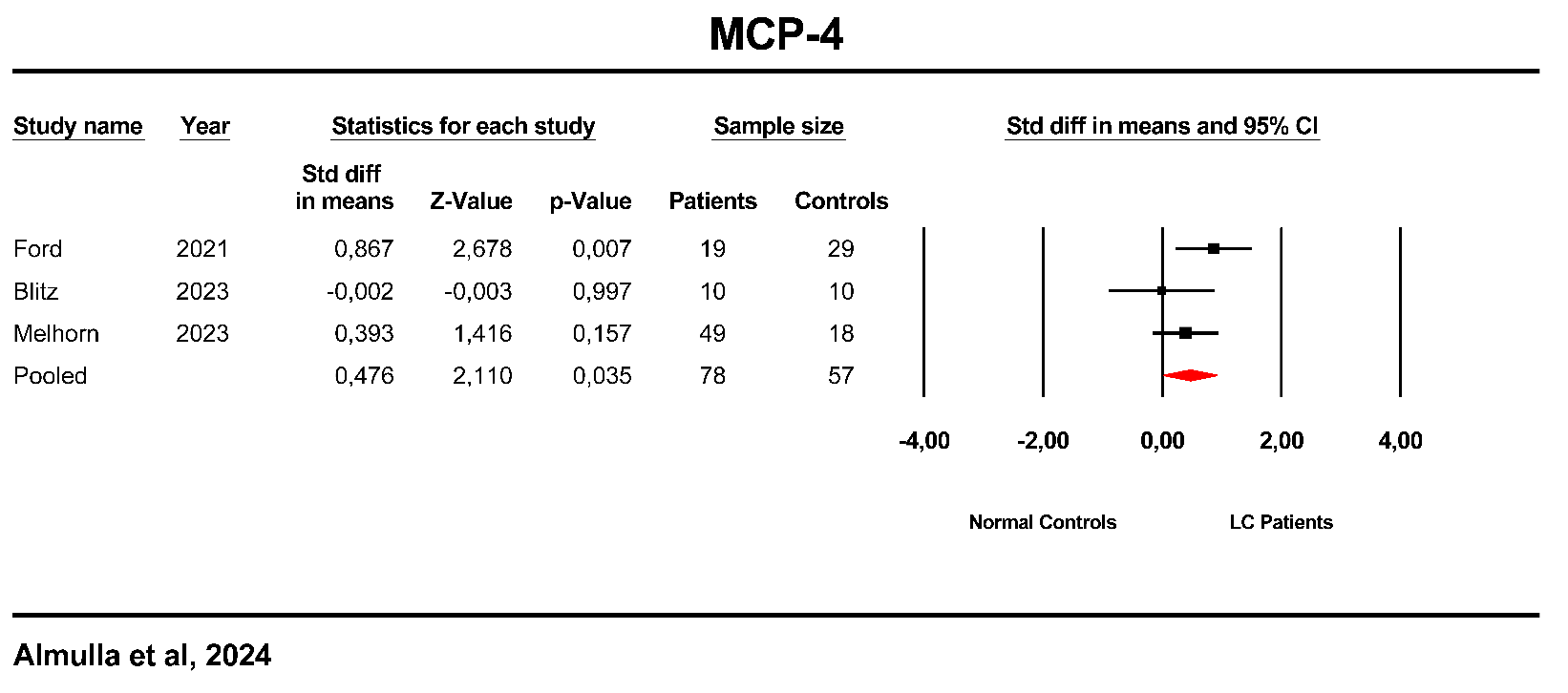

**ESF, Figure 22**. Forest plot of Monocyte chemoattractant protein (MCP)-4 in patients with Long COVID (LC) and normal controls.

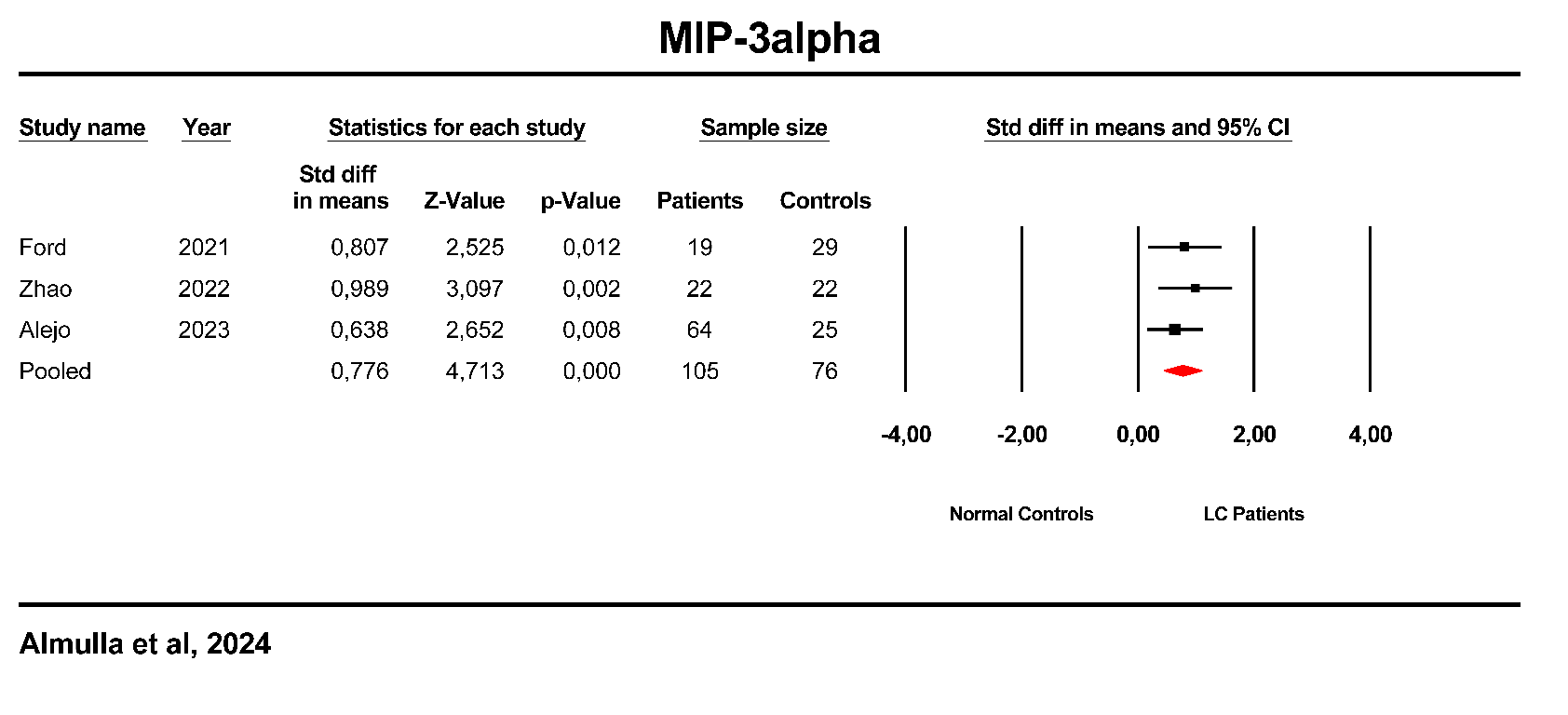

**ESF, Figure 23**. Forest plot of Macrophage Inflammatory Protein-3 Alpha (MIP-3α) in patients with Long COVID (LC) and normal controls.

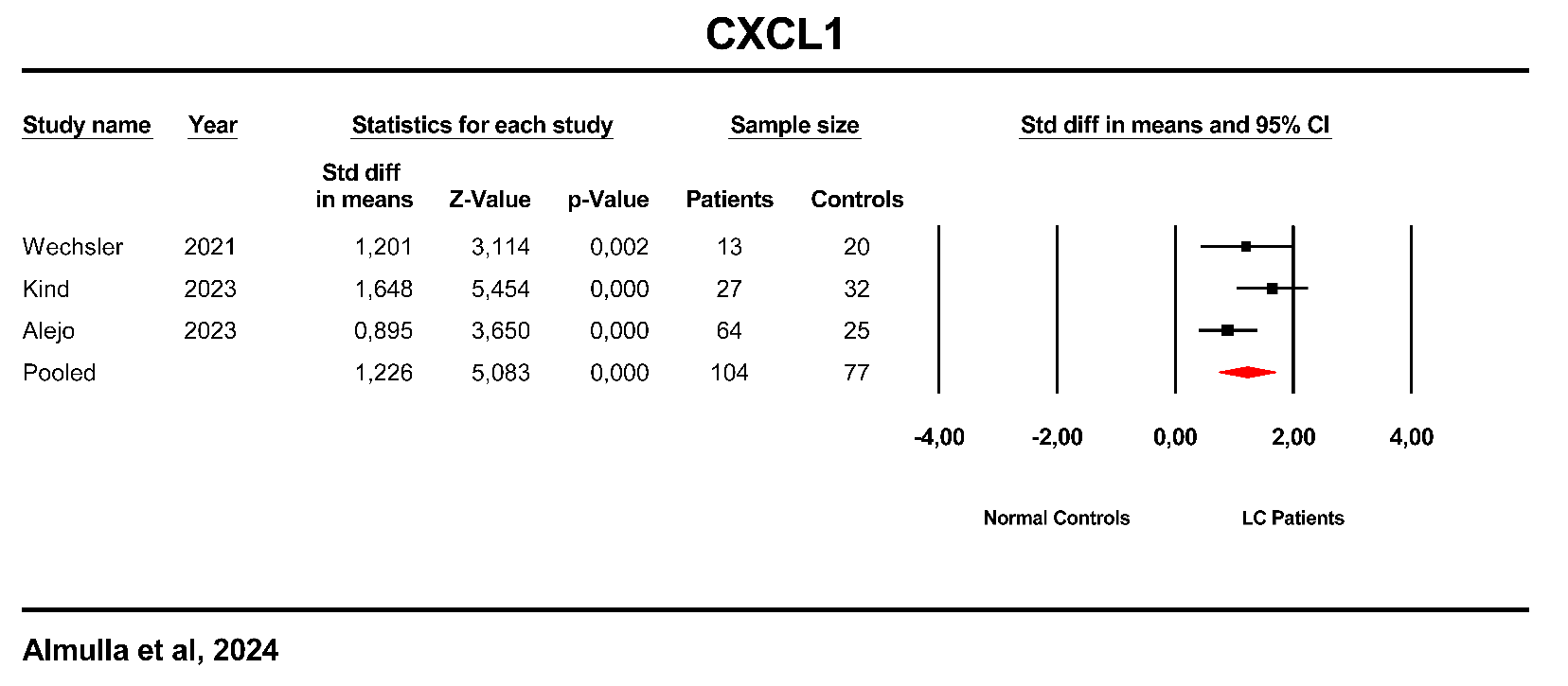

**ESF, Figure 24**. Forest plot of CXCL1 in patients with Long COVID (LC) and normal controls.

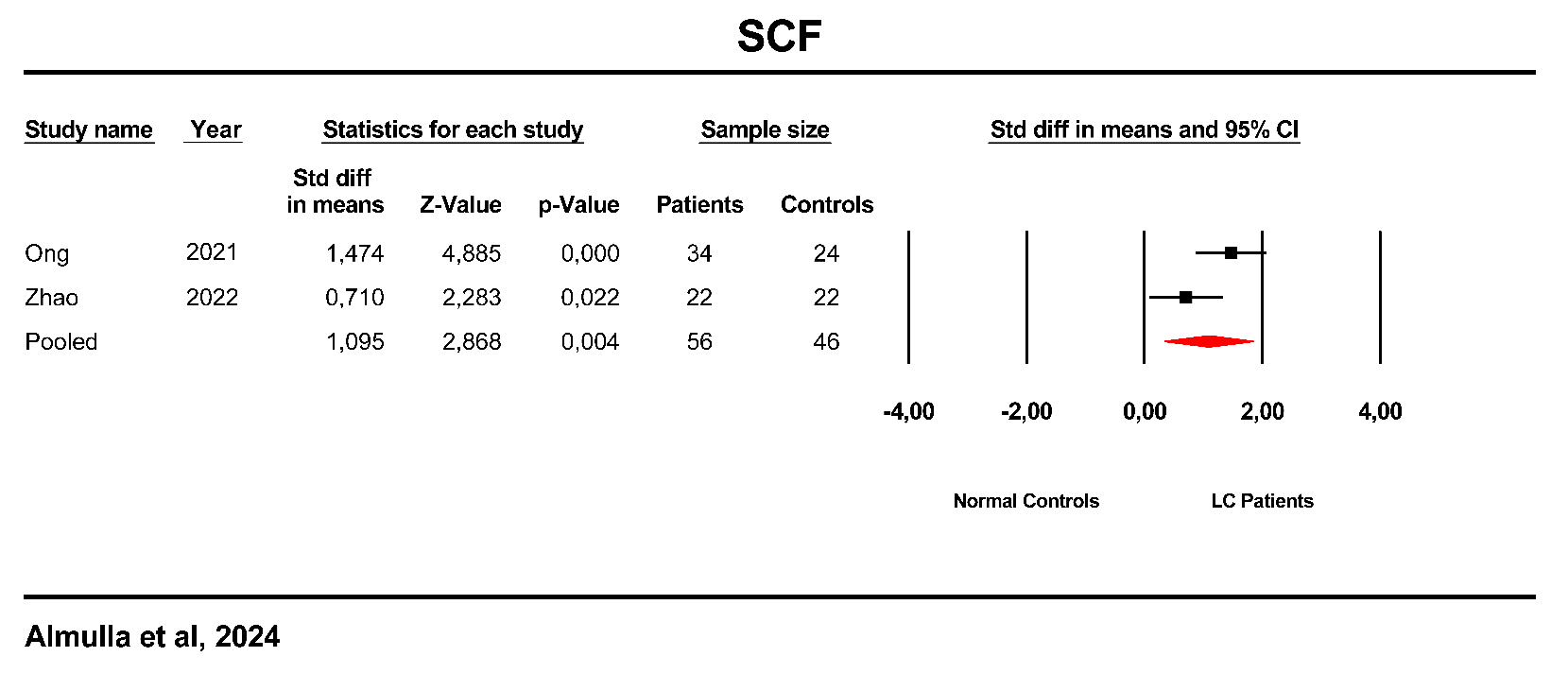

**ESF, Figure 25**. Forest plot of Stem cell factor (SCF) in patients with Long COVID (LC) and normal controls.
